# Electronic health record implementation: how to reduce the negative impacts

**DOI:** 10.64898/2026.03.24.26347438

**Authors:** Paula Fuscaldo Calderon, Nelson Wolosker

## Abstract

**Objective:** Develop a methodology to implement action plans that mitigate the negative impacts associated with the EHR implementation project and evaluate their effectiveness in reducing these issues.

**Methods:** The research involved the development of mitigation plans for the potential negative impacts of implementing an electronic health record system, ensuring their execution and subsequently analyzing the effectiveness of the method.

**Results:** Findings confirmed that 19.3% of 264 identified impacts were resolved through 52 plans before Go Live. During Go Live, the remaining 213 impacts were addressed through 337 plans. Six months later, 190 impacts were confirmed, and the plans were considered effective or partially effective in 80.5% of cases.

**Conclusions:** Effective governance, a multidisciplinary methodology, and well-planned and executed actions increase the likelihood of success for health technology projects.

## Introduction

Digital transformation has advanced rapidly on a global scale and, in the healthcare sector, has become a central element in improving services and outcomes in population health (1)(2)(3)(4). In this context, the acquisition and implementation of new healthcare technologies require institutions not only to manage projects efficiently, but also to have solid corporate governance and a well-structured change management strategy (1)(5). These initiatives often involve significant changes in workflows (6) and organizational culture, requiring professionals to adapt their daily practices and incorporate new ways of thinking and acting in their daily lives (7)(8)(9).

All organizational change requires planning, strategy, and continuous monitoring to reduce resistance among affected individuals and maximizing the expected benefits (10). In this context, change management plays an essential role in supporting professionals in accepting and adapting to new working models resulting from the technology adoption (5)(11).

When a healthcare institution decides to adopt technological solutions such as an Electronic Health Record (EHR), it becomes necessary to manage resistance not only from healthcare professionals (5)(12)(13)(14), but also from employees in other areas of the hospital (15)(12). The EHR implementation is a cause for great concern, mainly due to the high failure rates reported in the literature—often exceeding 50%—which can compromise the effort and time invested, in addition to generating significant financial losses. This risk is highly associated with the target audience adherence, which is not multifactorial and depends on several factors (16). First, for users to accept the system and be satisfied with its use, the EHR must be aligned with the institution’s existing processes and workflows, so that its quality and usefulness are perceived in practice (17). Second, strong support from the organization’s leadership is a key component in enabling and sustaining change (18).

The introduction of EHRs in hospitals generally requires careful review of workflows (19) to identify necessary adaptations, adjust processes, and provide adequate training (5)(15)(20)(21). However, these measures tend to be more effective when leaders and managers actively engage (14), who need to support implementation, demonstrate its importance, engage and motivate employees, and adopt a positive attitude toward change (7). In addition, it is essential to have a clear, effective communication plan for the institution’s main stakeholders (5)(20).

As part of the change management process in EHR implementation projects, it is necessary to identify in advance potential negative impacts on the daily lives of professionals involved in the change (6) and to adopt measures to promote successful implementation (10). The initial mapping of these negative impacts, followed by the stratification of problems by severity, facilitates the structuring of strategies to minimize problems (22)(23). It is assumed that implementing mitigation plans is more effective than attempting to repair damage or address difficulties after they occur (23). However, there are still no studies proving the effectiveness of these actions, and their evaluation is necessary to quantify the real impact on reducing problems during the system implementation.

### Objective

The objective of this study was to develop a methodology for implementing action plans to mitigate the negative impacts associated with the EHR implementation project, and to evaluate their effectiveness in reducing these problems.

### Methodology

This is a retrospective, descriptive study that examines the implementation of CERNER®’s Millennium software, version 2015. 01.09, Kansas, Missouri, USA, in all care areas of a hospital complex with 750 beds, an outpatient unit, and an emergency room (urgent and emergency care), which has 15,000 employees and 8,000 physicians. The project began in January 2014, and implementation (Go Live) took place in October 2016.

Thirty-eight leading professionals in their fields were hired, with extensive knowledge of hospital practices and representing various stakeholders, exclusively for the EHR implementation project. The group included: 10 physicians (including nephrologists, geriatricians, cardiologists/intensivists, anesthesiologists, oncologists, pediatricians, neonatologists, hematologists, and emergency physicians), 9 nurses from key sectors (ICUs, maternity, emergency room, oncology, surgical center, inpatient and outpatient), two biomedical professionals (imaging and laboratory), two physical therapists, one speech therapist, one nutritionist, four pharmacists, three registration professionals, four from the financial area, and two from the commercial area.

Key users mapped the main workflows for each hospital sector, focusing exclusively on negative impacts and identifying necessary changes (see attachment 1).

The team identified 264 potential negative impacts that could occur during the new EHR implementation process. Based on actions previously described in the project management literature, five action plans were developed to mitigate these impacts (see Table 1).

**Table 1.** Action Plans.

| Action Plan | Description |
| --- | --- |
| Training focus point | Address to the training development team the impacts that should receive attention during training. |
| Alignment with Area Manager | Prepare the manager of the affected area for change in their sector, as well as guide them in preparing their team. |
| Communication Plan | Address the impacts affecting interest groups in the institution's communication channels, such as forums and workshops. |
| Review and adaptation of the tool with the supplier | Discuss with the supplier adjustments to the tool to enable the implementation of a specific feature for executing a particular process. |
| Review and adaptation of institutional processes | Change a specific workflow or institutional process to enable the implementation of the tool. |

Each identified impact should be accompanied by at least one action plan to mitigate it or, if possible, eliminate it. The team had the autonomy to select the most appropriate mitigation plans for each situation.

Six months after implementing the system in the hospital complex, the action plans established to address anticipated impacts were analyzed, and the actions that eliminated the identified problems were verified. For impacts that were not resolved before Go Live, it was assessed which ones actually occurred. In cases of confirmed impacts, the project teams analyzed whether the action plans implemented were effective in minimizing them. When there was disagreement among the teams regarding the effectiveness of an action plan, which was considered effective in some hospital sectors and ineffective in others, it was classified as partially effective.

### Ethical considerations

The data in this study are retrospective, did not involve experimentation on humans or the use of tissues, and did not use sensitive data or identifiable information. Based on these premises, the Hospital Ethics Committee waived the need for formal approval and informed consent for publication.

**Table 1.** Descriptive analysis of mitigation plans (n=389).

| Mitigation Plan | Total |
| --- | --- |
| Training Focus Point | 197 (50.64%) |
| Alignment with Area Manager | 109 (28.02%) |
| Review and adaptation of the tool with the supplier | 32 (8.22%) |
| Communication Plan (Forums, Workshops) | 17 (4.37%) |
| Review and adaptation of the institutional process | 34 (8.74%) |
| Total | 389 (100%) |

## RESULTS

Among the 264 potential negative impacts identified, 389 action or mitigation plans were established. Most of these impacts received solutions focused on training and alignment with managers in the affected areas (Table 1).

Among the 264 negative impacts predicted for the EHR implementation project, 51 were resolved through action plans prior to Go Live, while the remaining 213 were mitigated during Go Live **(Figure 1).**

**Figure 1.**
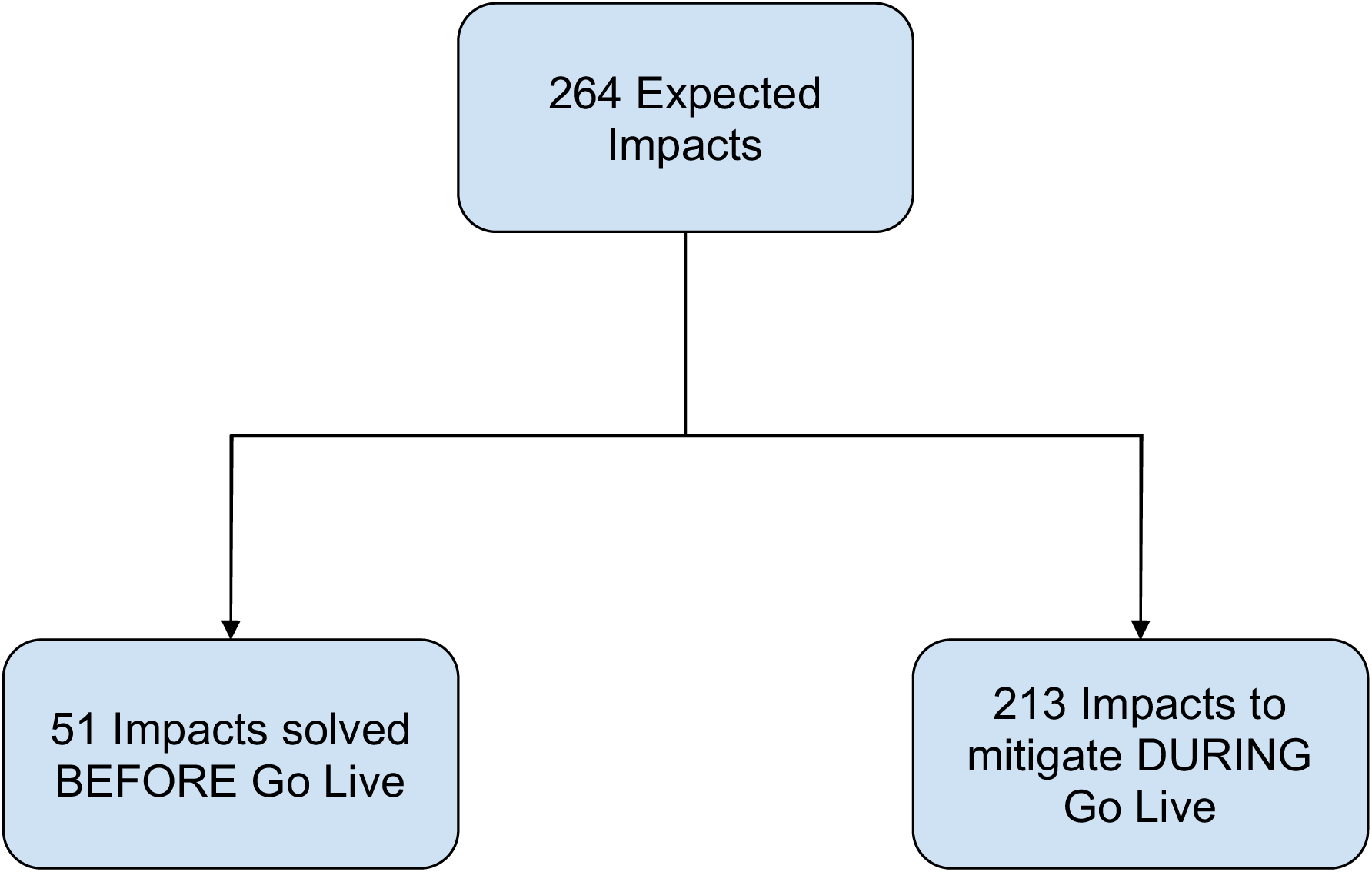
Distribution of expected impacts.

For the 51 impacts resolved prior to Go Live, 52 action plans were created, most of which were resolved through the review and adaptation of institutional processes, as detailed in Table 2.

**Table 2.** Descriptive analysis of action plans for resolved impacts (n=52).

| Action Plan Solved | Total |
| --- | --- |
| Training Focus Point | 00 (00.00%) |
| Alignment with Area Manager | 00 (00.00%) |
| Review and adaptation of the tool with the supplier | 18 (34.61%) |
| Communication Plan (Forums, Workshops) | 00 (00.00%) |
| Review and adaptation of the institutional process | 34 (65.38%) |
| Total | 52 (100%) |

For the remaining 213 impacts, 337 action plans were established. More than 50% of these impacts were addressed as priority points in training, and more than 30% through alignment with managers in the affected areas (Table 3).

**Table 3.** Descriptive analysis of impact mitigation plans at Go Live (n=337).

| Mitigation Plan for Go Live | Total |
| --- | --- |
| Training Focus Point | 197 (58.45%) |
| Alignment with Area Manager | 109 (32.34%) |
| Review and adaptation of the tool with the supplier | 14 (4.15%) |
| Communication Plan (Forums, Workshops) | 17 (5.04%) |
| Review and adaptation of the institutional process | 00 (00.00%) |
| Total | 337 (100%) |

In the comparative analysis between the mitigation plans implemented for impacts resolved before Go Live (52 plans) and those that needed to be mitigated during Go Live (337 plans), we observed that, for impacts resolved prior to implementation, the structural approach predominated, with 65.38% of plans focused on reviewing and adapting institutional processes and 34.61% on reviewing and adapting the tool with the supplier. During Go Live, the focus shifted to people-oriented actions: 58.45% of the plans were treated as points of attention in training, and 32.34% involved alignment with area managers, as shown in Table 4.

**Table 4.** Comparative analysis of impact mitigation plans resolved before and during Go Live (n=389).

| Mitigation Plan | Solved<br>BEFORE Go<br>Live | To be solved<br>DURING Go<br>Live | Total |
| --- | --- | --- | --- |
| Training Focus Point | 00 (00.00%) | 197 (58.45%) | 197 (50.64%) |
| Alignment with Area Manager | 00 (00.00%) | 109 (32.34%) | 109 (28.02%) |
| Review and adaptation of the tool with the supplier | 18 (34.61%) | 14 (4.15%) | 32 (8.22%) |
| Communication Plan (Forums, Workshops) | 00 (00.00%) | 17 (5.04%) | 17 (4.39%) |
| Review and adaptation of the institutional process | 34 (65.38%) | 00 (00.00%) | 34 (8.74%) |
| Total | 52 (100%) | 337 (100%) | 389 (100%) |

Six months after Go Live at the hospital complex, we interviewed key users to assess the 213 anticipated impacts that had not been resolved during the design phase and were therefore managed during Go Live. In the users’ opinion, 190 of the anticipated impacts were confirmed, i.e., they occurred during Go Live. In 117 of these, the action plans were considered effective in mitigating the problems, and in 37, partially effective, according to the users’ perceptions (Table 5).

**Table 5.**
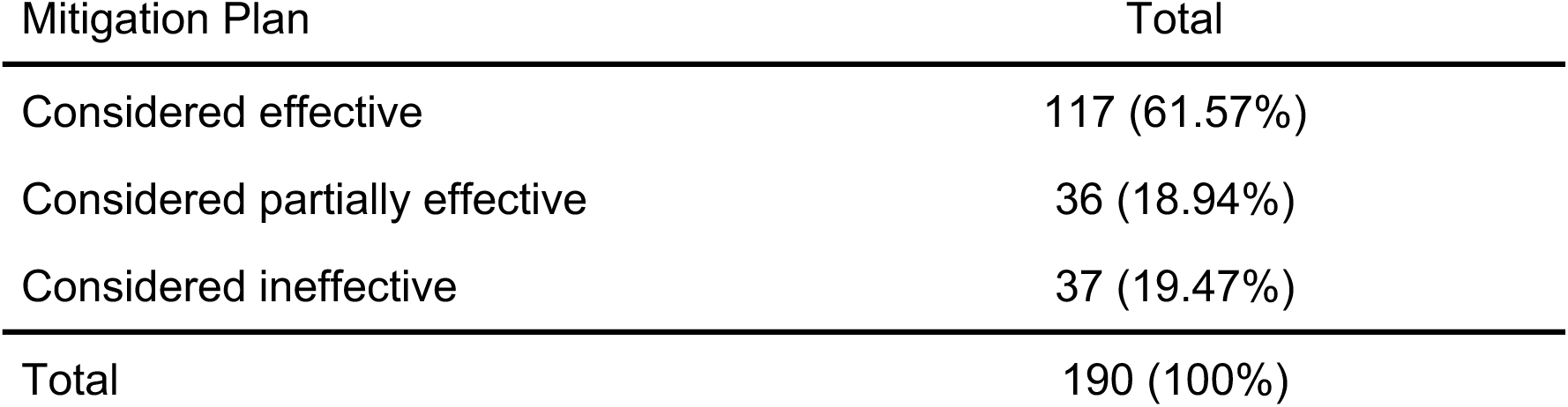
Effectiveness of the Action Plan (n=190).

| Mitigation Plan | Total |
| --- | --- |
| Considered effective | 117 (61.57%) |
| Considered partially effective | 36 (18.94%) |
| Considered ineffective | 37 (19.47%) |
| Total | 190 (100%) |

In 80.51% of cases, action plans were considered effective or at least partially effective in minimizing the impact of the changes.

## DISCUSSION

PEP implementations in hospital settings are complex initiatives aimed at improving the quality of care, patient safety, and operational efficiency. However, this process faces significant challenges. Typically, users are required to undergo prior training on how to use the tool, which is an integral part of the schedule for any project of this type (18)(21) and usually takes place a few weeks before Go Live. This training usually covers the main features and their use, with the aim of empowering users. However, when training is designed with a focus on critical processes that involve profound changes in the way of working, it becomes much more effective. In the present project, the training was developed prioritizing precisely these points considered critical.

A structured and multidisciplinary methodology was used to implement the EHR, in line with best practices for change management in a hospital environment. The initiative, with the participation of professionals from various fields (doctors, nurses, pharmacists, physical therapists, speech therapists, nutritionists, biomedical professionals, and administrators), enabled the comprehensive identification of 264 potential negative impacts associated with EHR implementation, accounting for different perspectives. Based on this mapping, 389 specific action plans were developed to mitigate this situation. Subsequently, these plans were analyzed to assess their effectiveness, identify which impacts were eliminated and which occurred, and determine to what extent the implemented measures were successful.

Among the 264 anticipated impacts, 51 were resolved during the pre-Go Live phase through 52 specific action plans. Most of these resolutions involved adapting existing institutional processes or adjusting the provided tool so that its functionality aligned with the institution’s operational routines.

It is important to note that there was no imposition on key users regarding the choice of action plans to be adopted. Decisions were made collaboratively, respecting the perceptions and experiences of those involved in day-to-day operations. This decentralized approach fostered greater commitment and effectiveness in the solutions applied.

In cases where institutional processes were modified to align with the software’s functionality, the impacts were fully resolved. The project team, in partnership with the leaders of each area, made a significant effort to promote the necessary changes and adjust the way employees worked. As this effort took place before Go Live, there was no need to give these points specific attention at the time of the system changeover, as users were already fully adapted to the new routines.

Although the purchased tool was a ready-to-install product, the supplier was able to customize features in some cases to suit the institution’s work processes and culture. These adaptations helped eliminate several impacts, so no mitigation actions were required during Go Live.

Although many problems had already been solved beforehand, there were still 213 impacts expected to occur during Go Live, and the team’s goal was to minimize them as much as possible. To this end, several action plans were implemented, such as: treating the impacts as points of attention in training; aligning with managers in the affected areas to prepare their teams for the changes that would occur; addressing certain problems in demonstrations of the tool in forums and workshops; and attempting to adapt the supplier’s product features to the institution’s work processes.

There were limitations to customizing the supplier’s tool, as the contract provided for the delivery of a ready-made product, without additional development. Thus, even when complete adjustments were not feasible, the team sought alternative solutions through internal parameterisations of the system. These configurations made the product more intuitive and functional for end users, helping to mitigate the impacts. Because each impact could receive multiple action plans, it was not possible to conduct a comparative analysis to determine which was most effective. However, this was not the focus of the study, and in many cases the combination of actions was decisive in achieving a satisfactory result.

Go Live took place on the previously scheduled date and, after implementation in the hospital complex, a six-month waiting period was observed for processes to stabilize, workflows to settle, and users to complete their learning curve. This study, designed to support successful software implementation, conducted interviews with key users to assess the 213 anticipated impacts that had not been resolved during the design phase, and each was reviewed in a joint meeting. According to the key user’s perception, 190 impacts were confirmed in GoLive, and the method of identifying mitigation plans was considered effective in 61.57% of cases, partially effective in 18.94%, and ineffective in 19.47%. Combining the effective and partially effective results yielded an approximately 80% resolution rate in mitigating problems at Go Live.

The experience in implementing the EHR highlights the complexity of this type of process and the need for a multidisciplinary approach to mitigate potential negative impacts. The identification of prior impacts and the development of mitigation strategies through specific action plans are fundamental to the project’s success.

In addition, the emphasis on training and alignment with area managers highlights the importance of team involvement and training in the adoption of new technologies. Users’ positive assessment of the effectiveness of action plans reinforces the relevance of these strategies for a successful transition to EHR use.

This experience can serve as a reference for other healthcare institutions seeking to implement EHR systems, highlighting best practices and lessons learned to mitigate the challenges associated with this digital transformation.

## CONCLUSIONS

After identifying 264 potential impacts on EHR implementation, 389 action plans were created, most of which focused on training (50.6%) and alignment with area managers (28%).

Before Go Live, 51 (19.3%) impacts were resolved through 52 plans, generating structural actions (65.4% process review and 34.6% tool adaptation). During Go Live, the remaining 213 impacts were addressed through 337 plans: 58.5% focused on training and 32.3% on alignment with managers.

Six months later, 190 impacts were confirmed, and the plans were considered effective or partially effective in 80.5% of cases.

These findings reinforce the concept that effective governance, a multidisciplinary methodology, and well-planned and executed actions increase the likelihood of success for health technology projects. Priority should be given to advance mapping, strategic allocation of training resources, and continuous engagement and assessment of key user perceptions.

## Data Availability

All data produced in the present study are available upon reasonable request to the authors

---

**Attachment 1.**
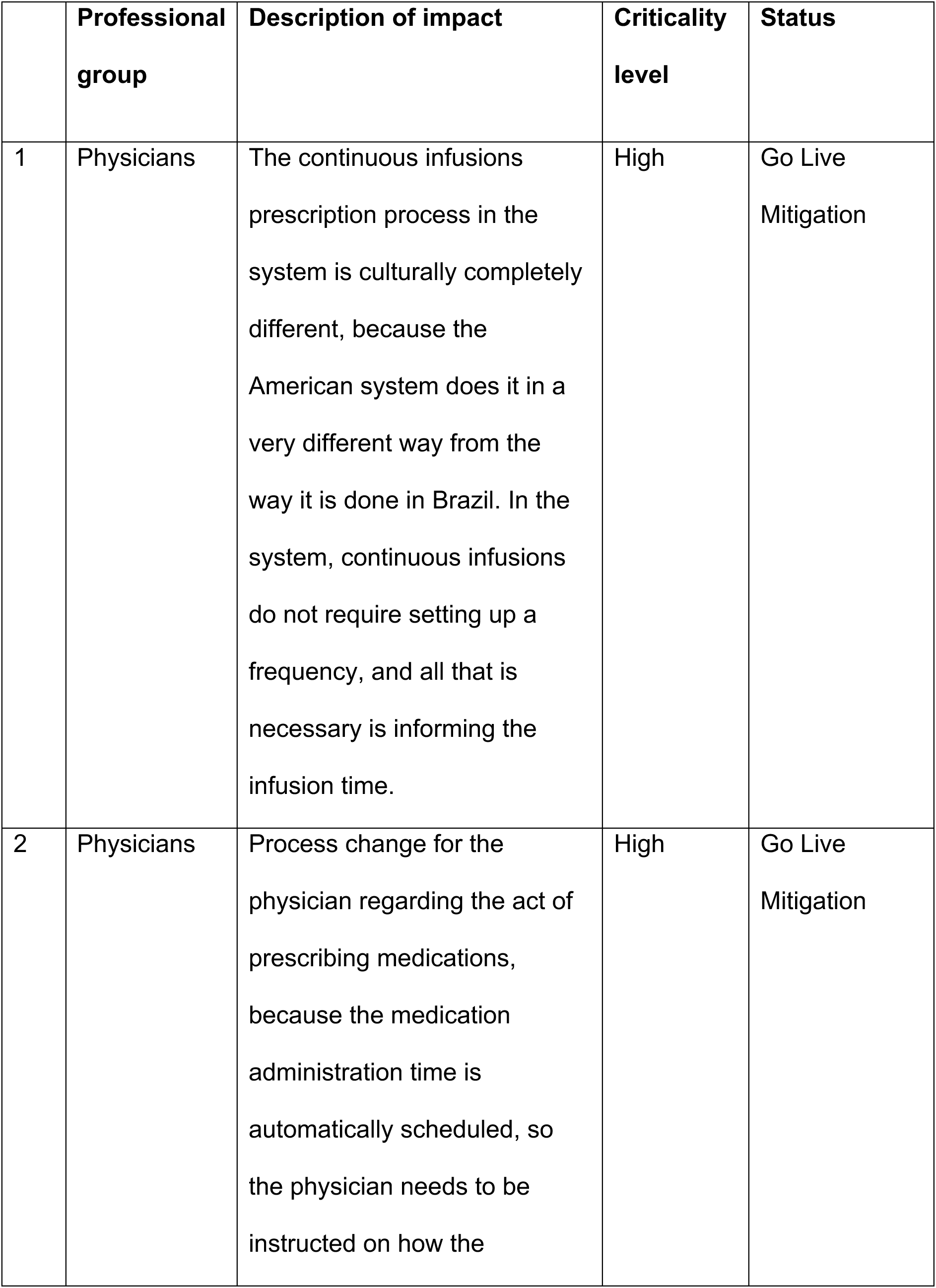
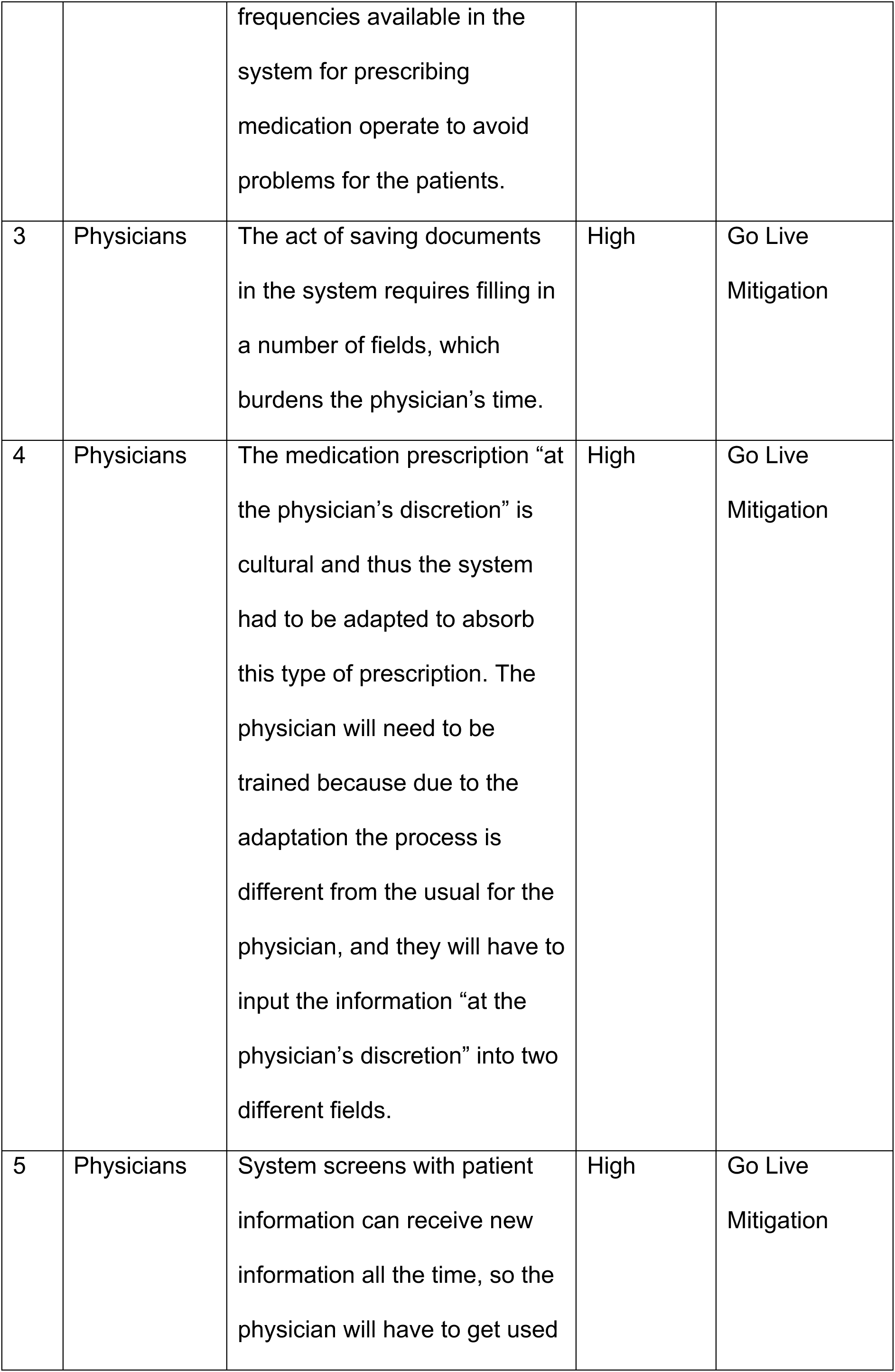
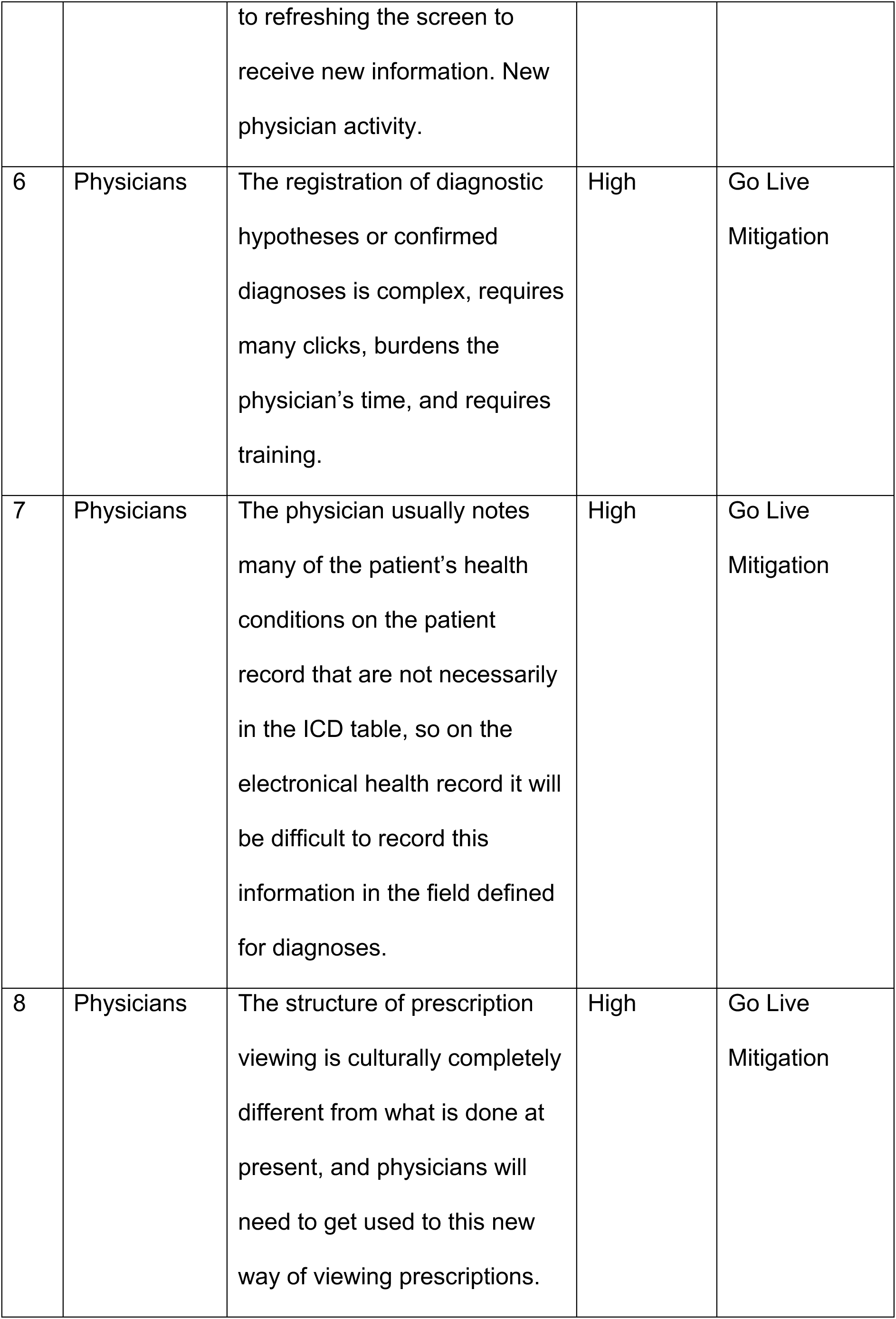
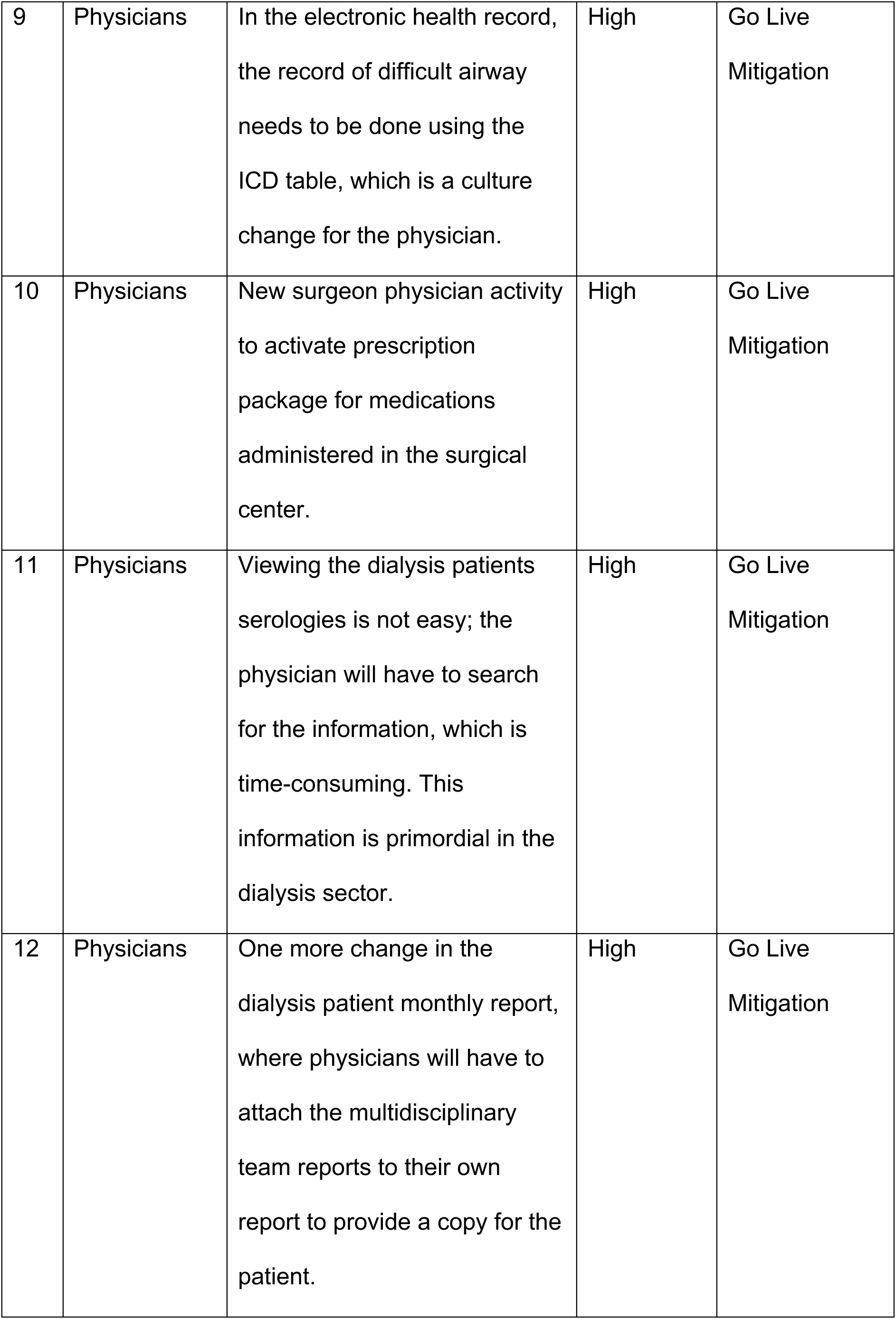
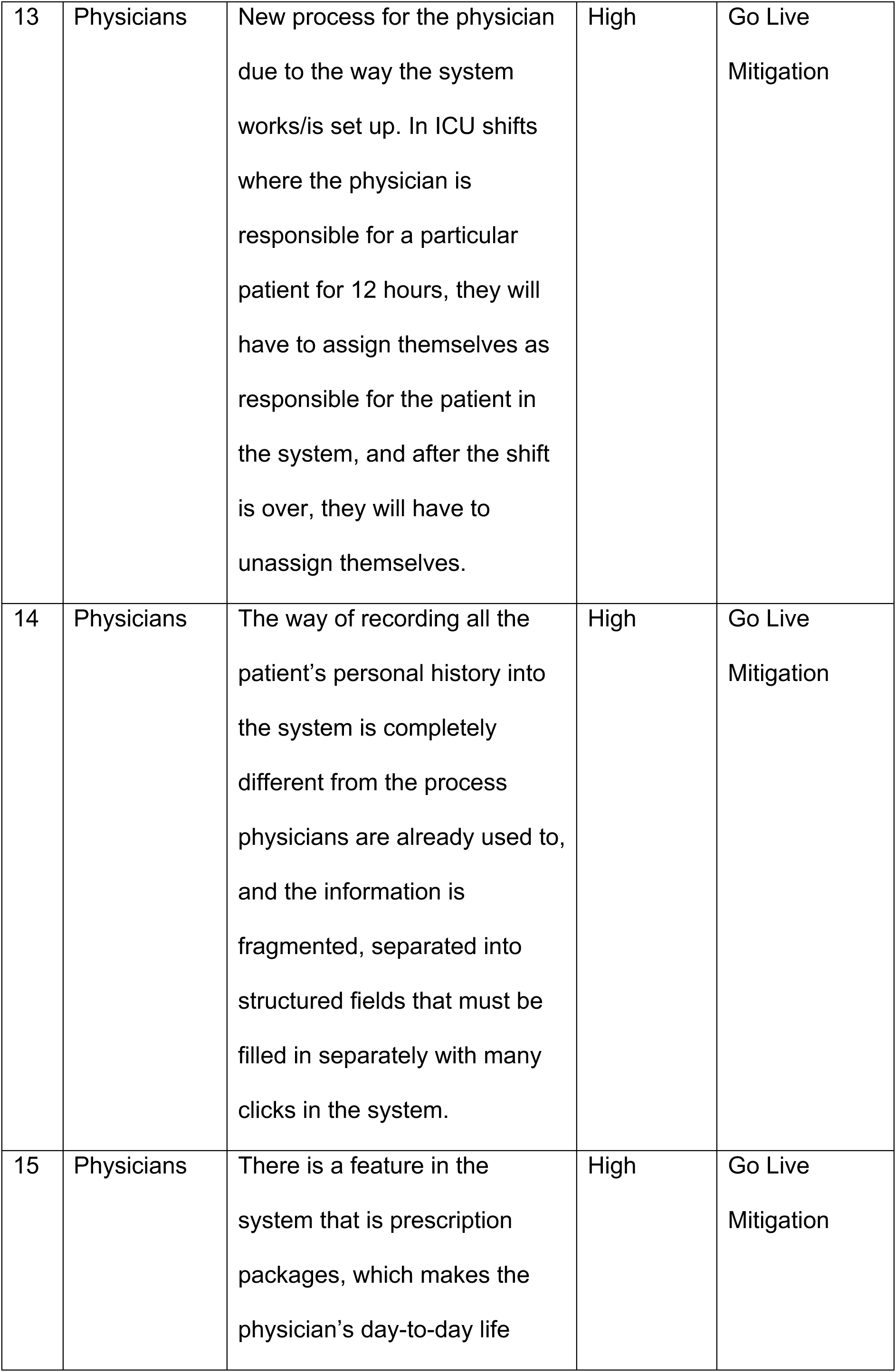
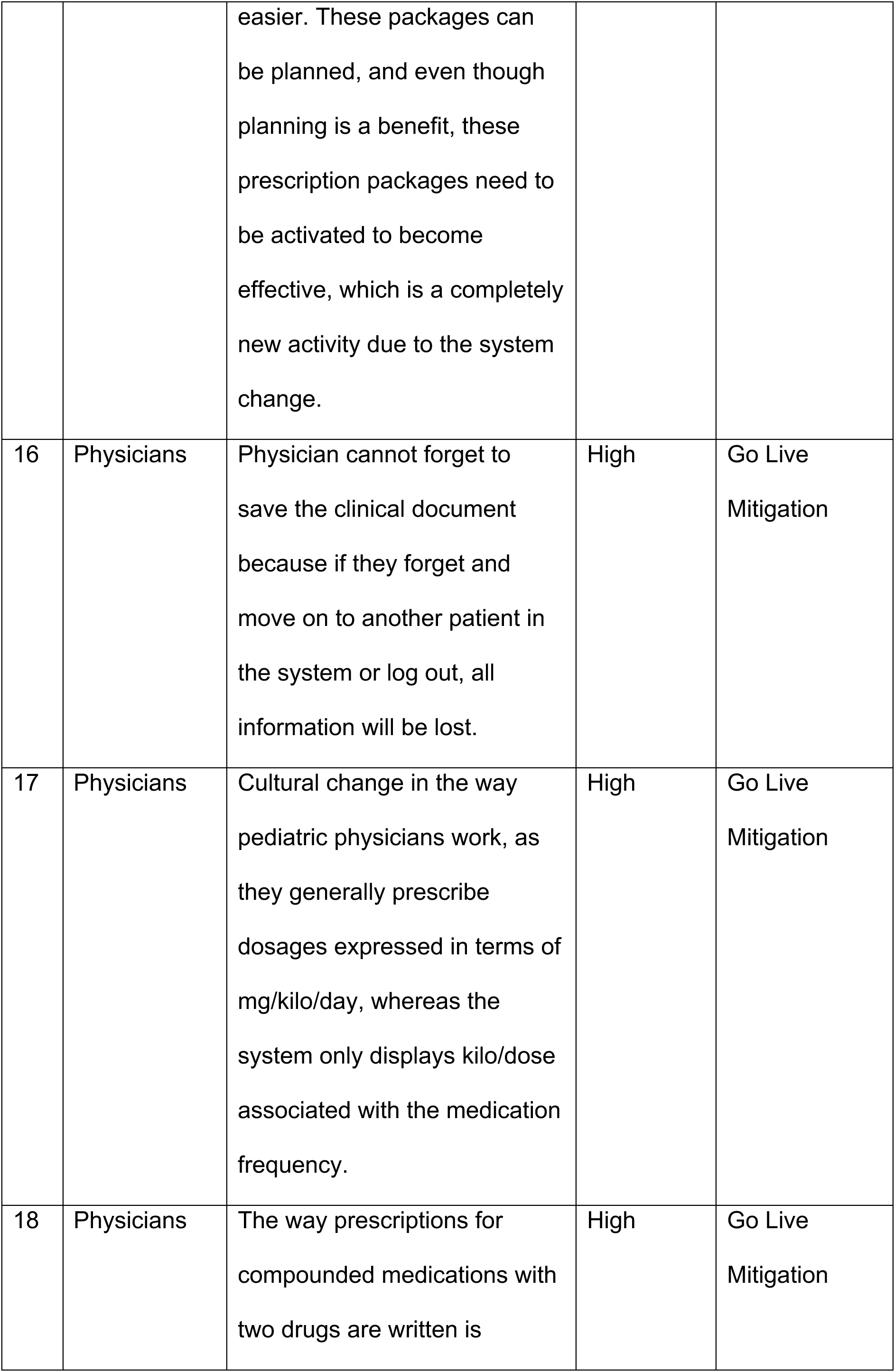
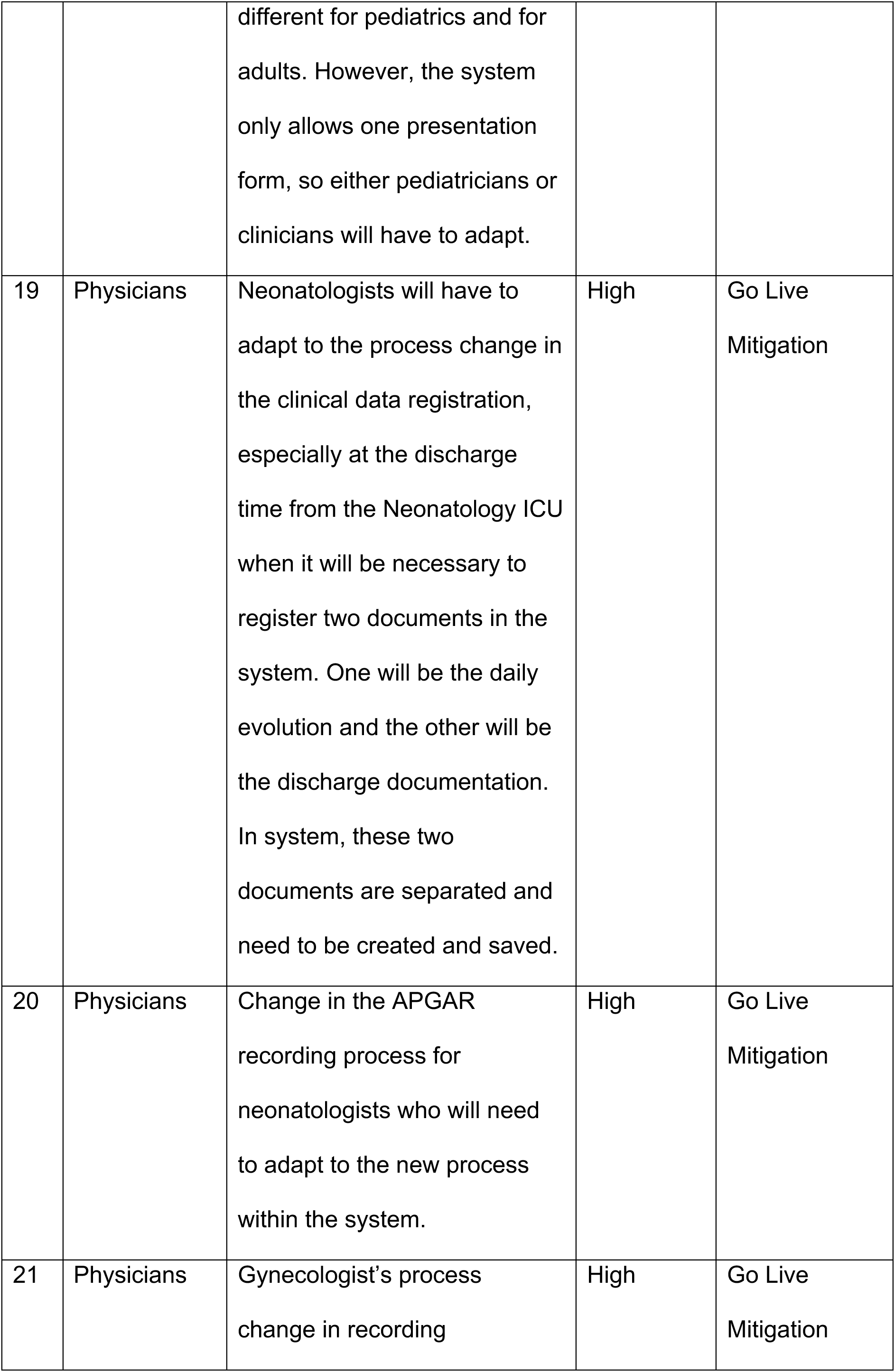
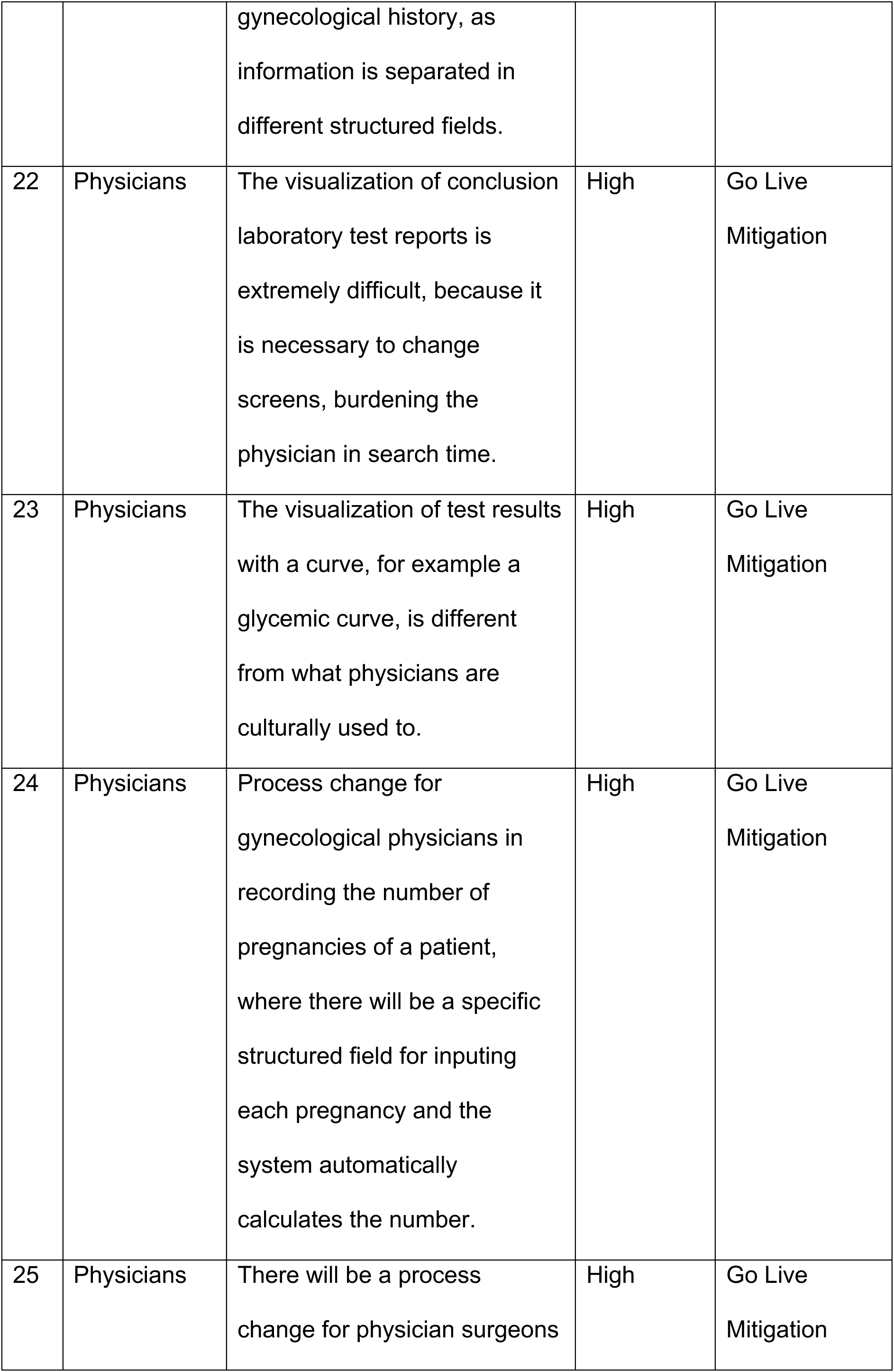
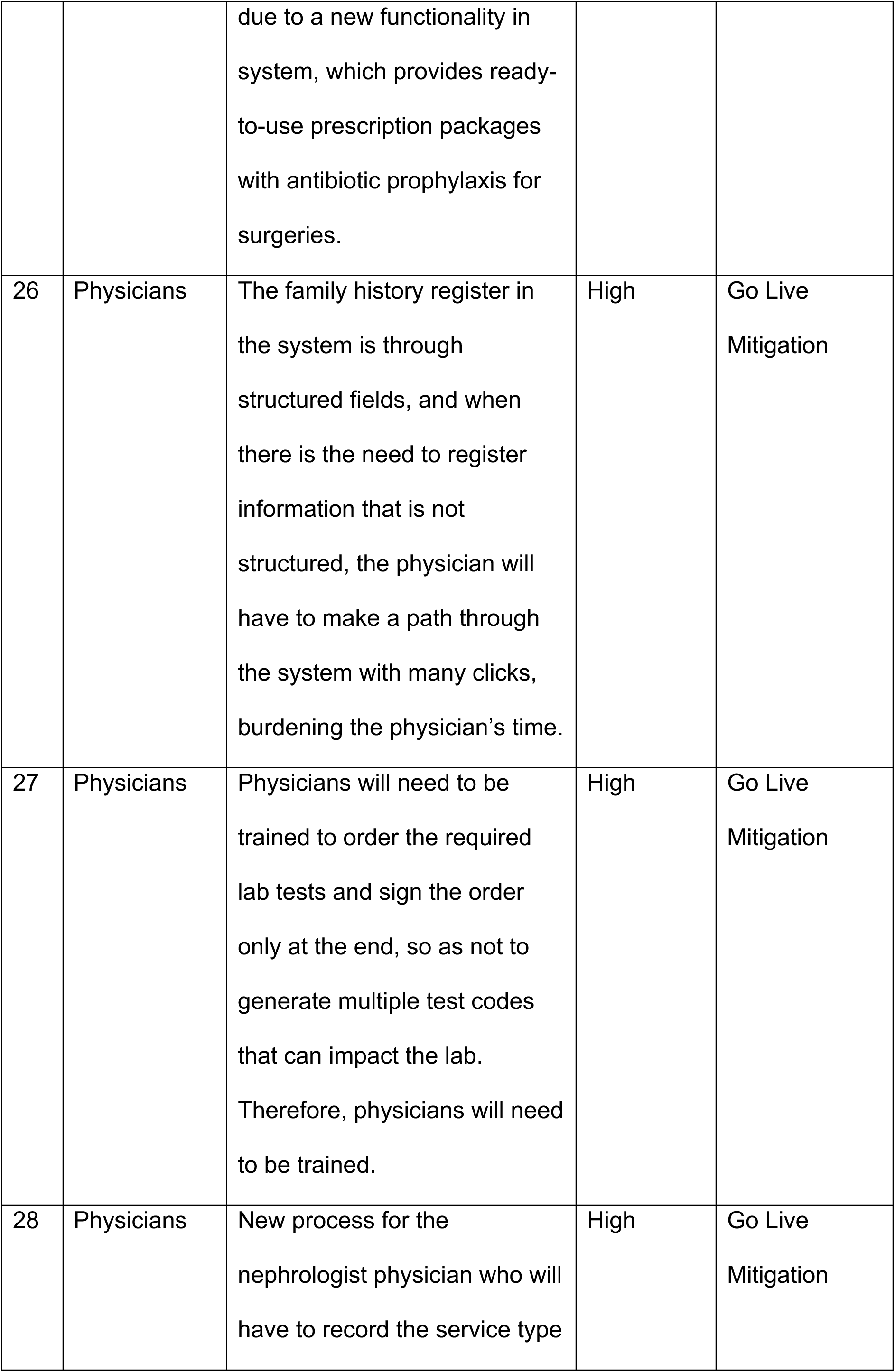
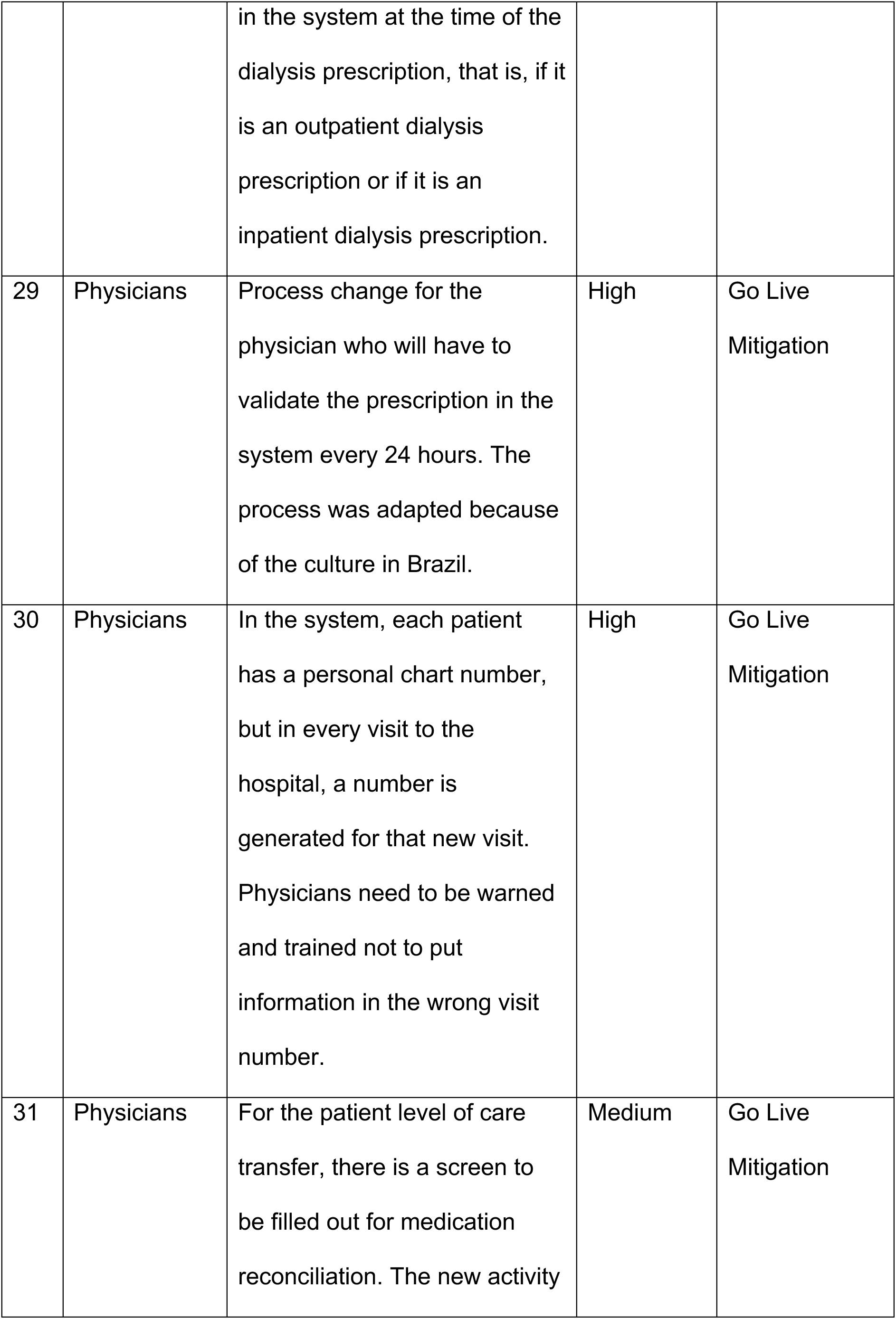
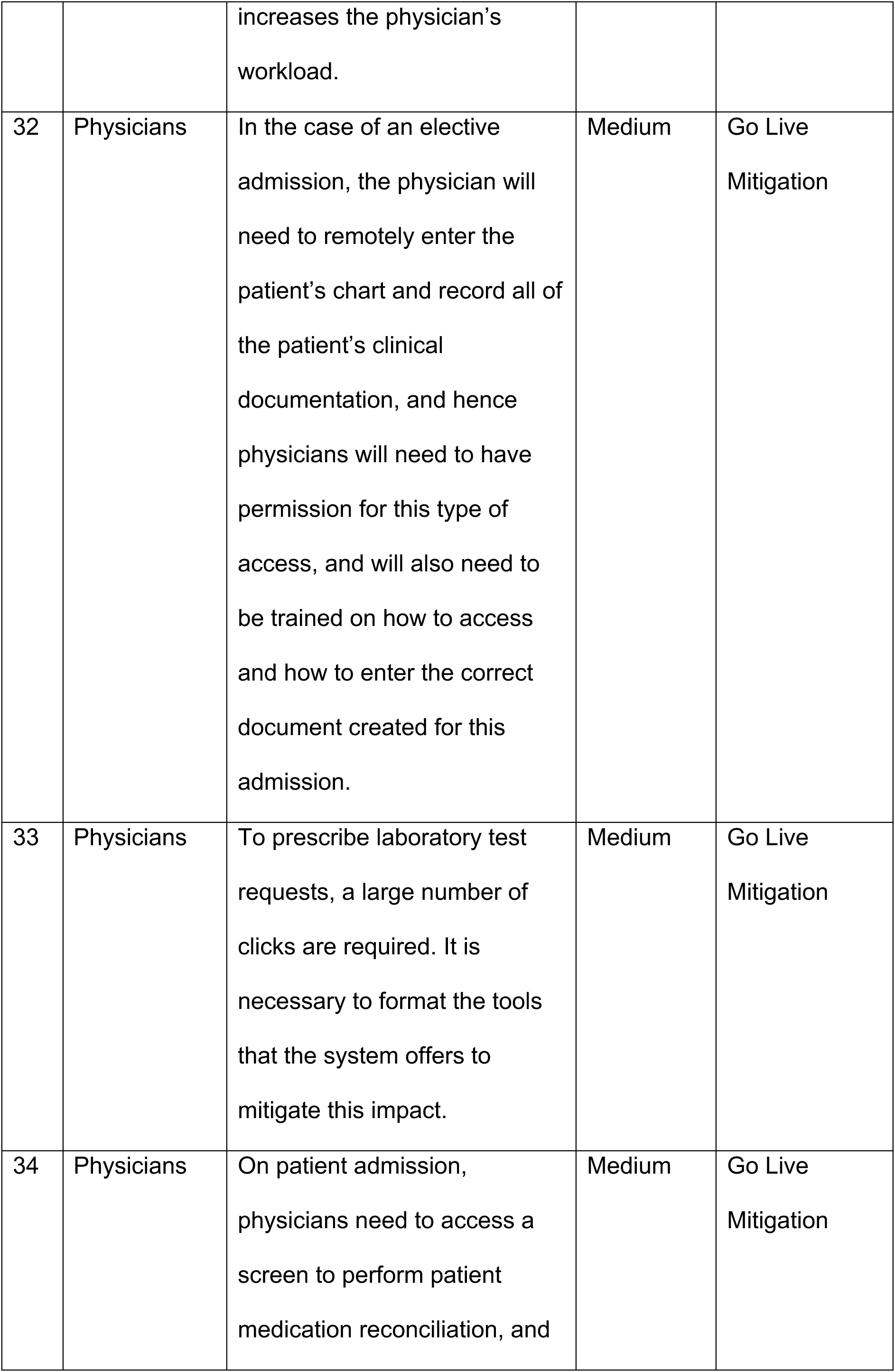
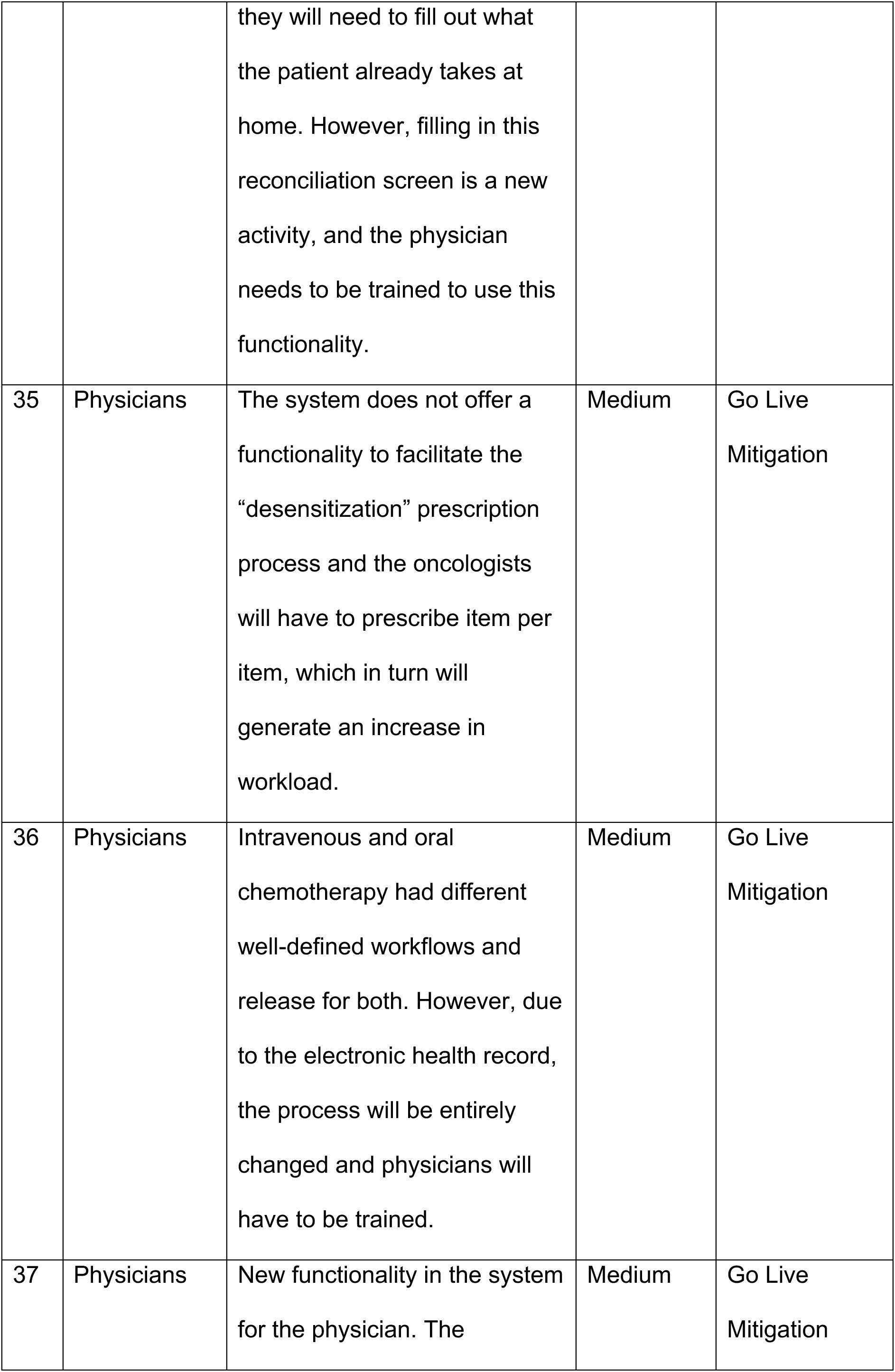
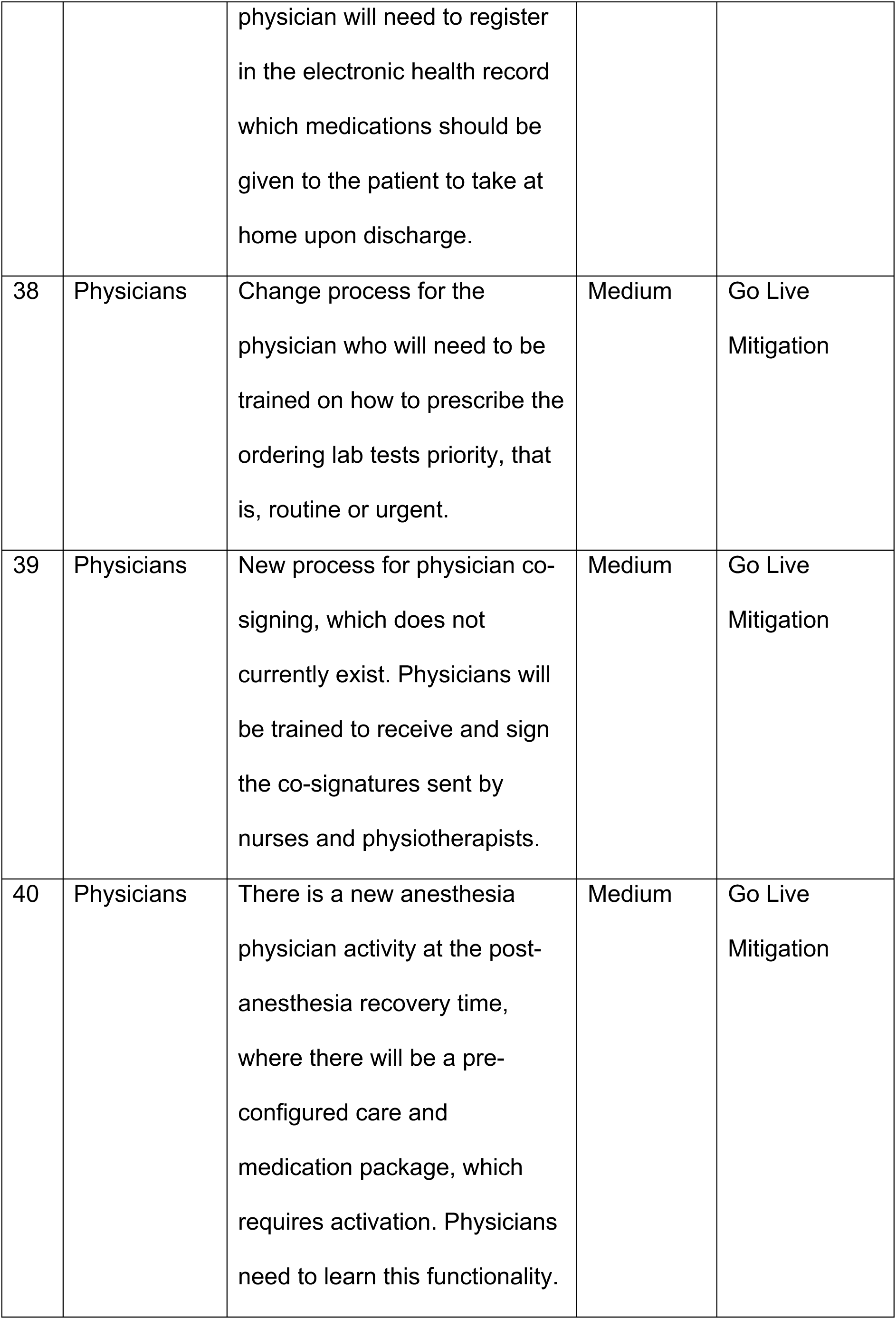
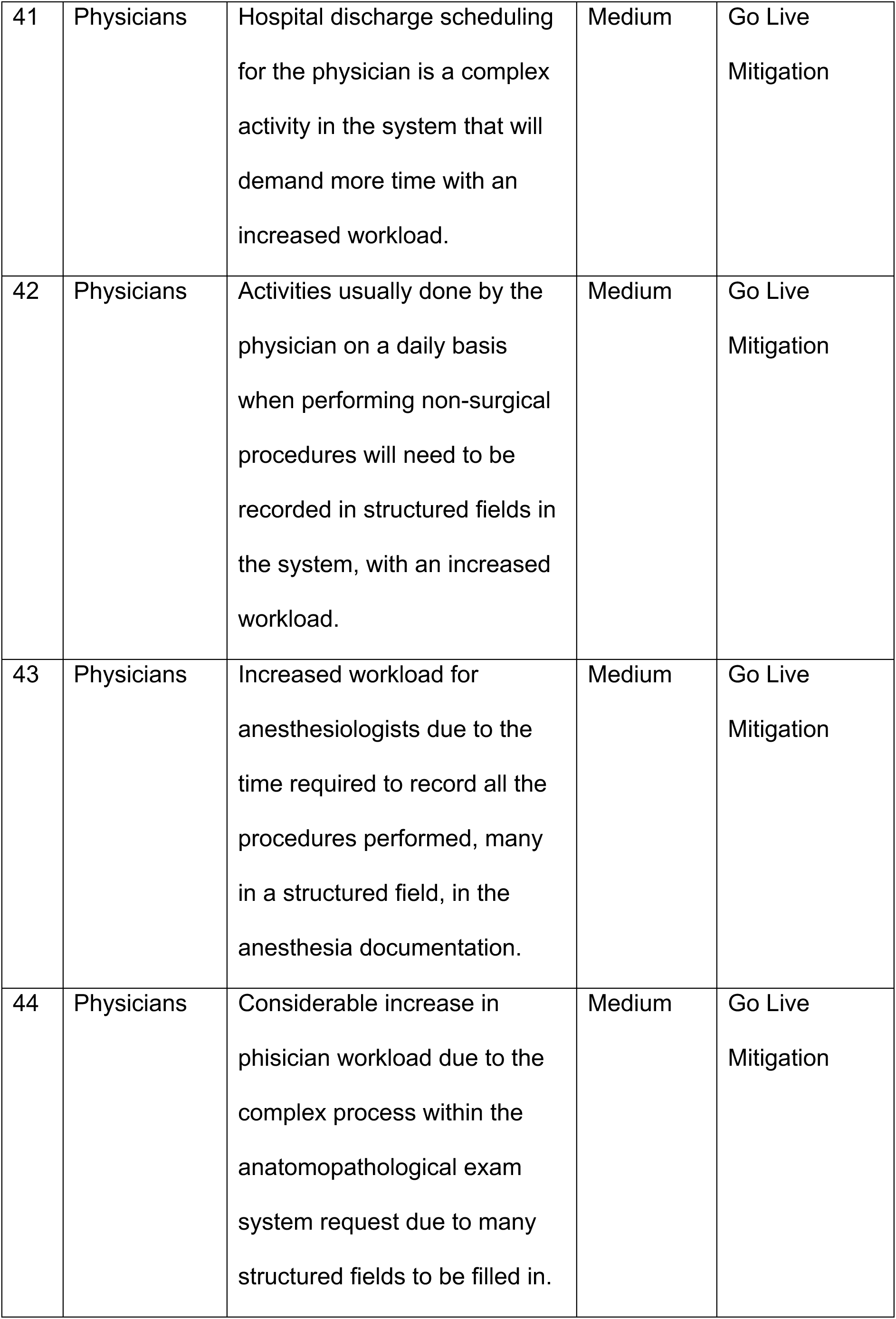
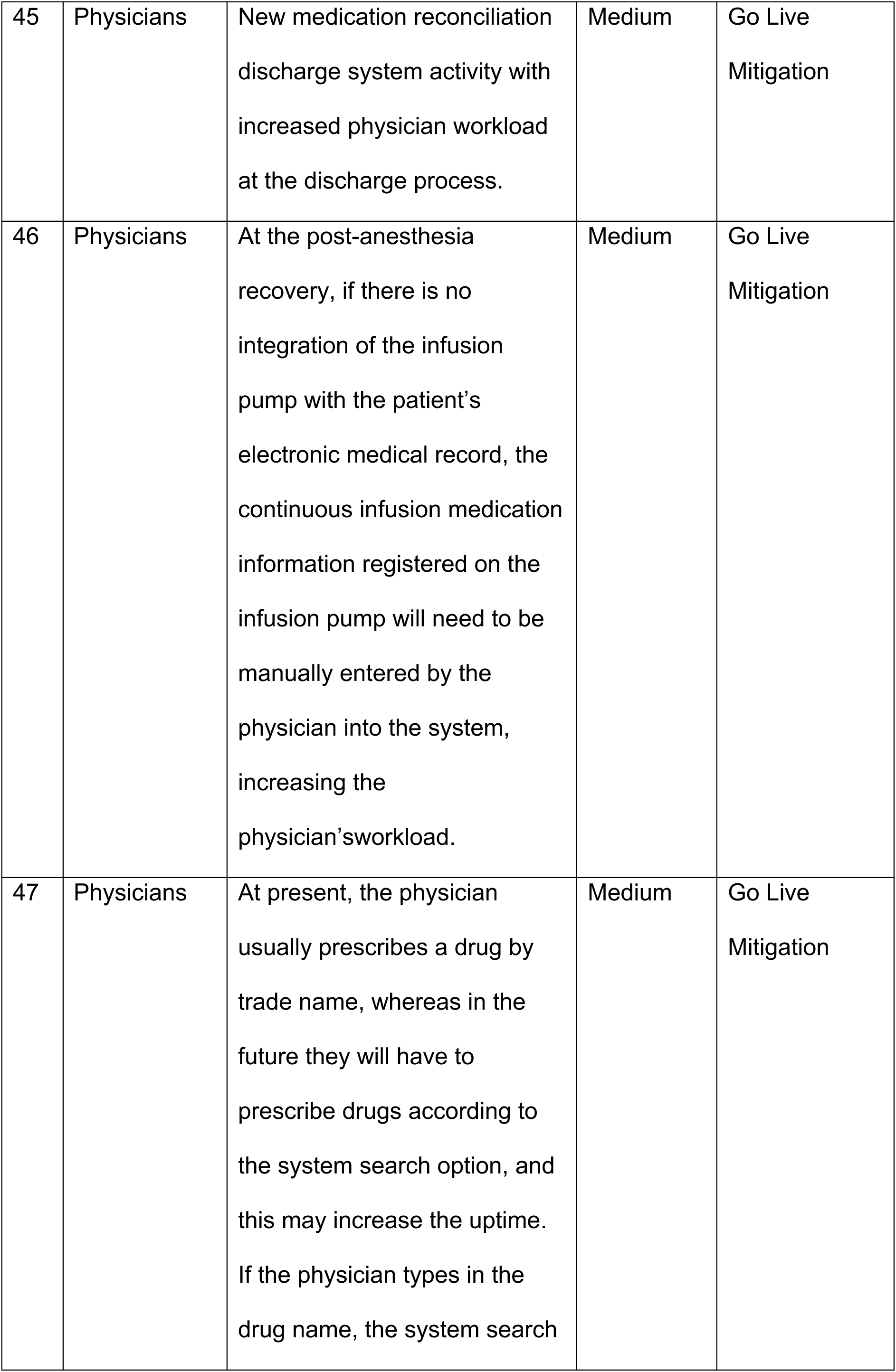
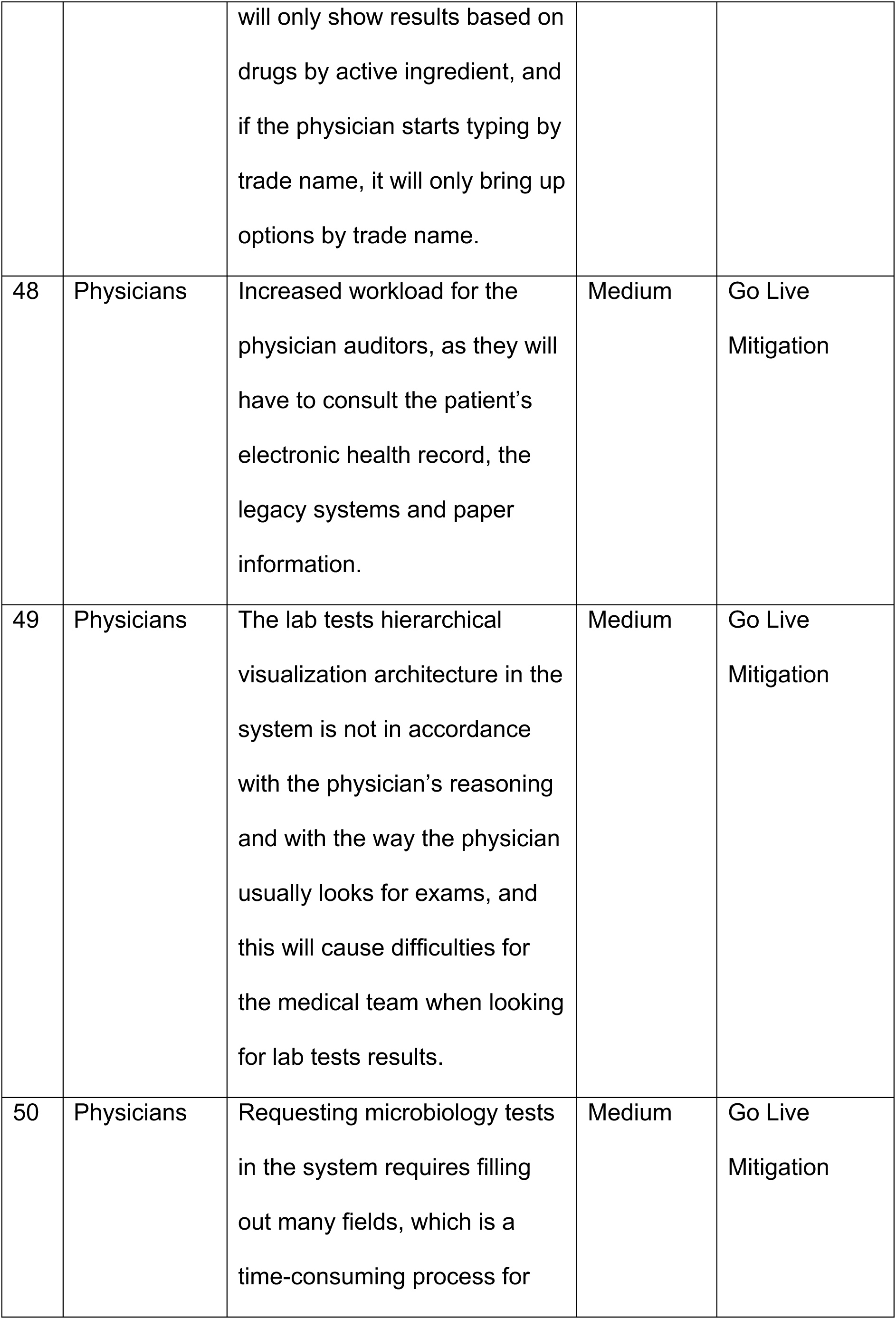
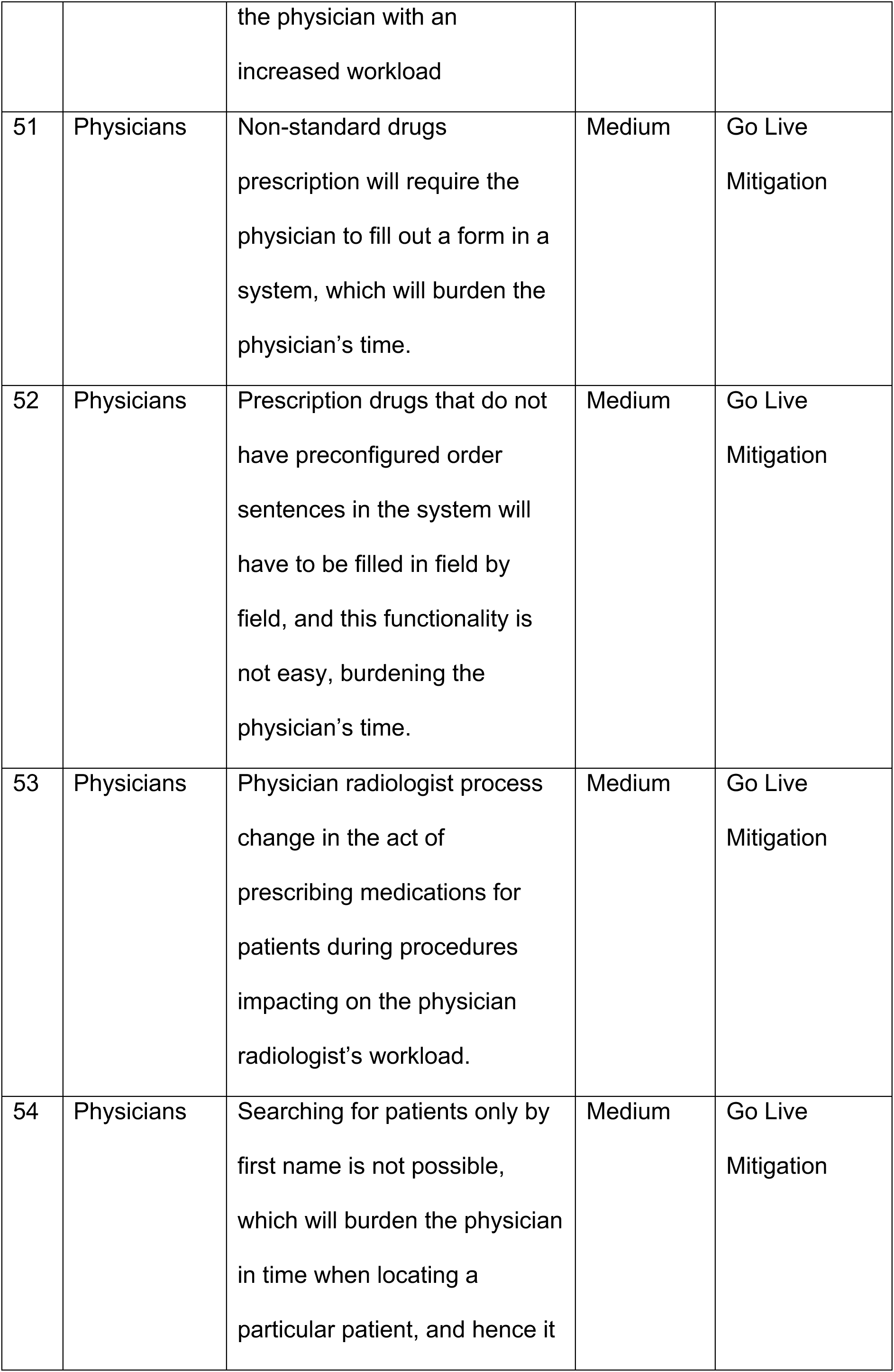
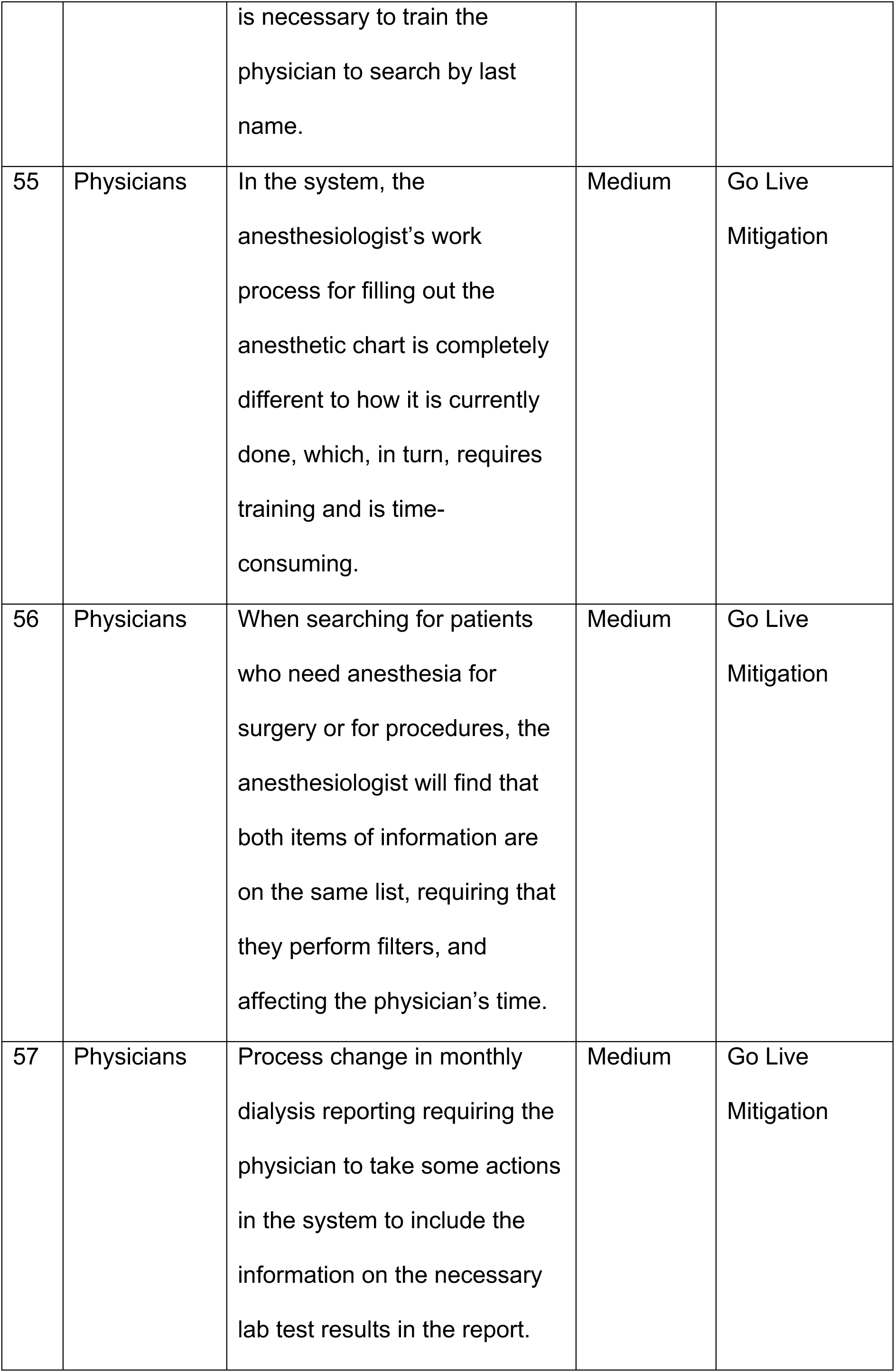
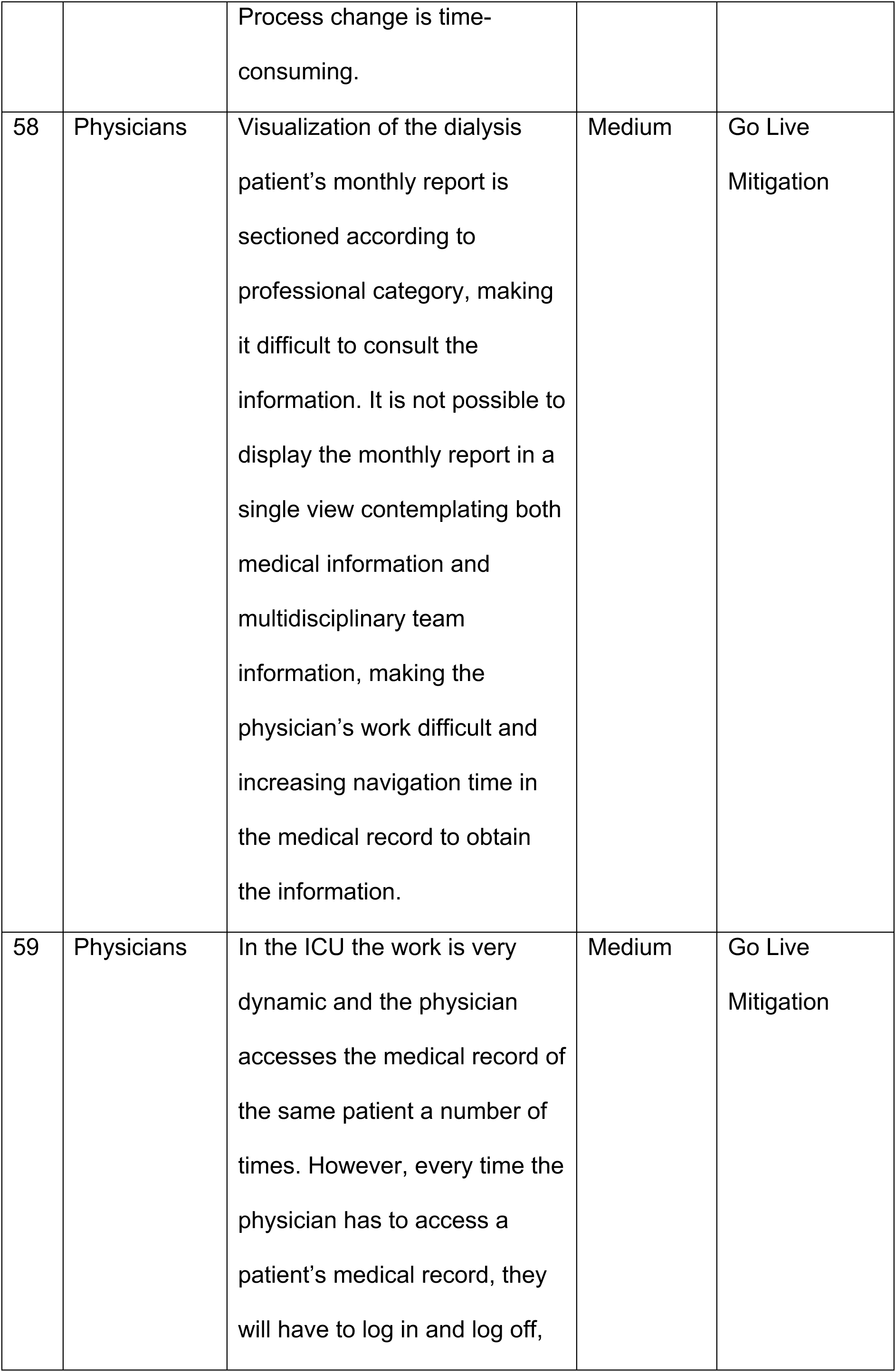
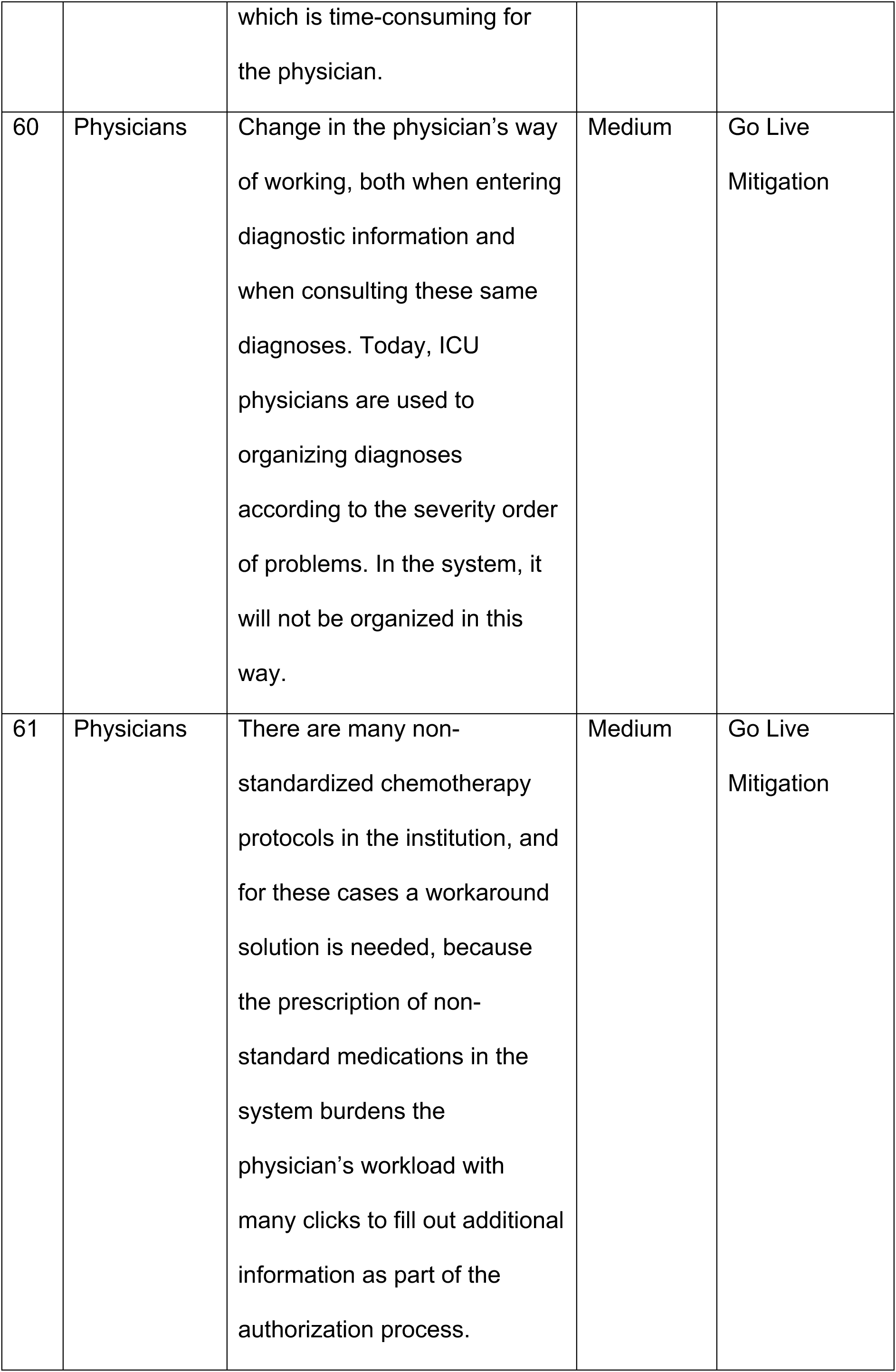
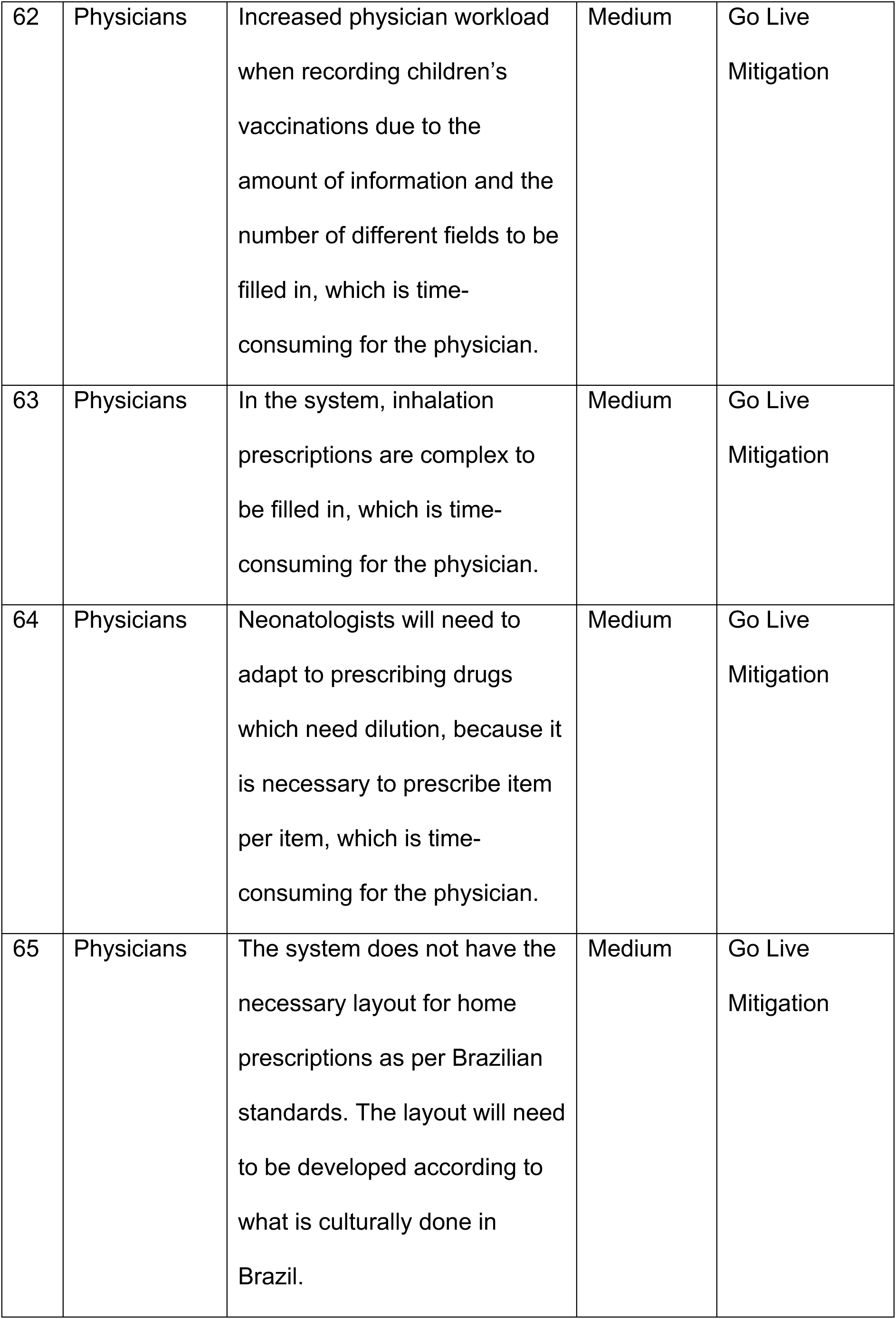
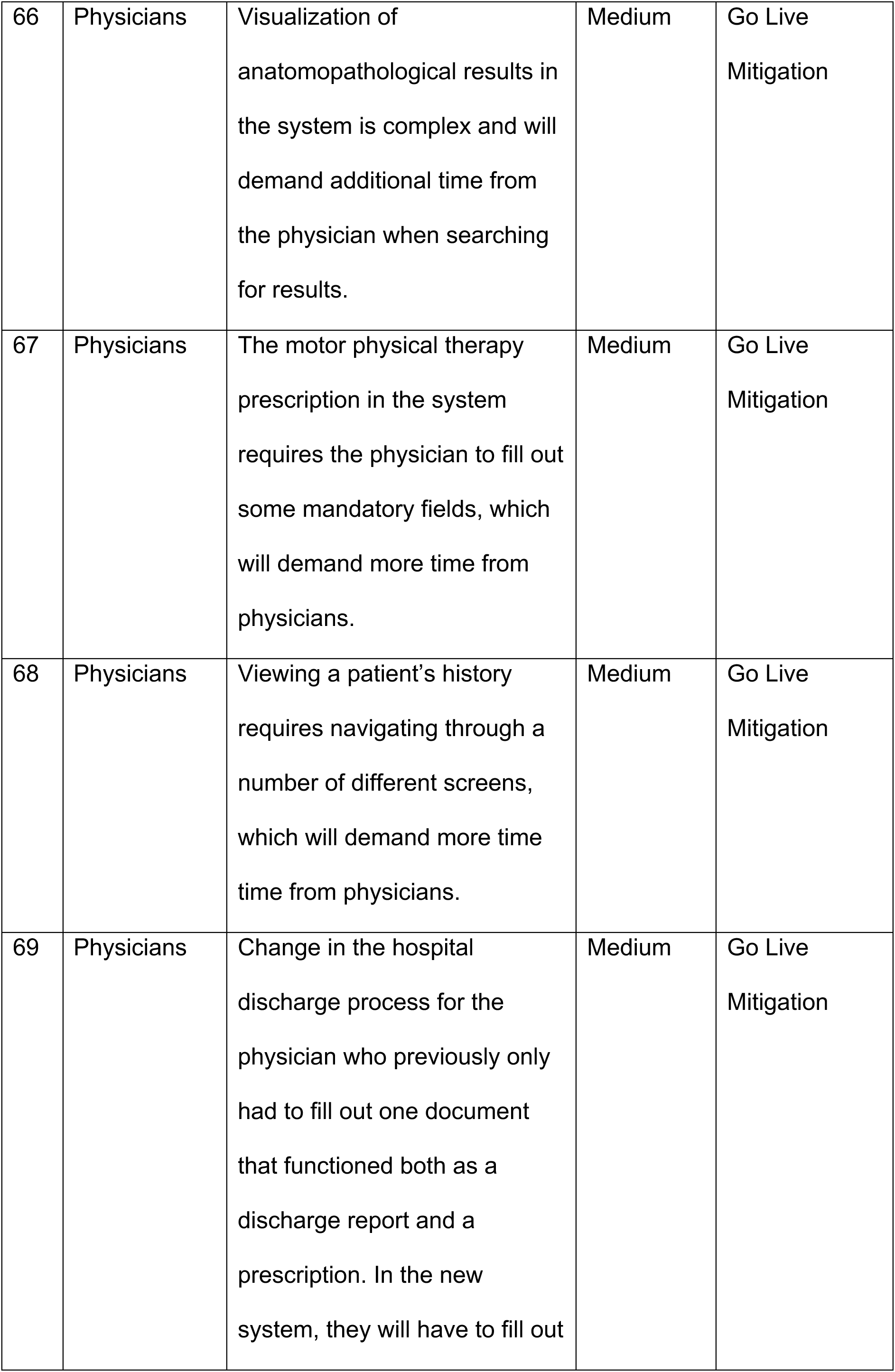
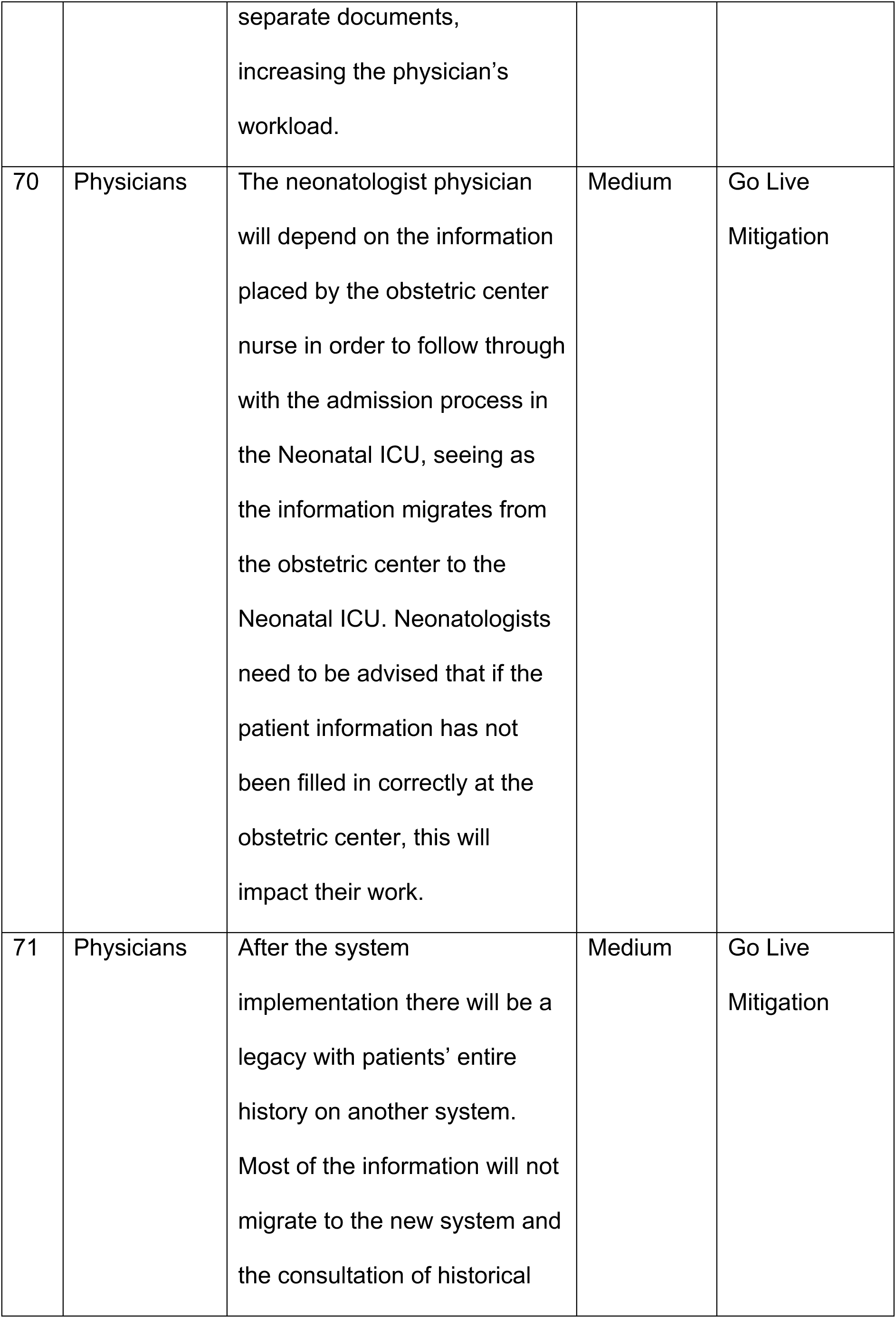
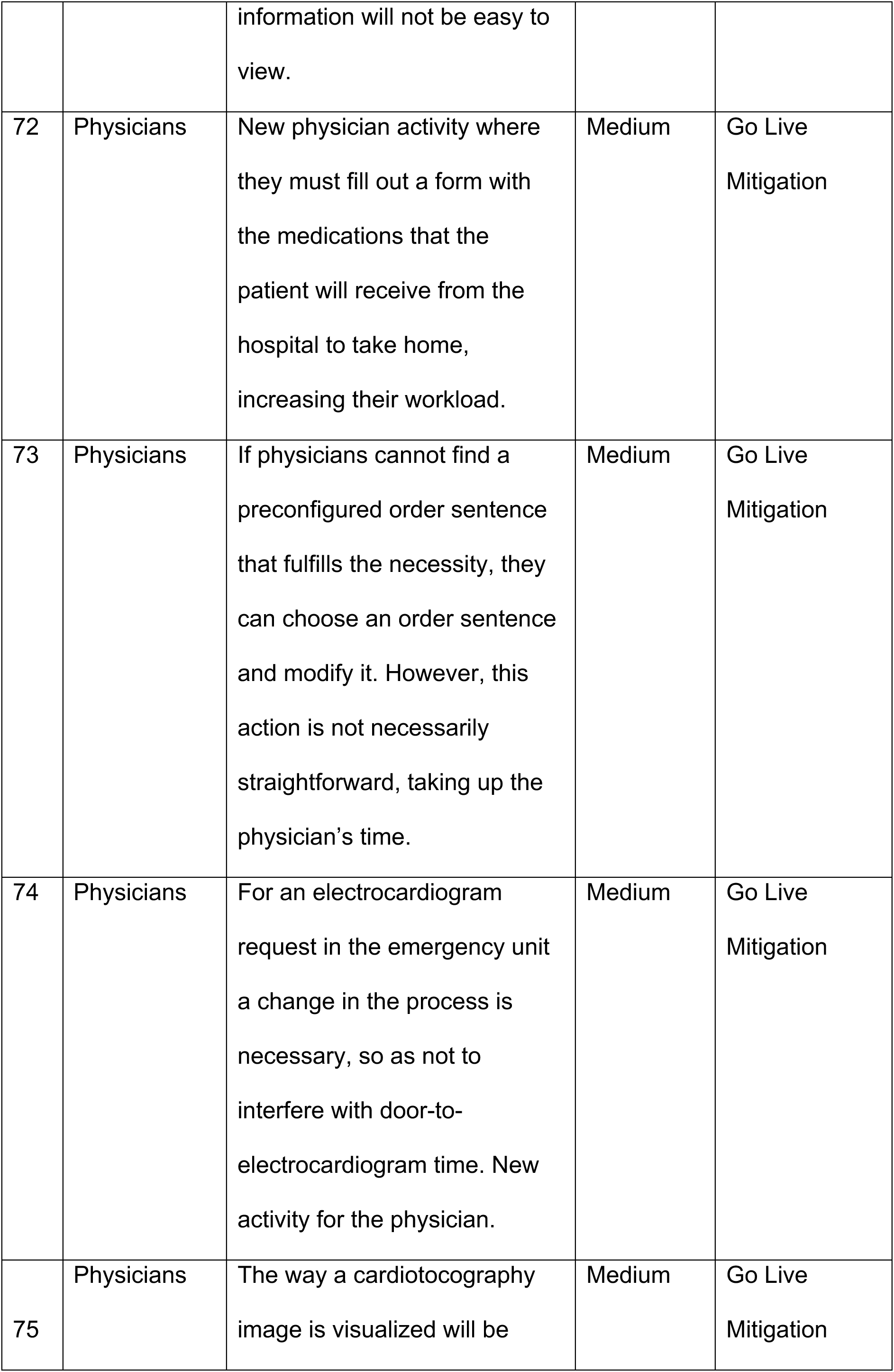
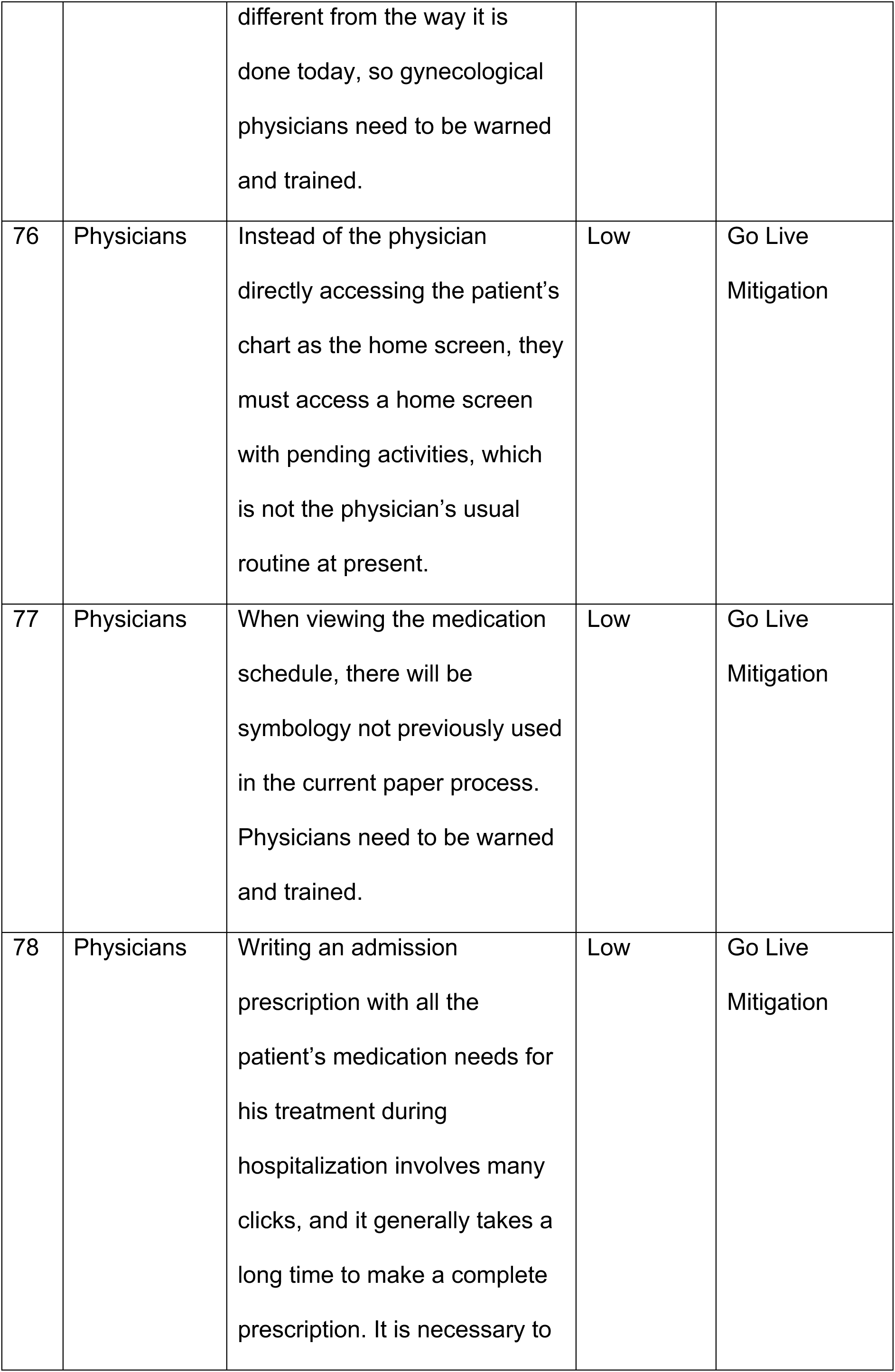
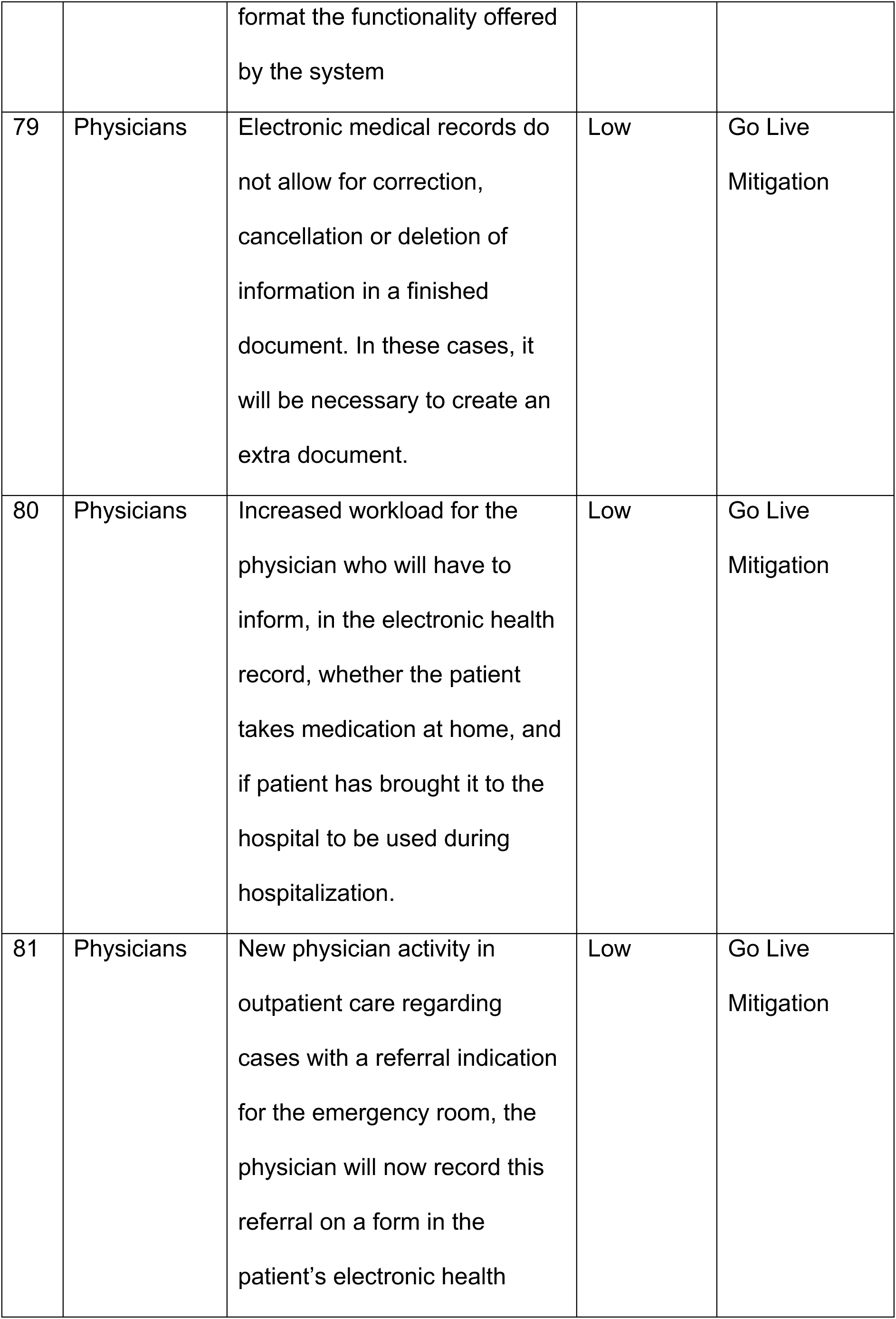
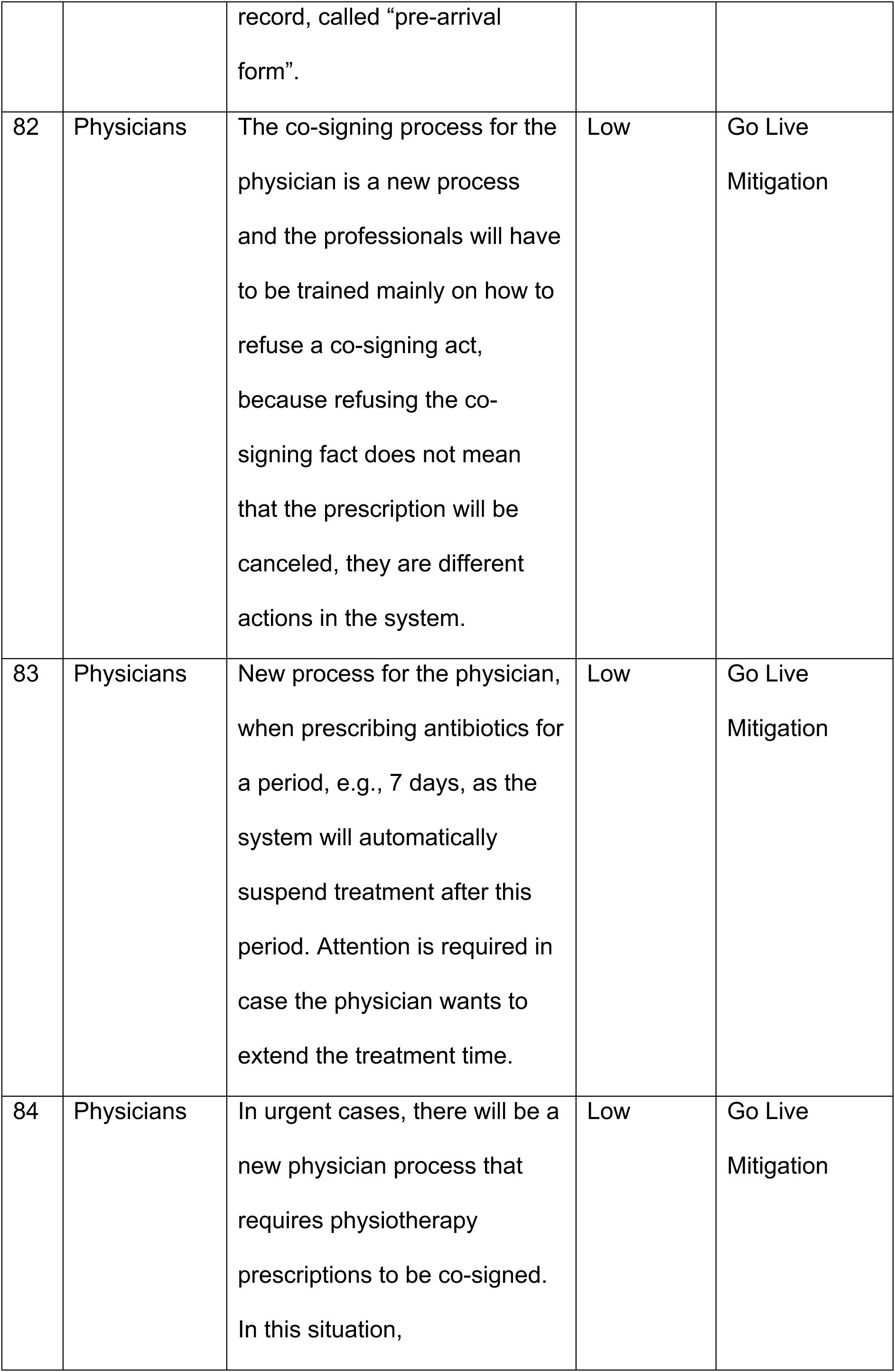
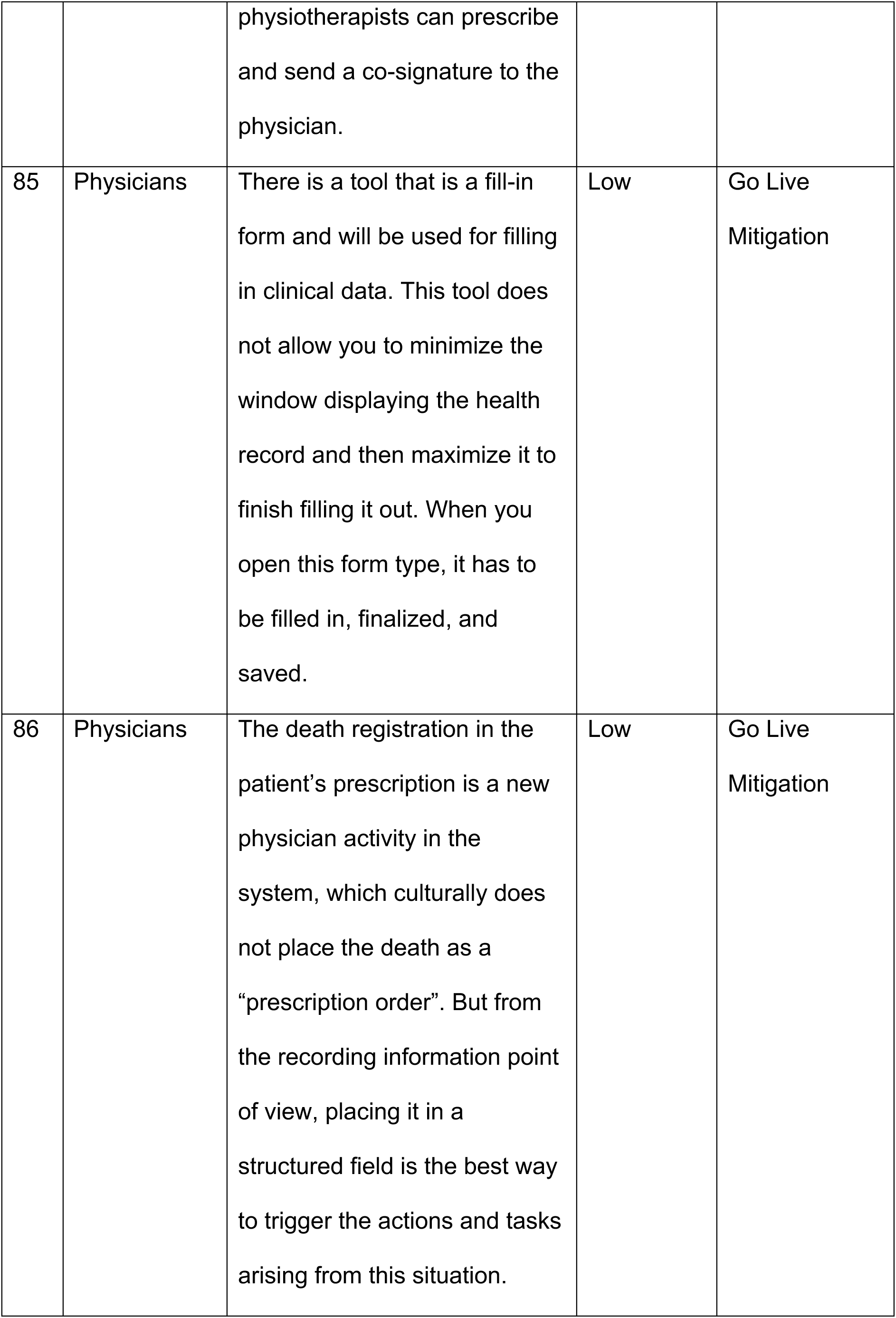
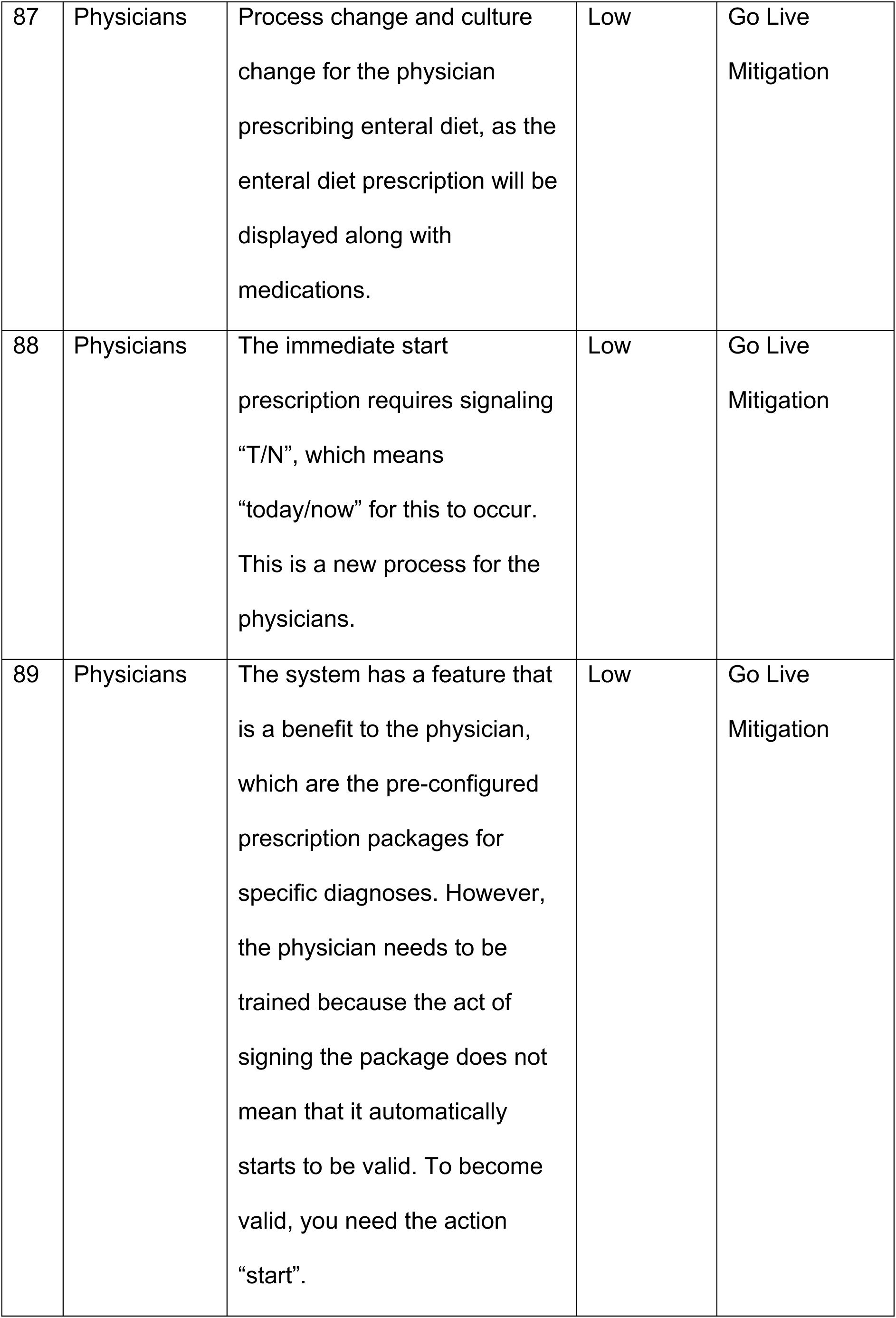
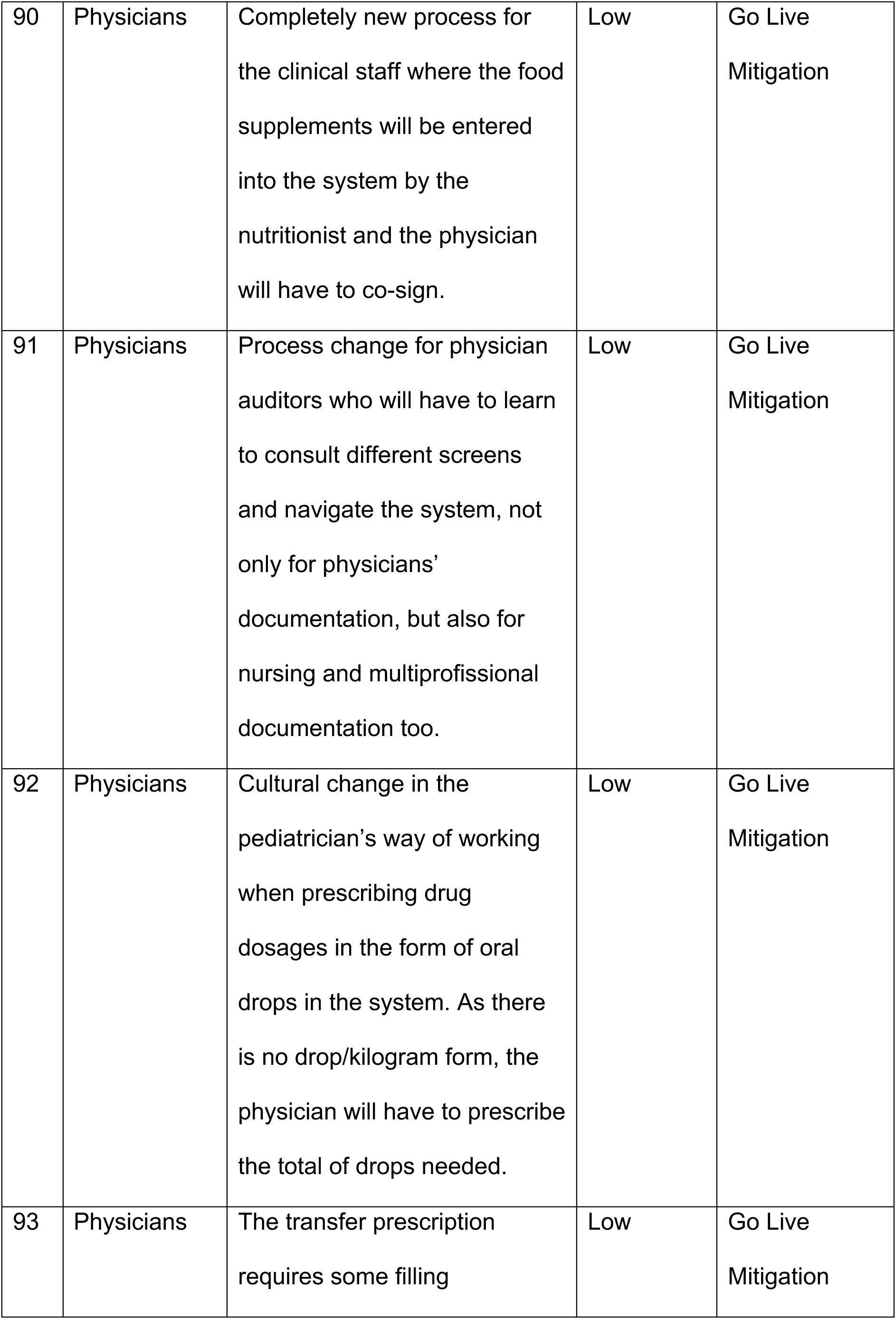
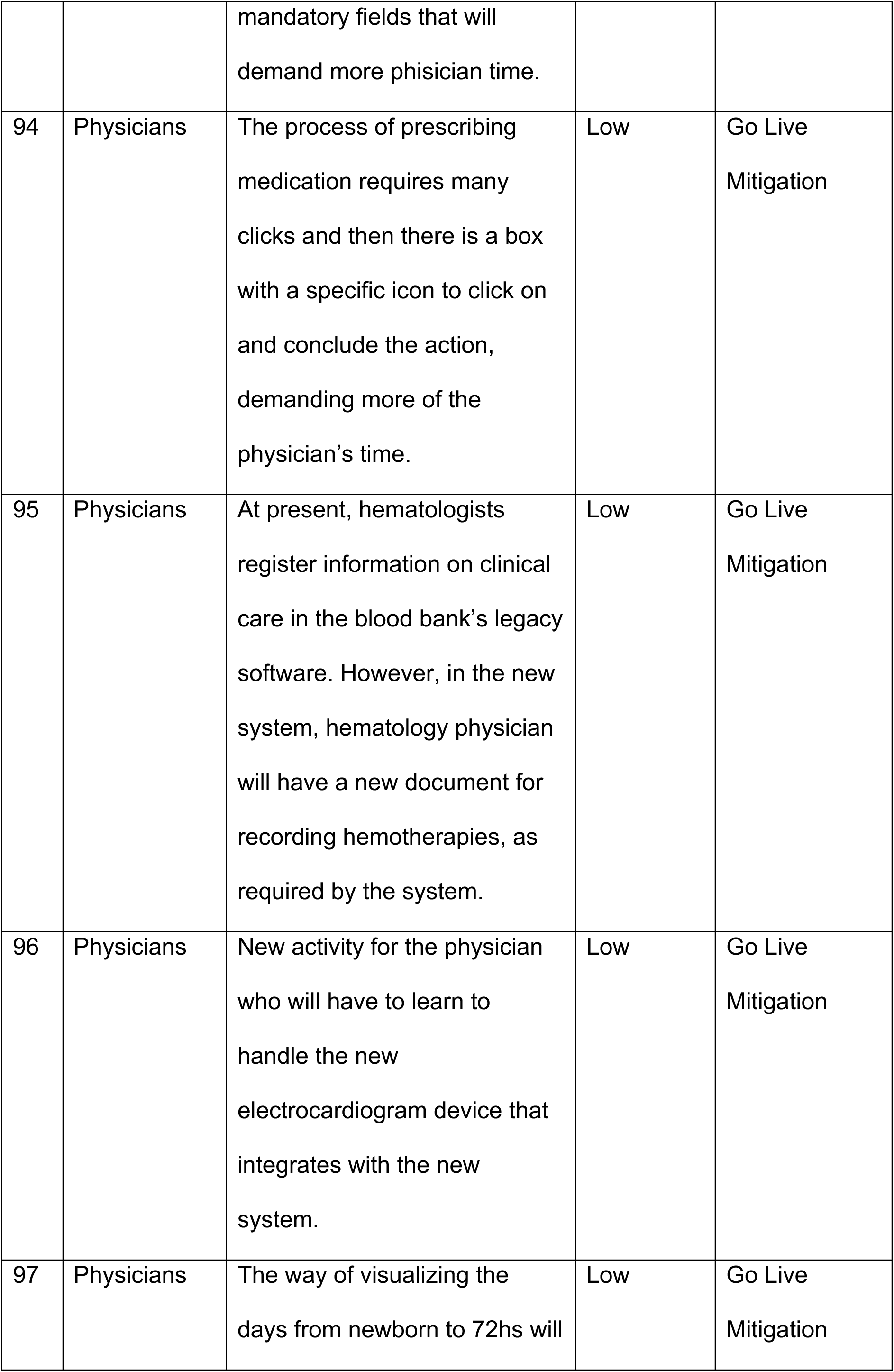
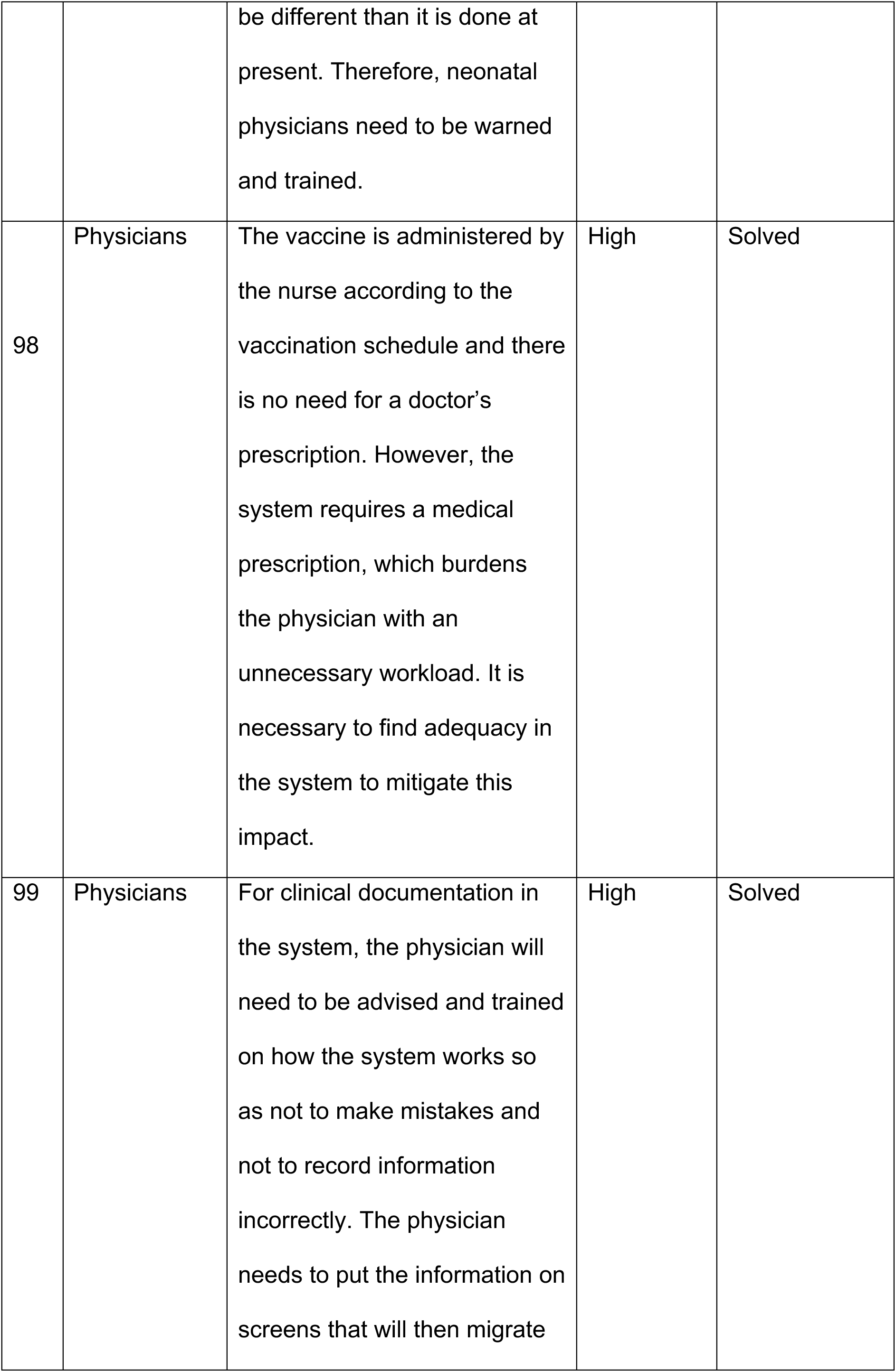
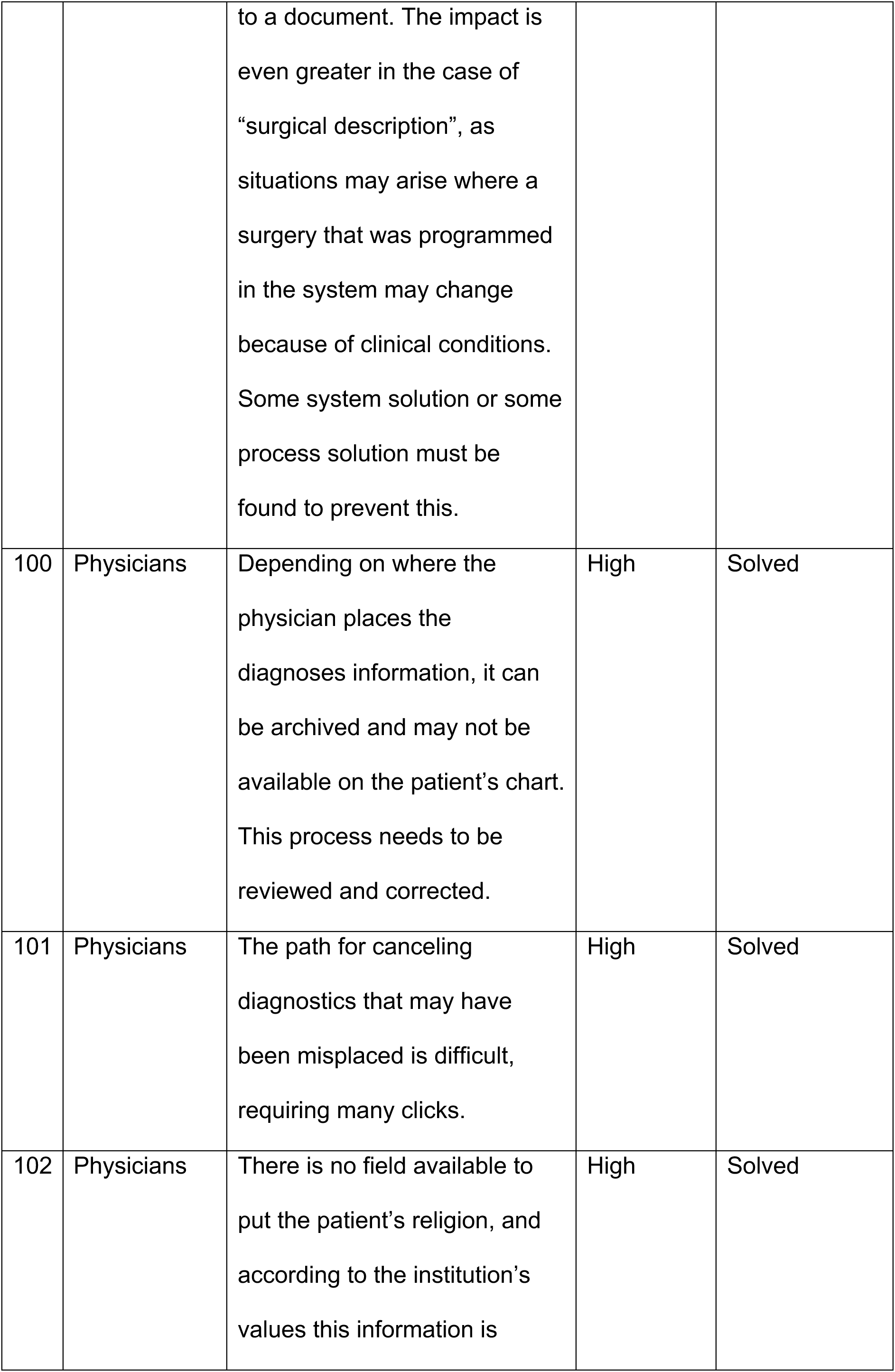
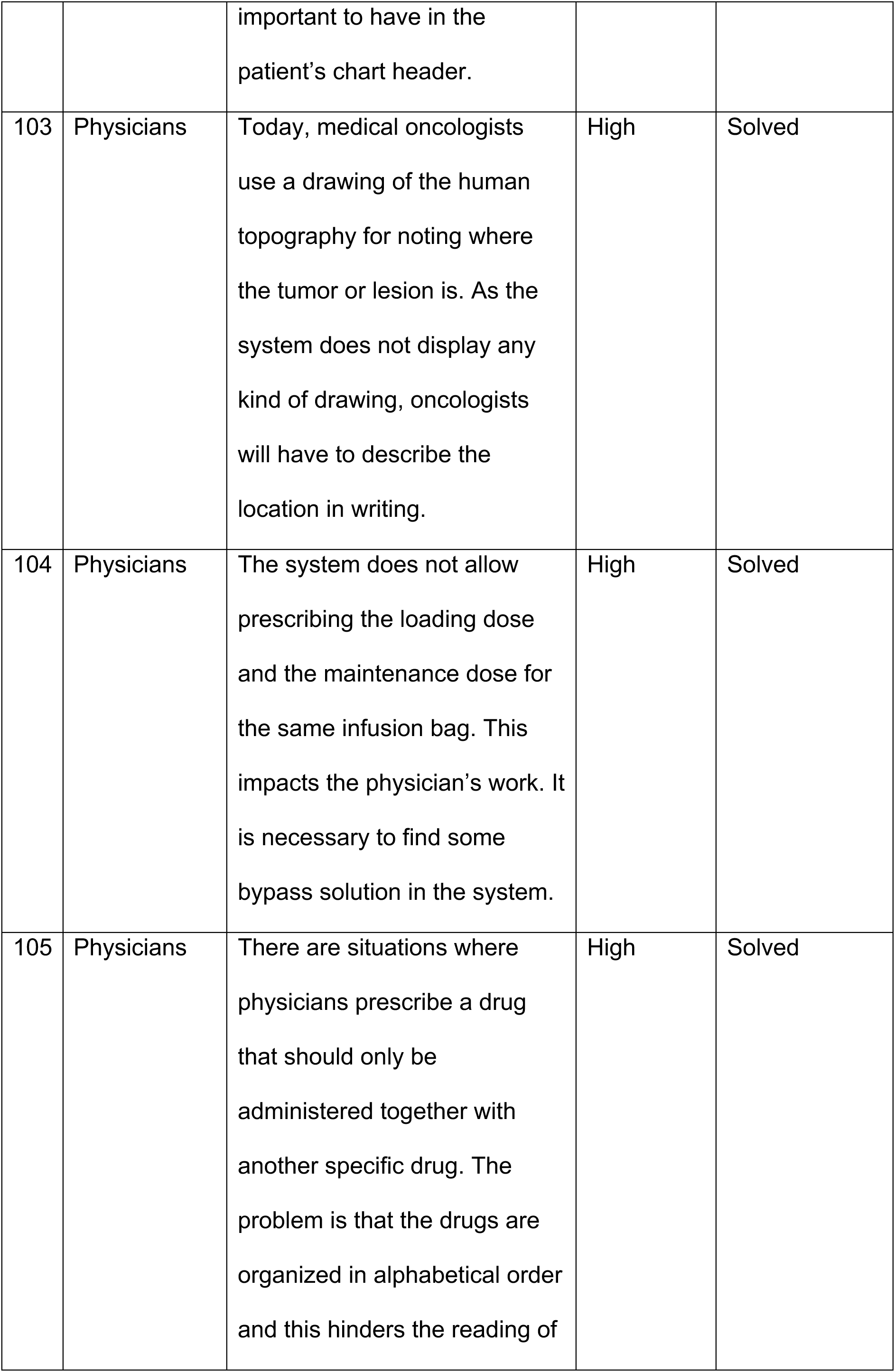
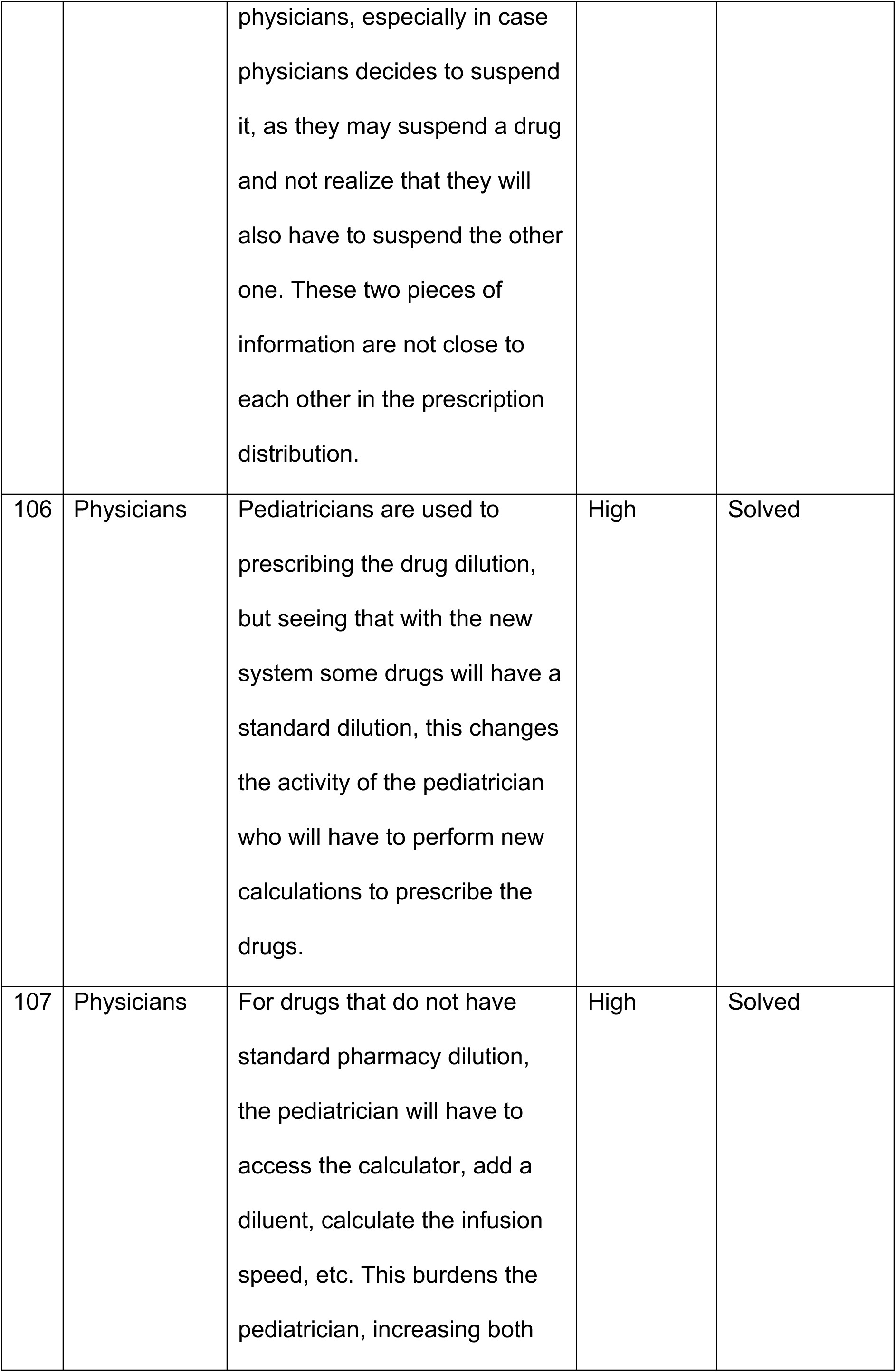
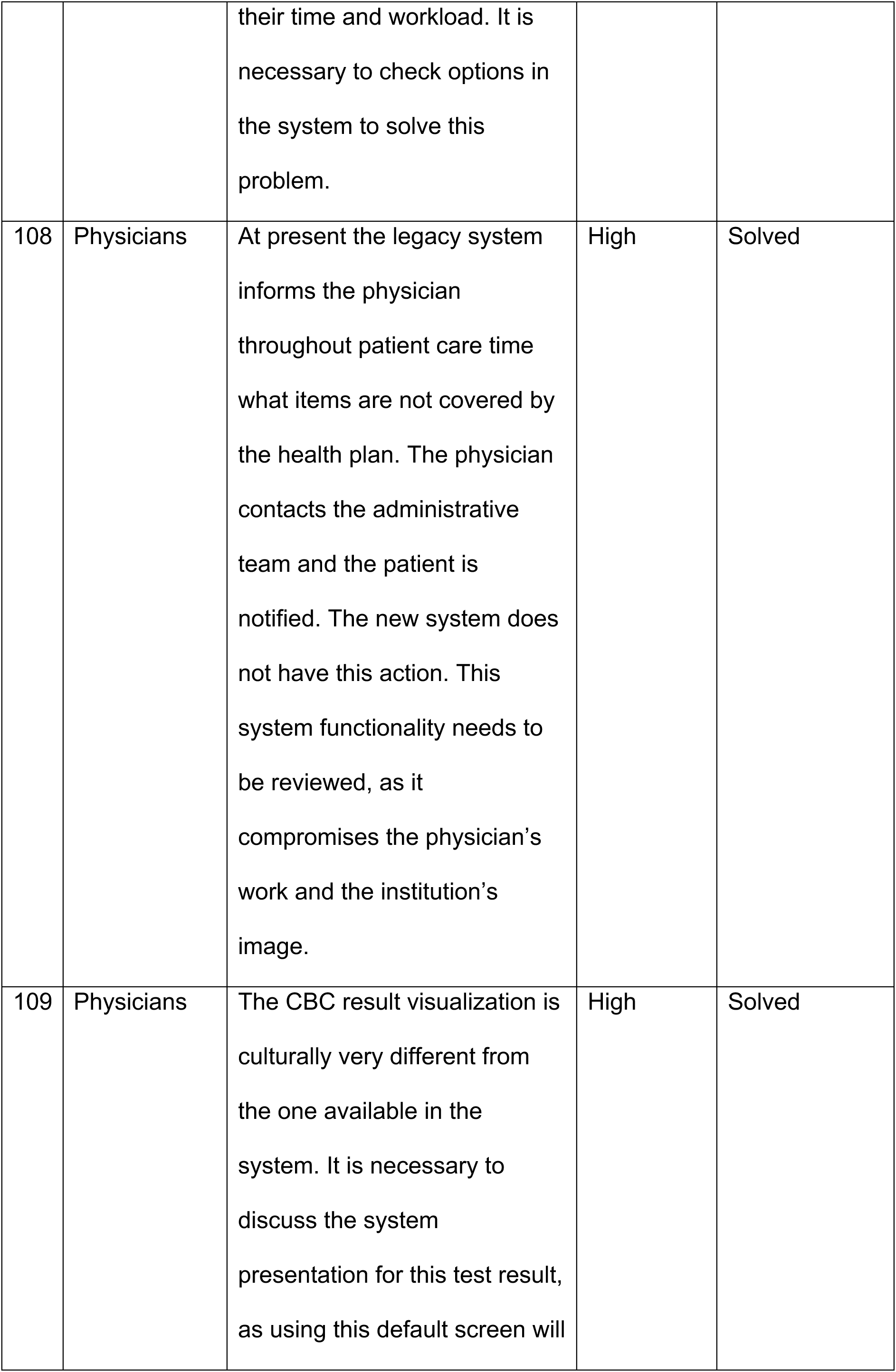
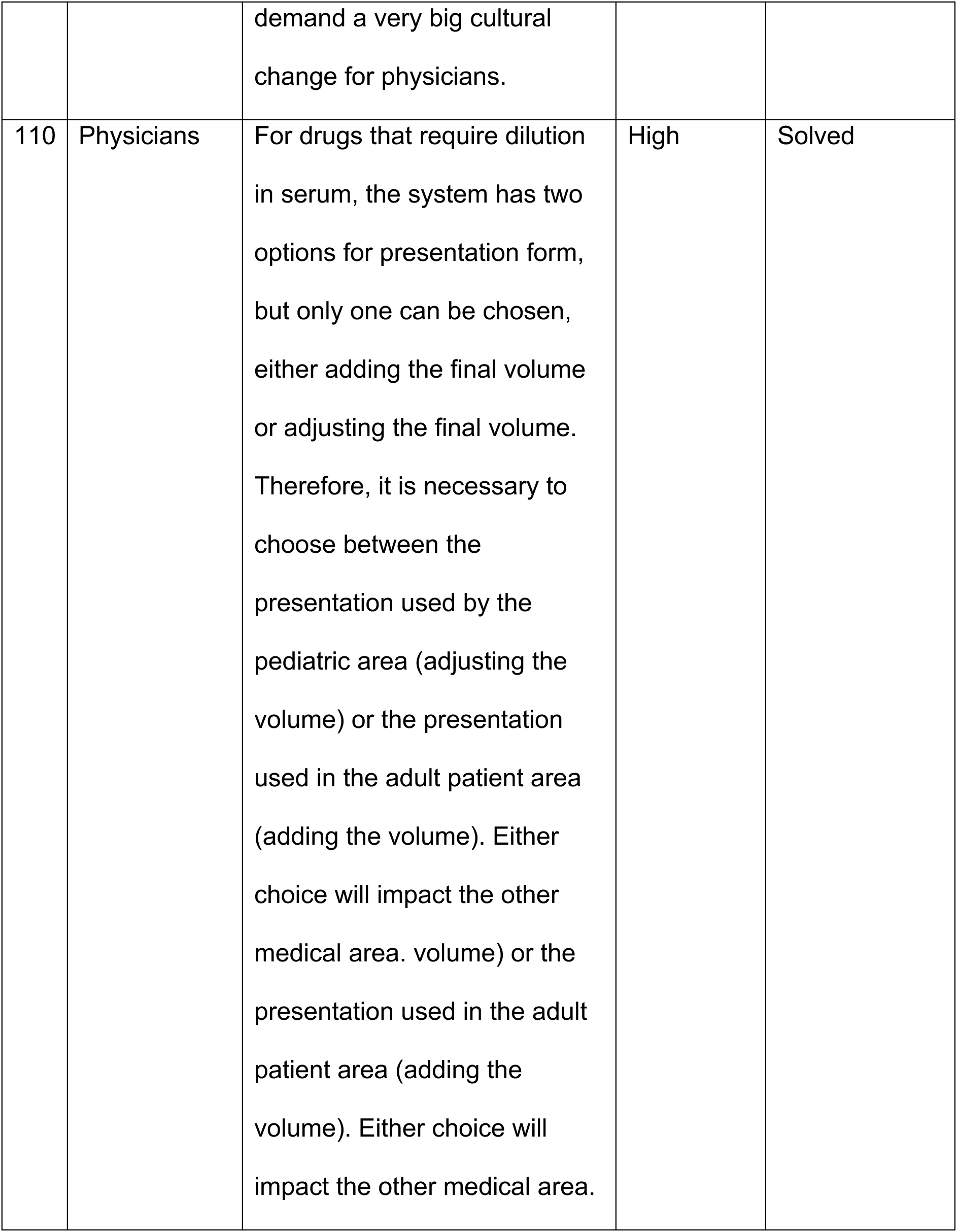
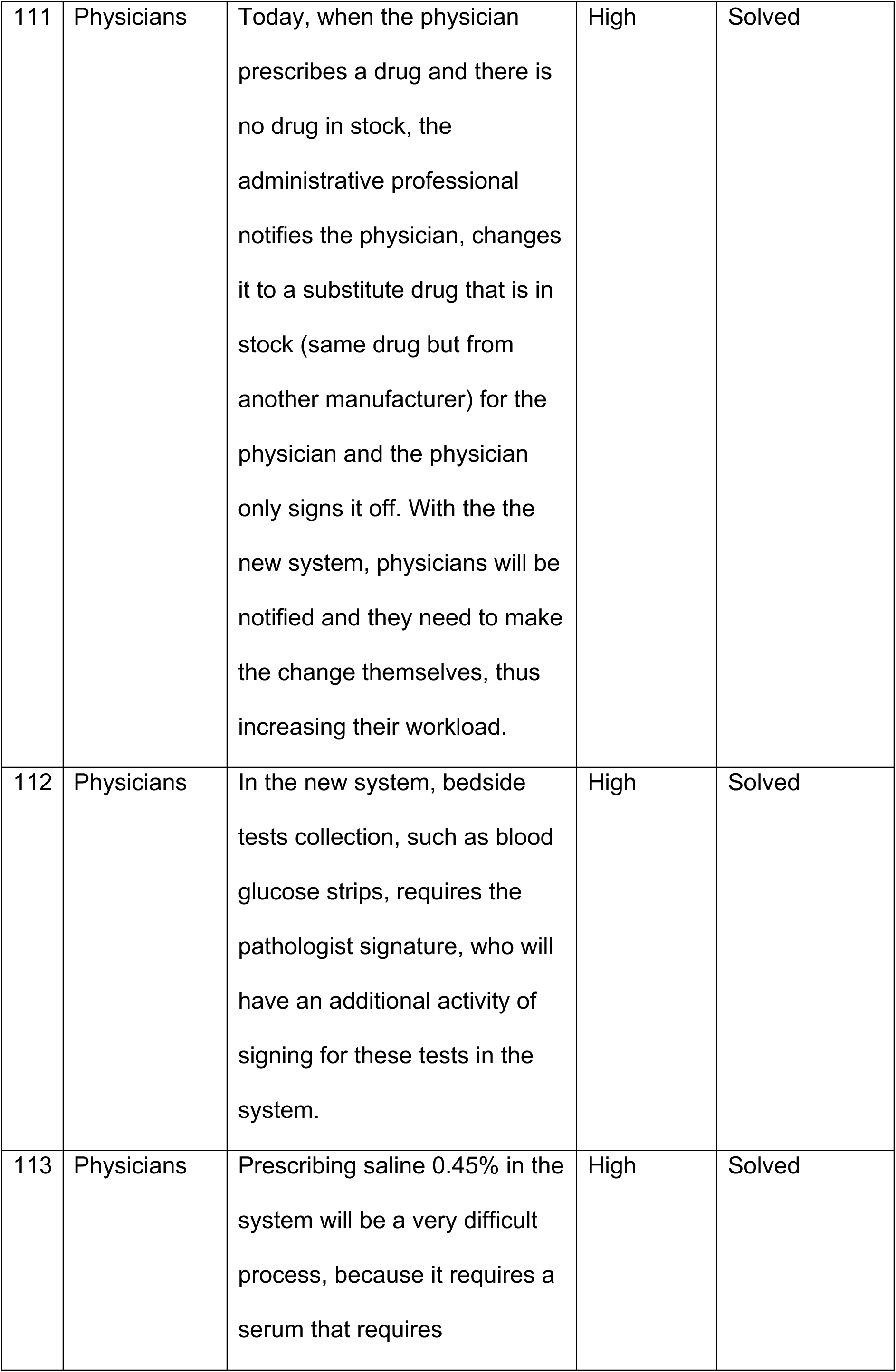
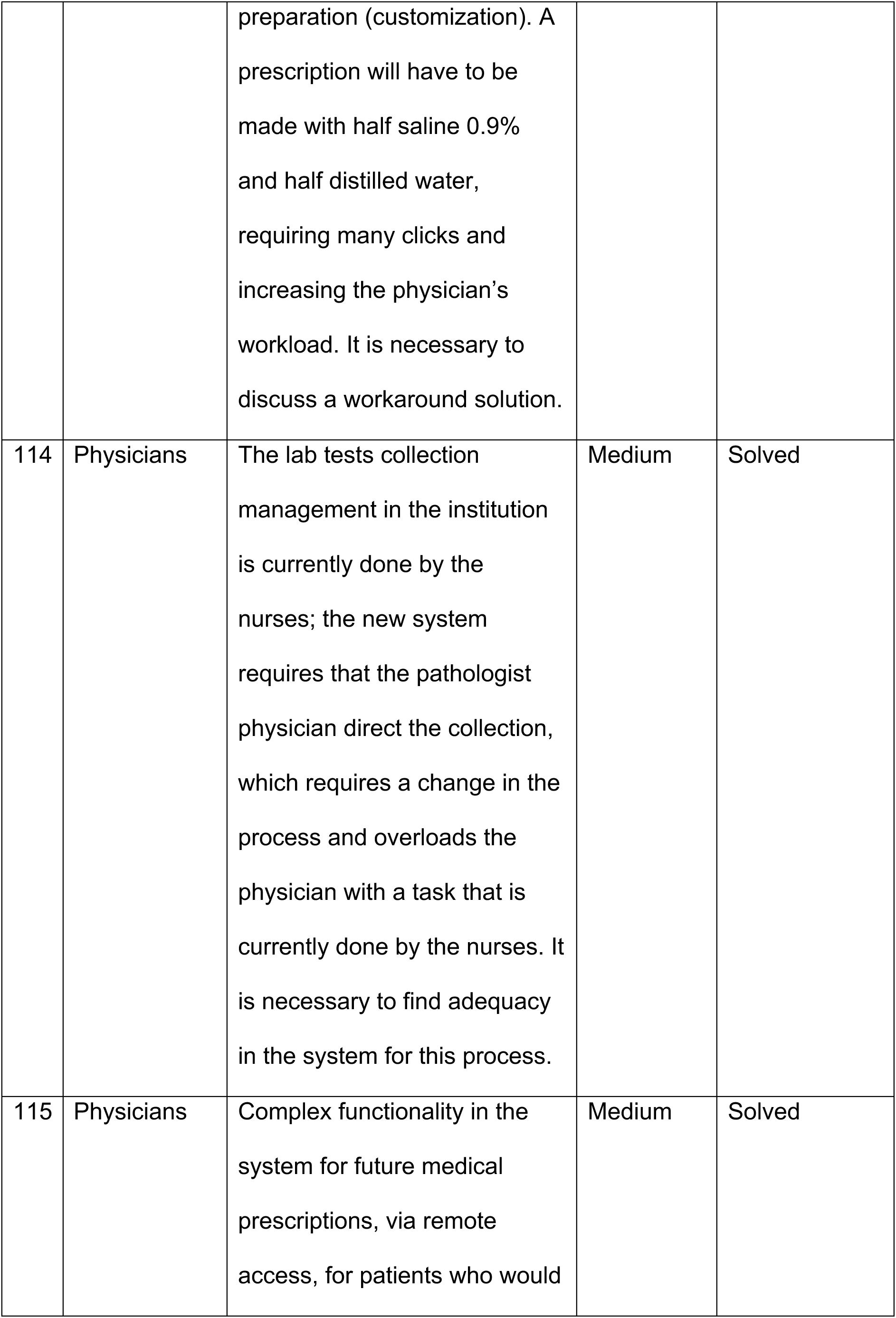
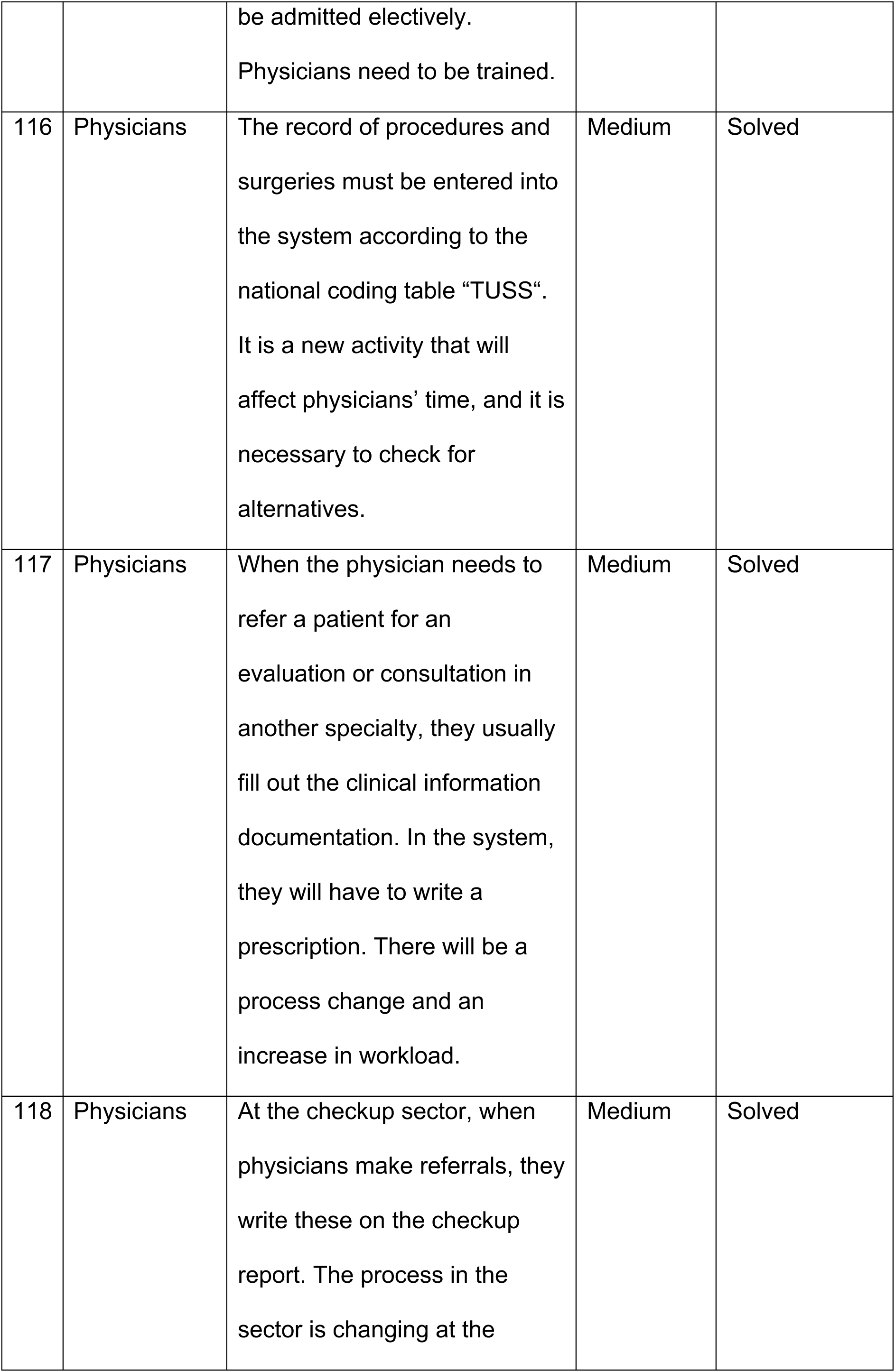
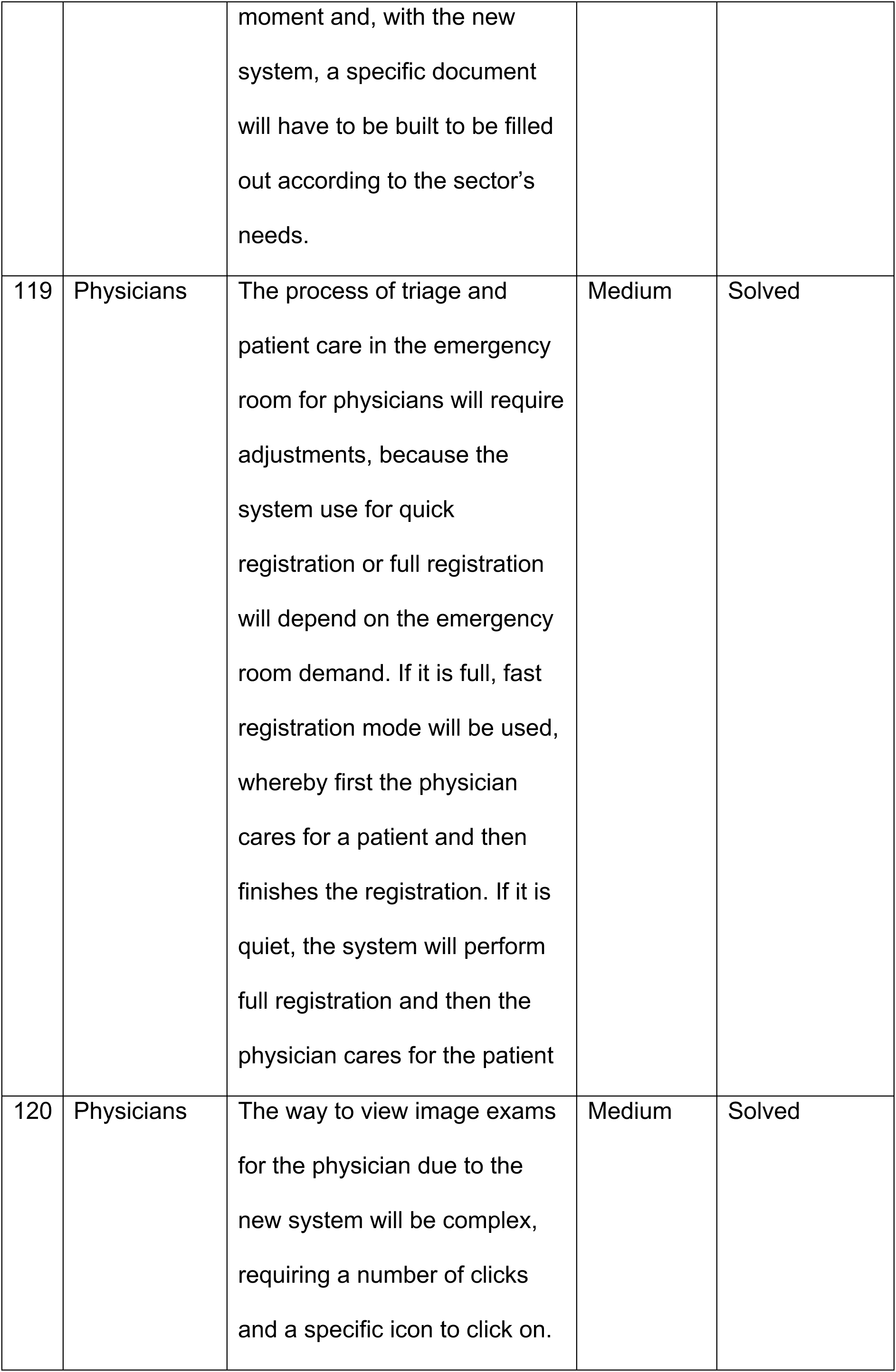
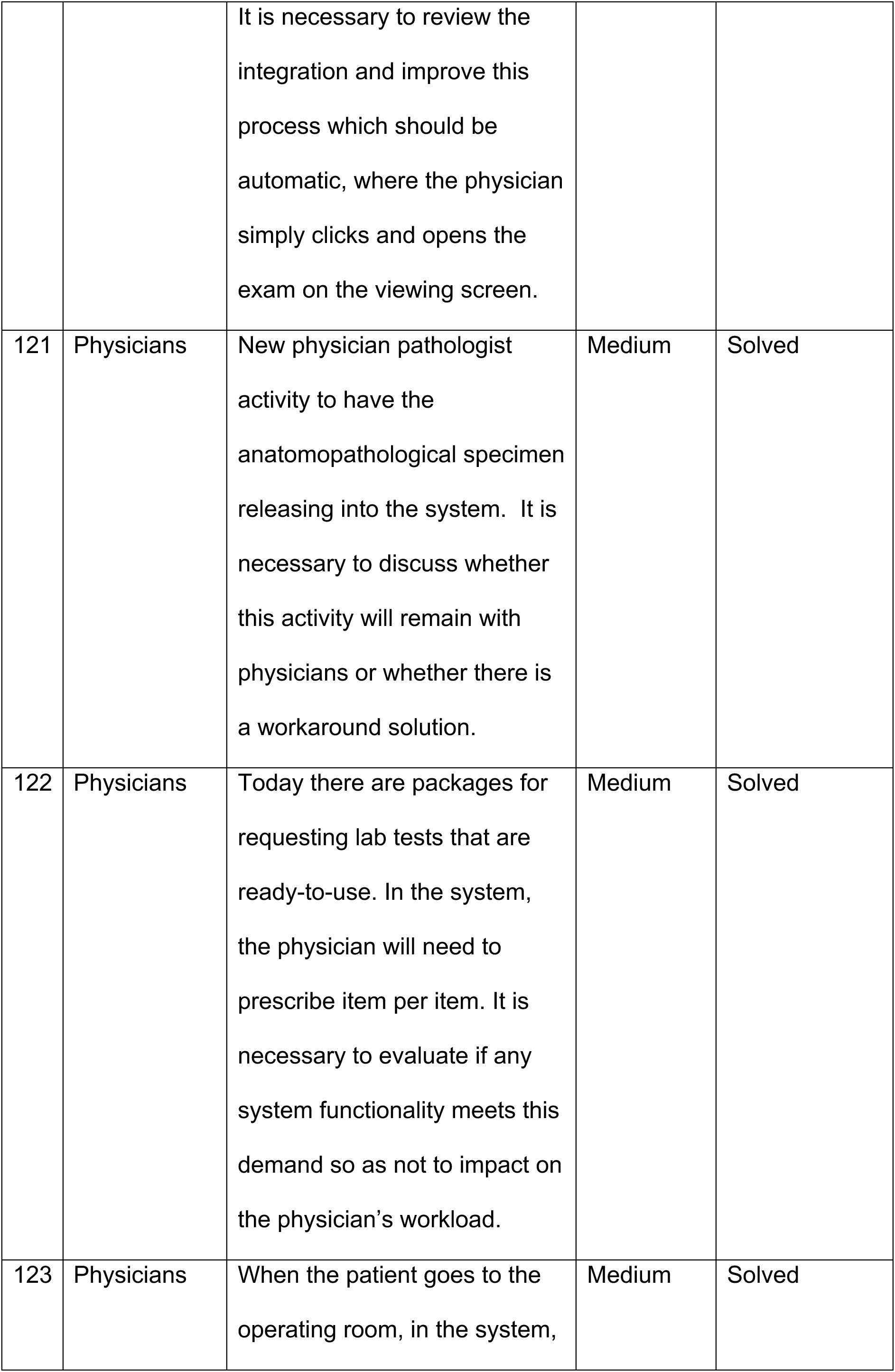
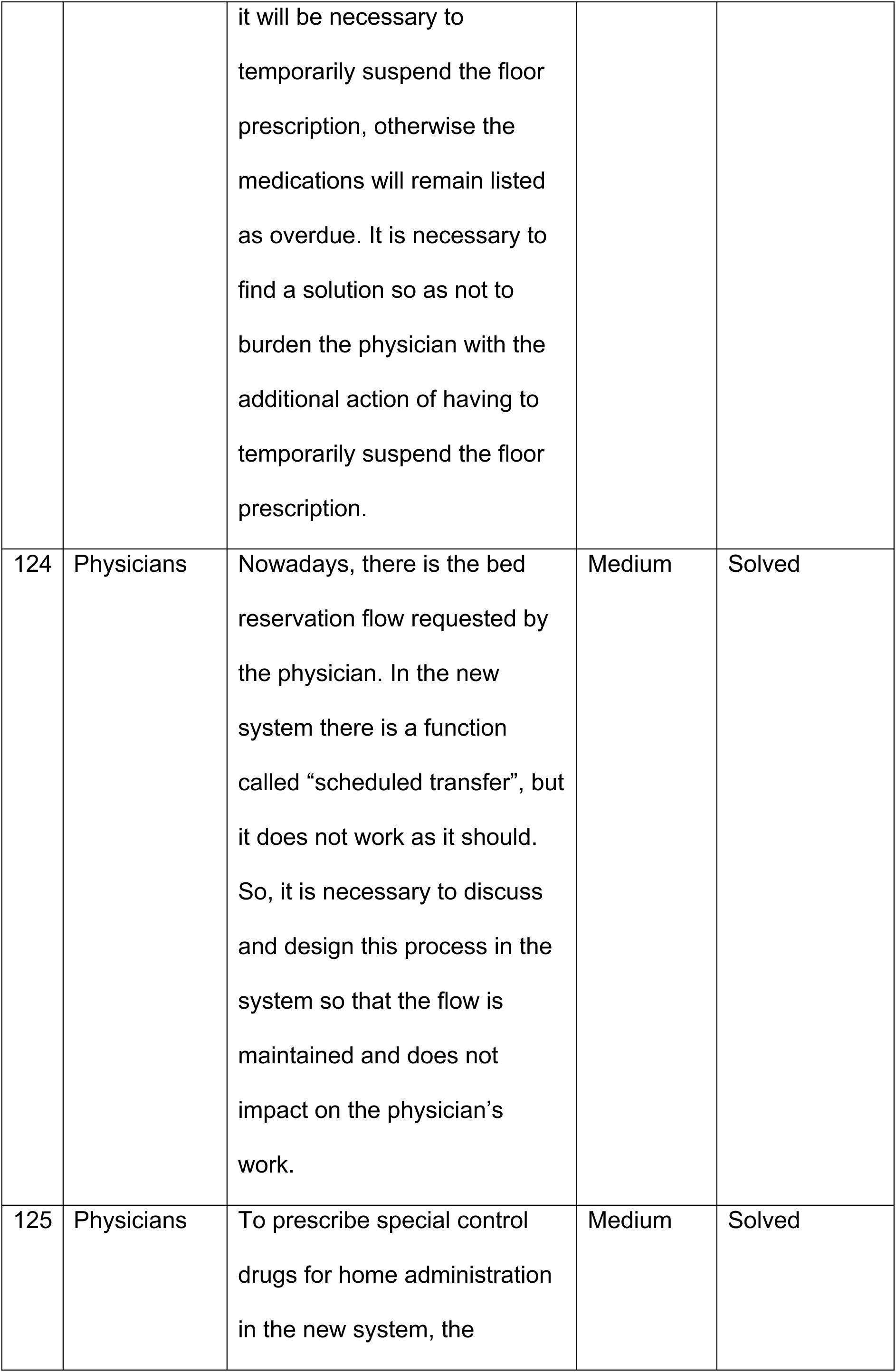
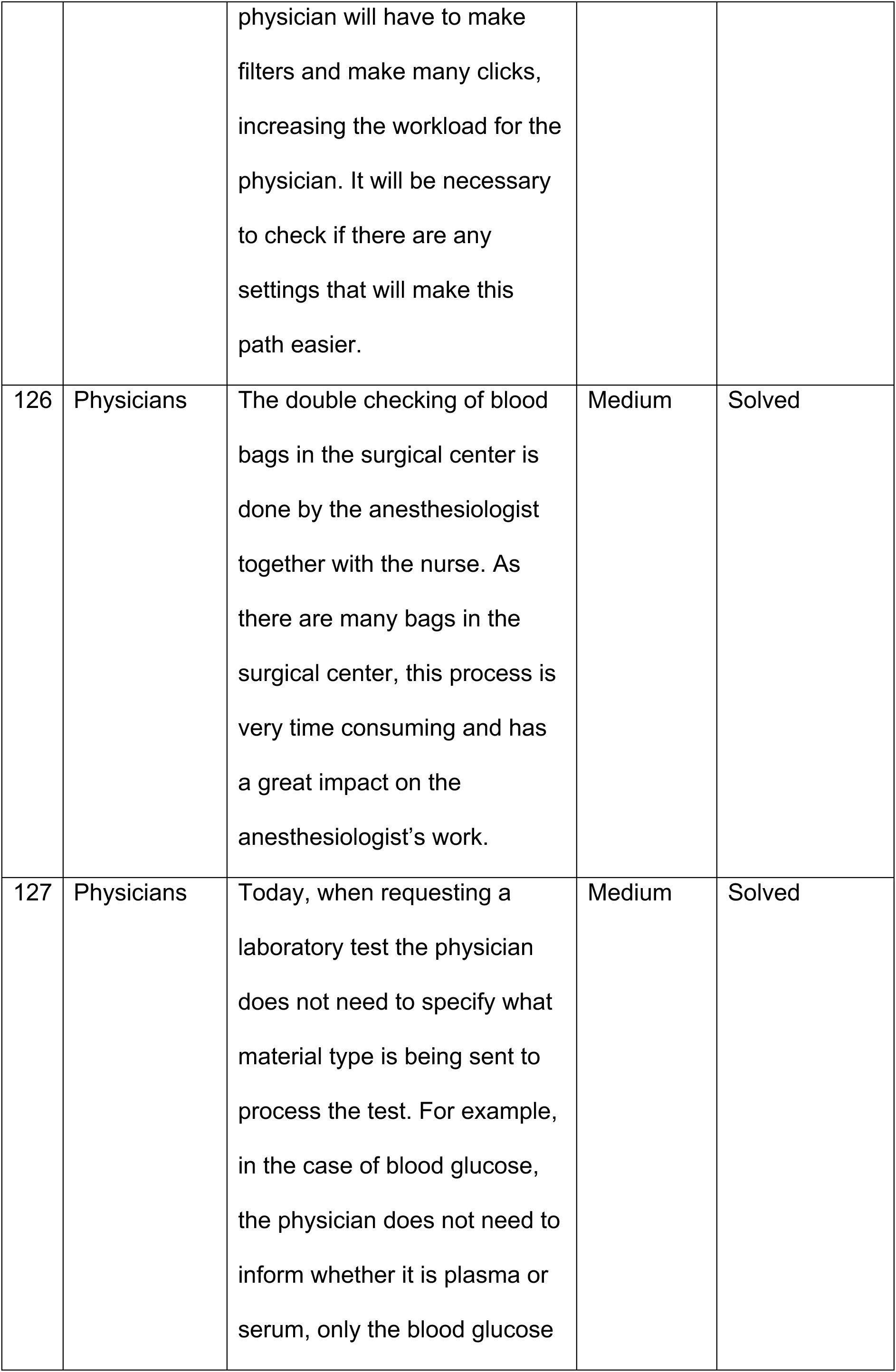
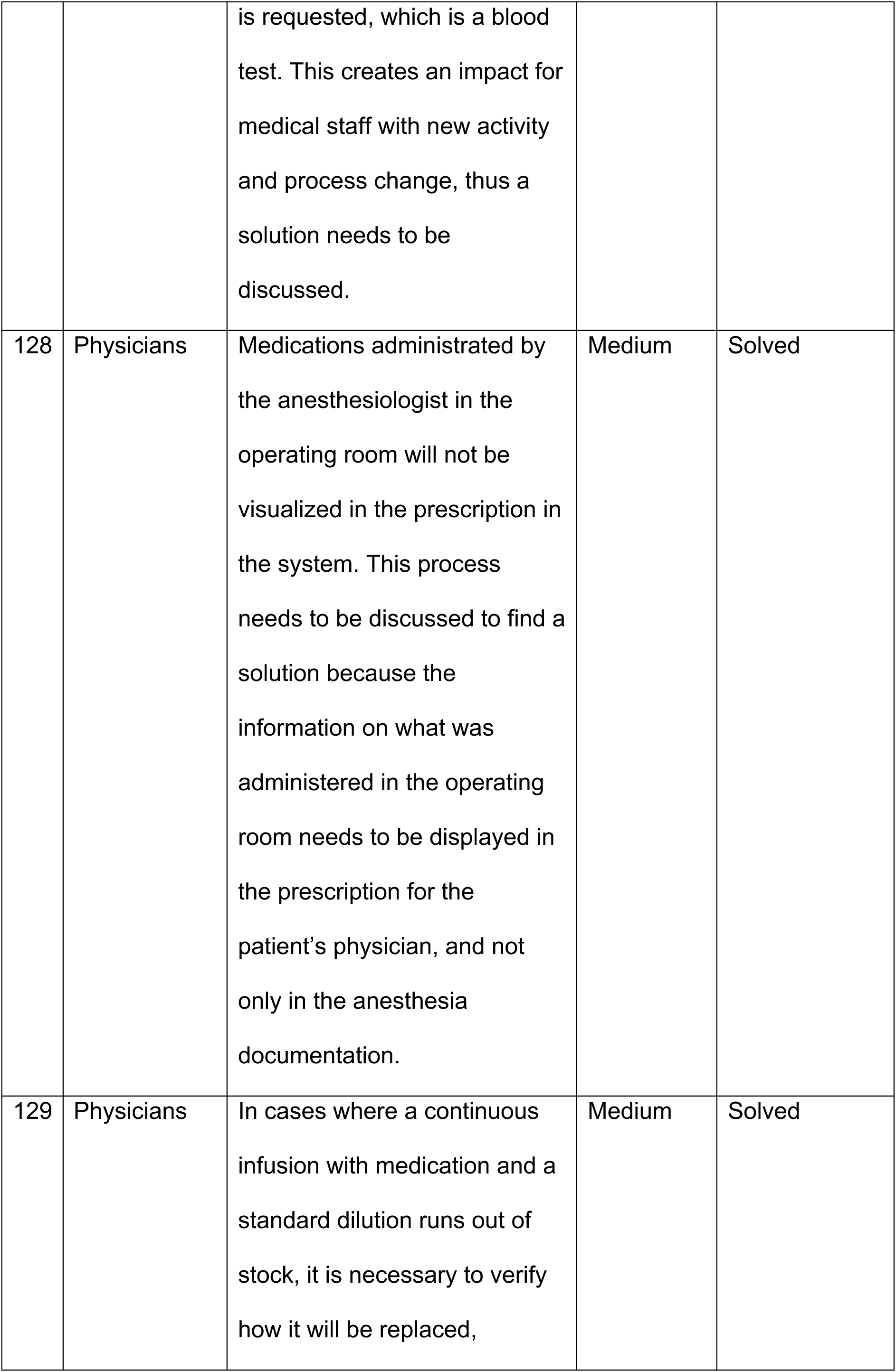
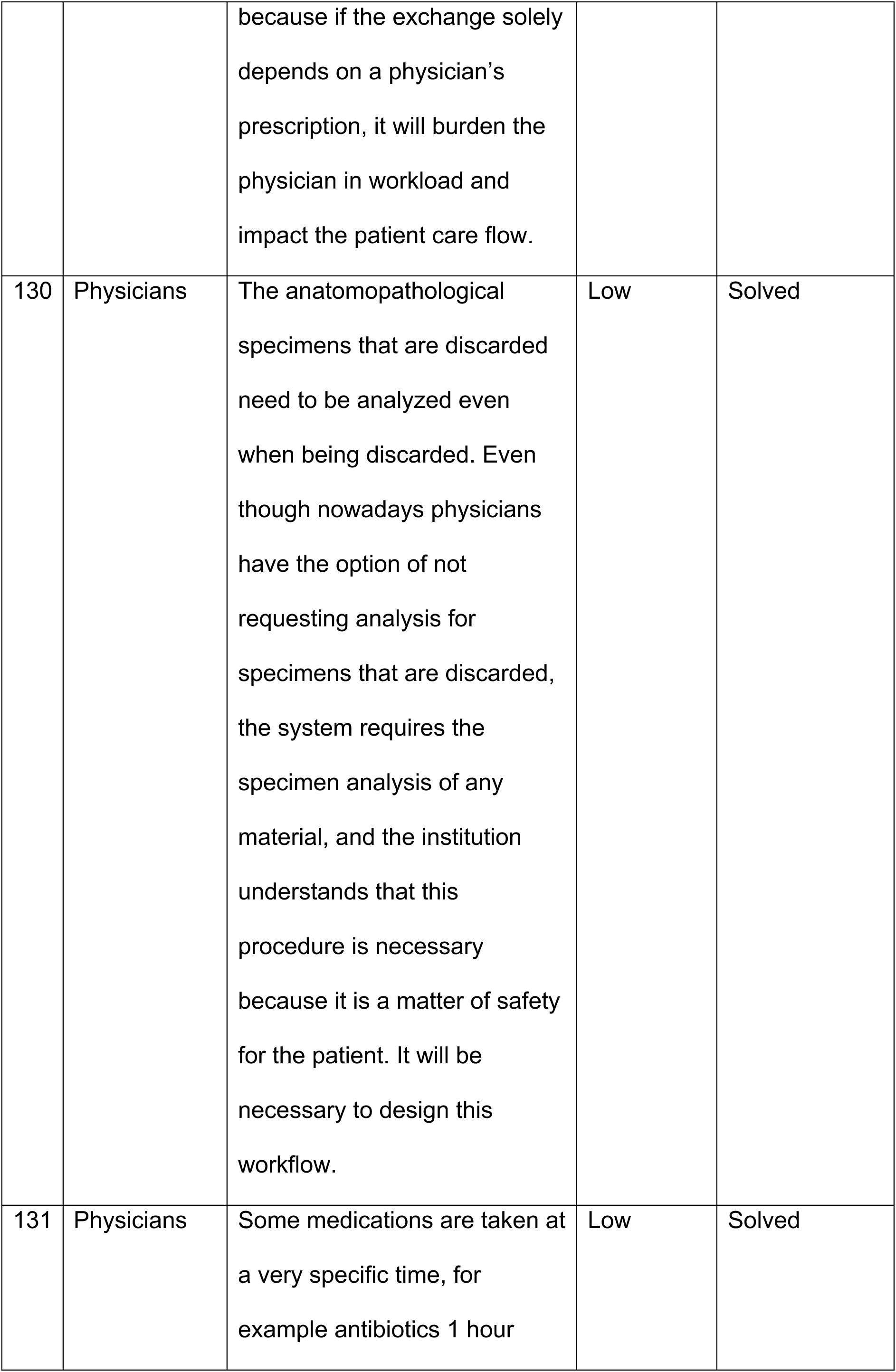
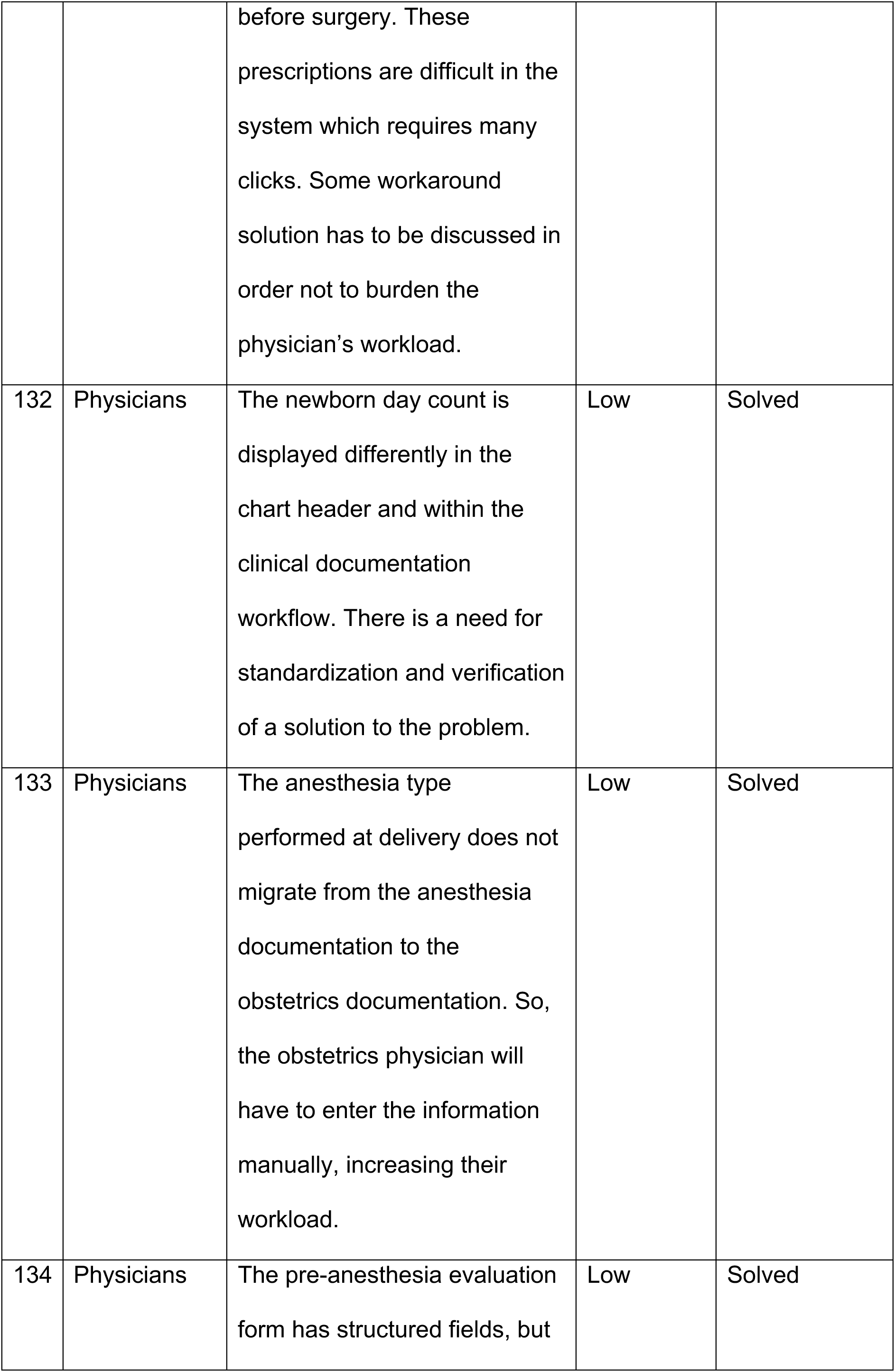
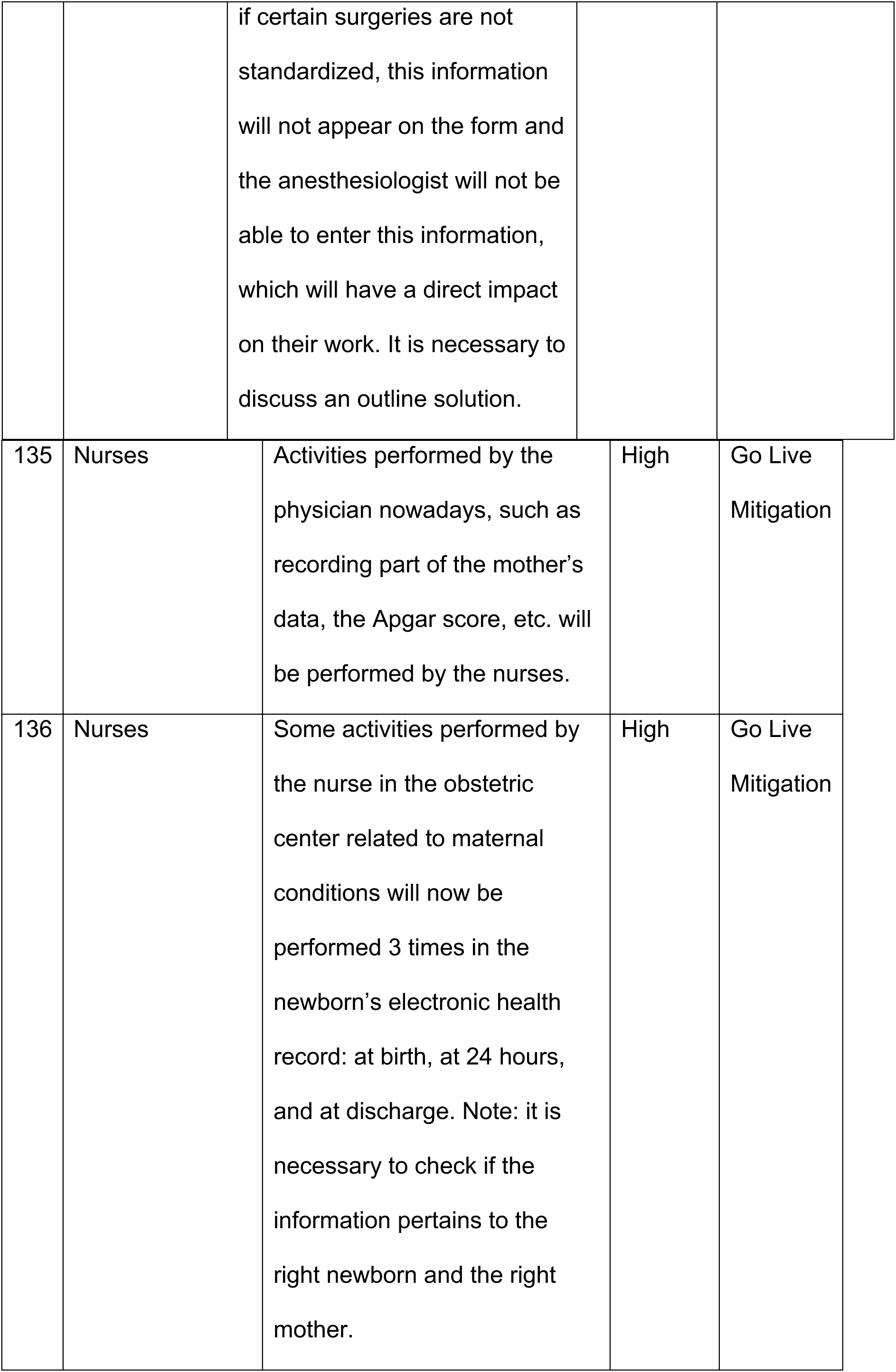
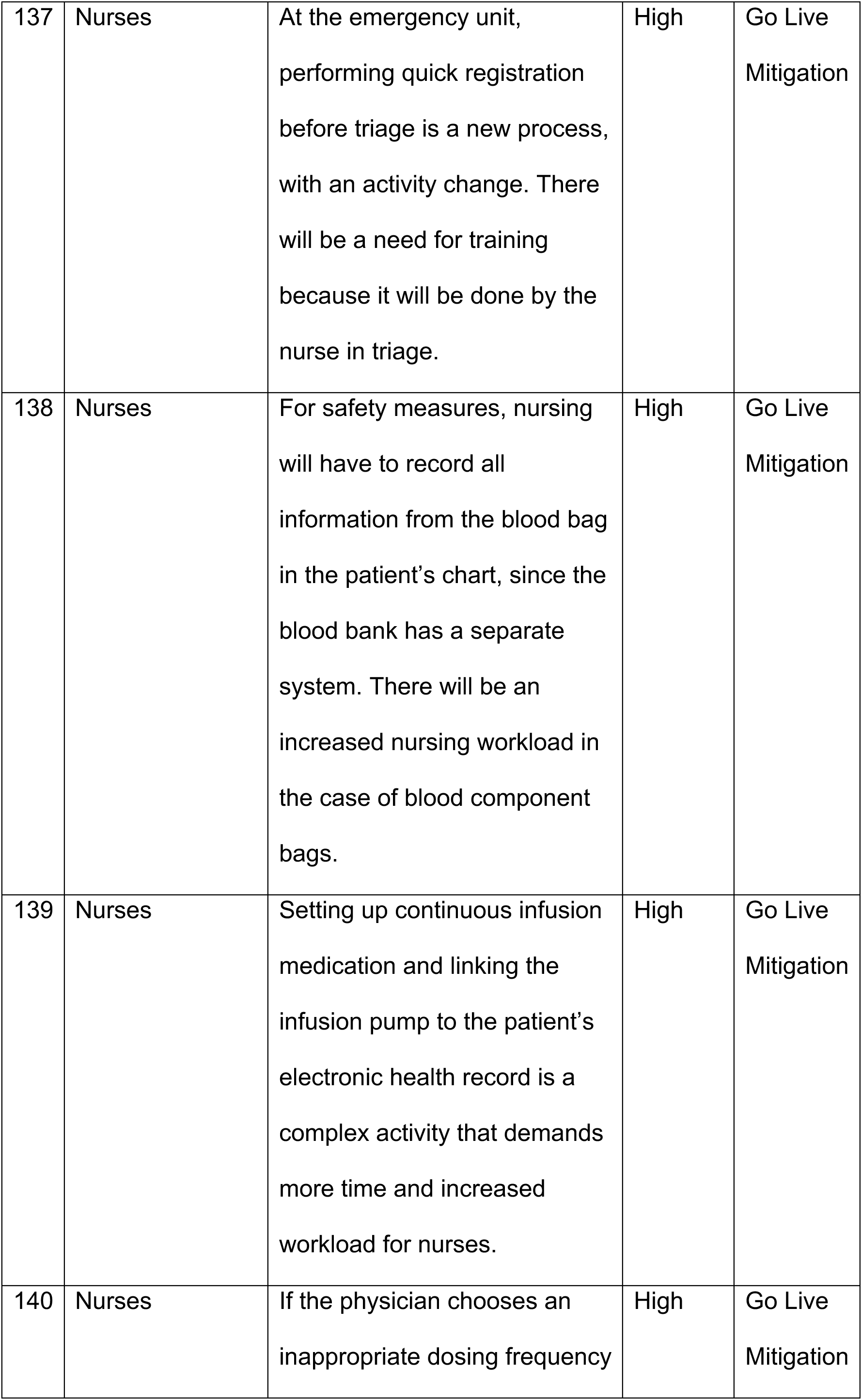
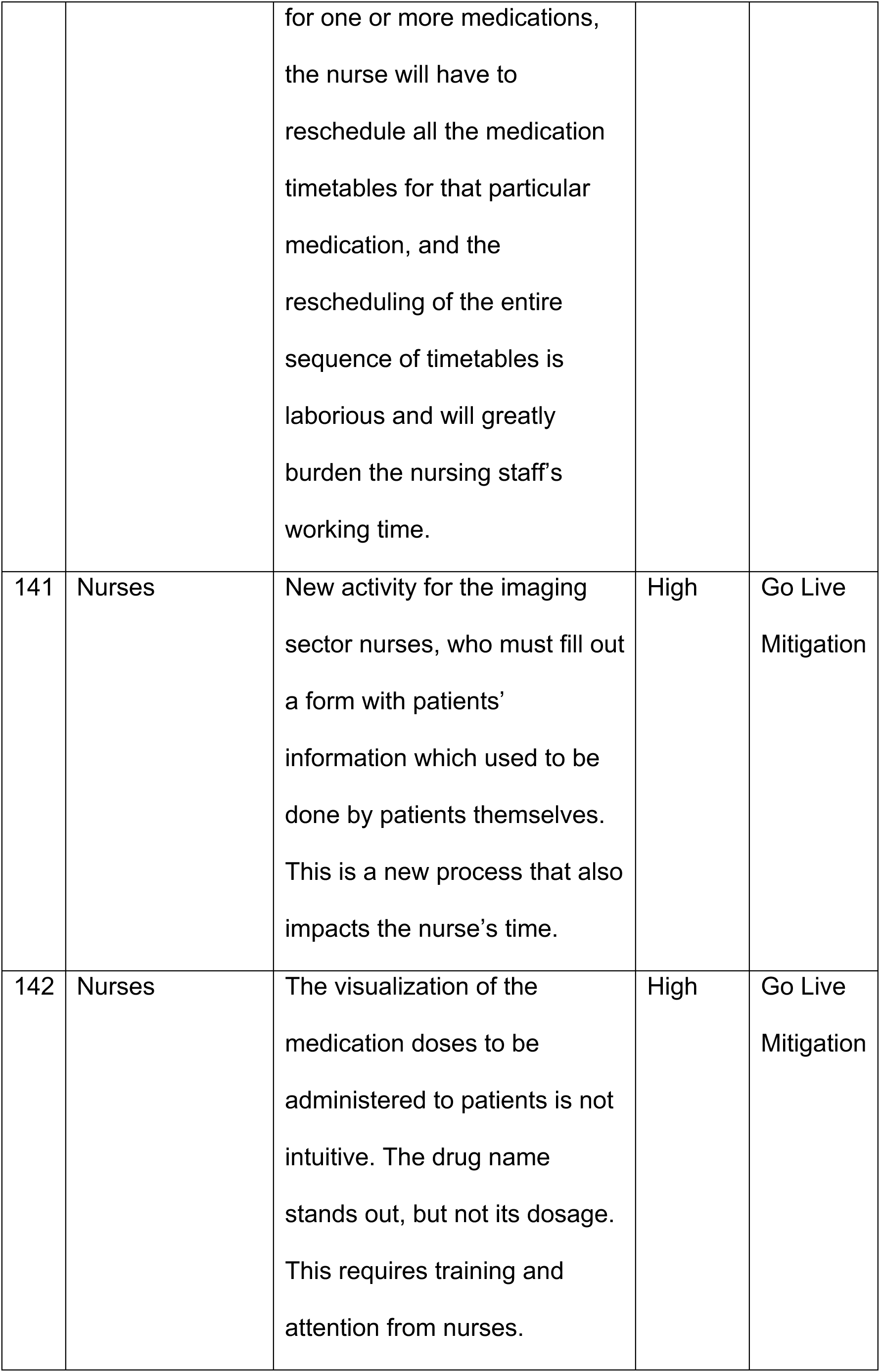
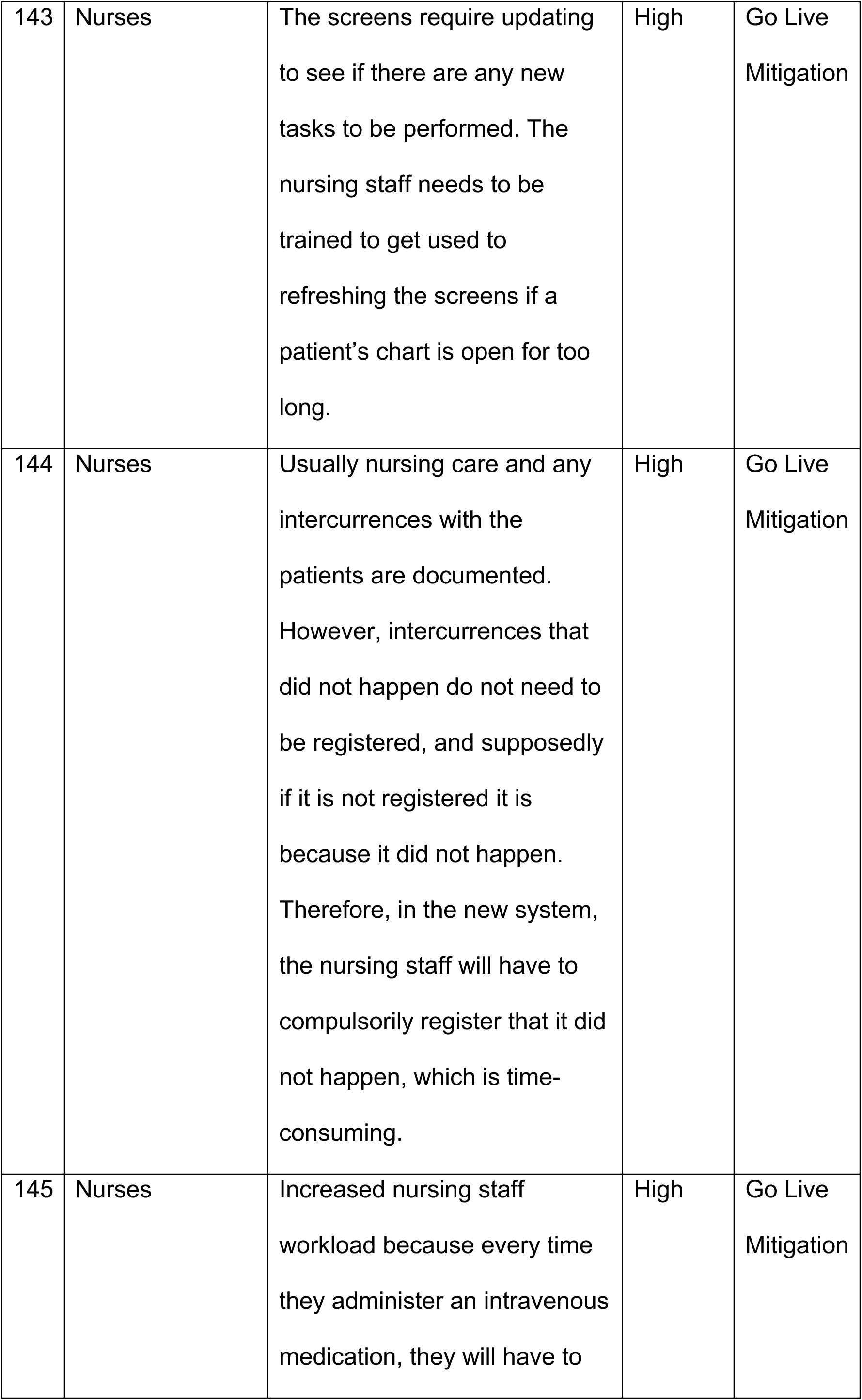
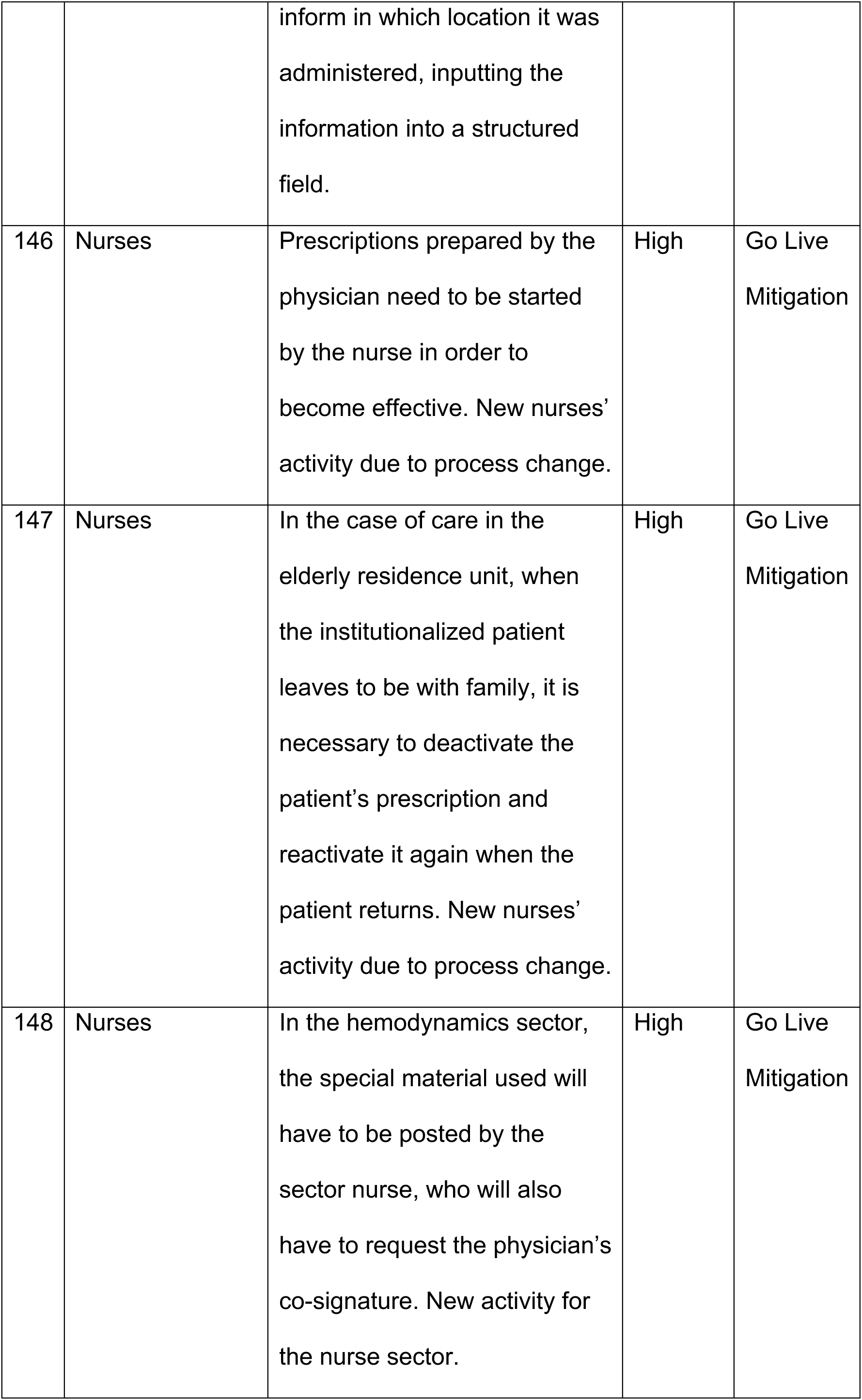
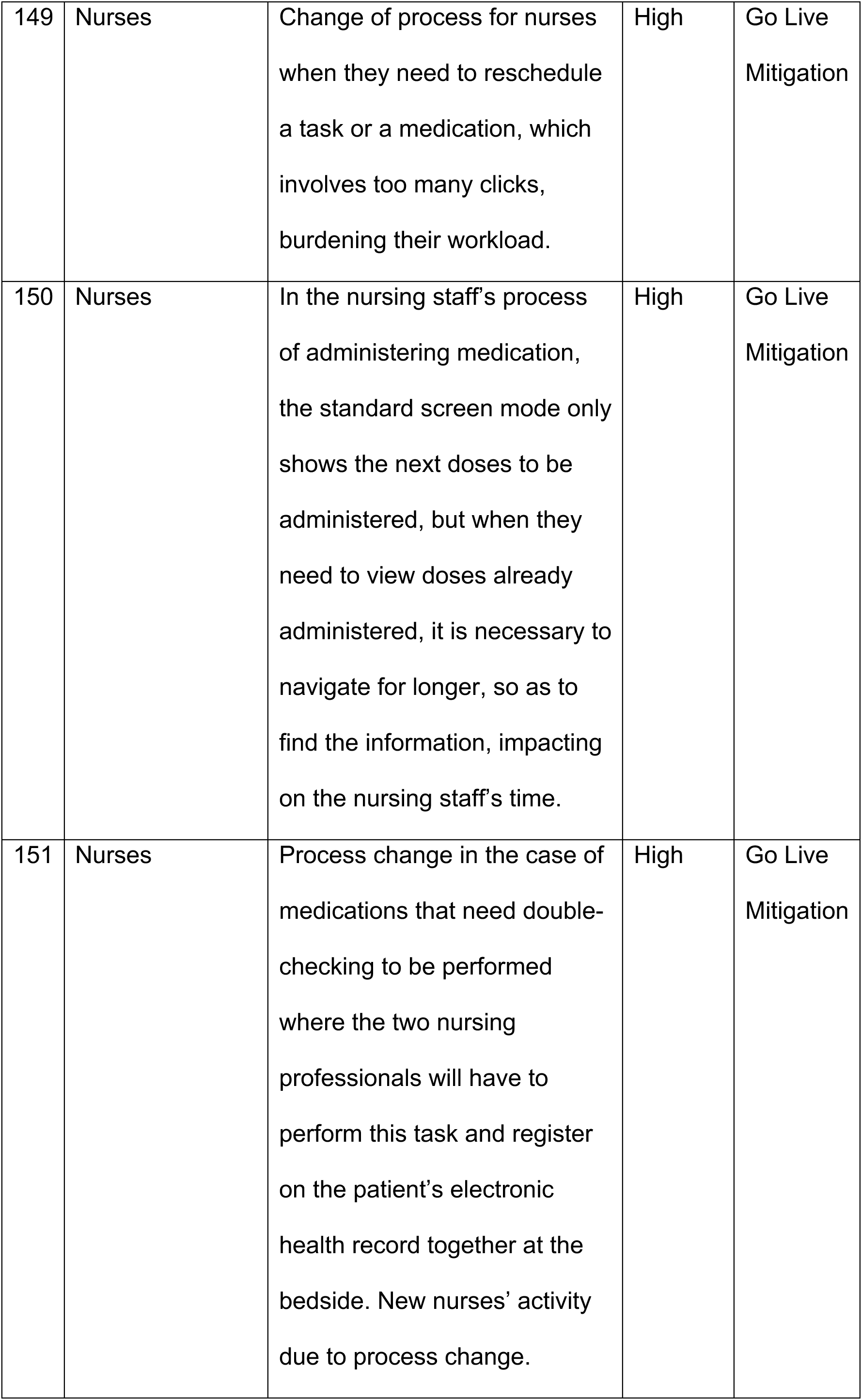
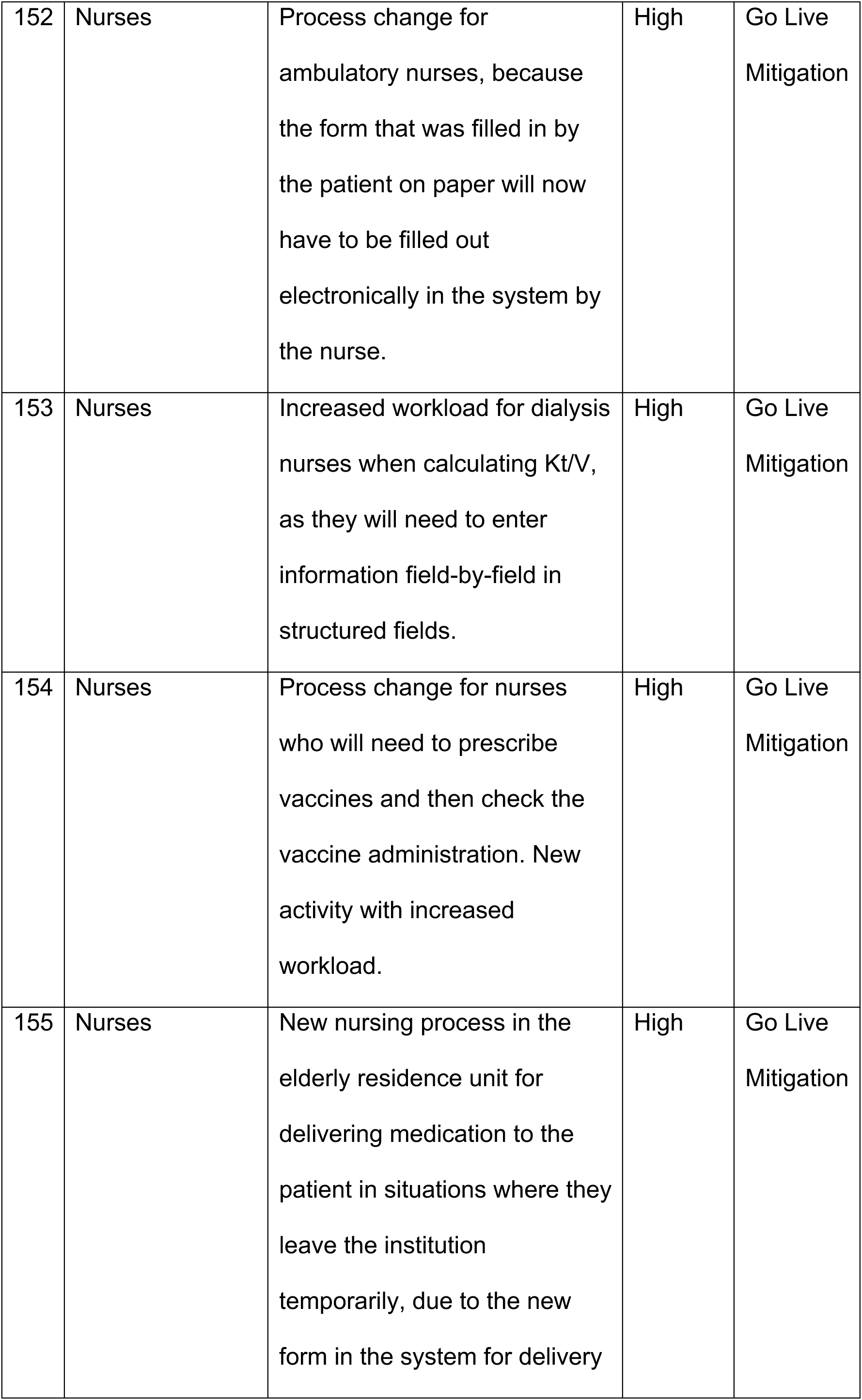
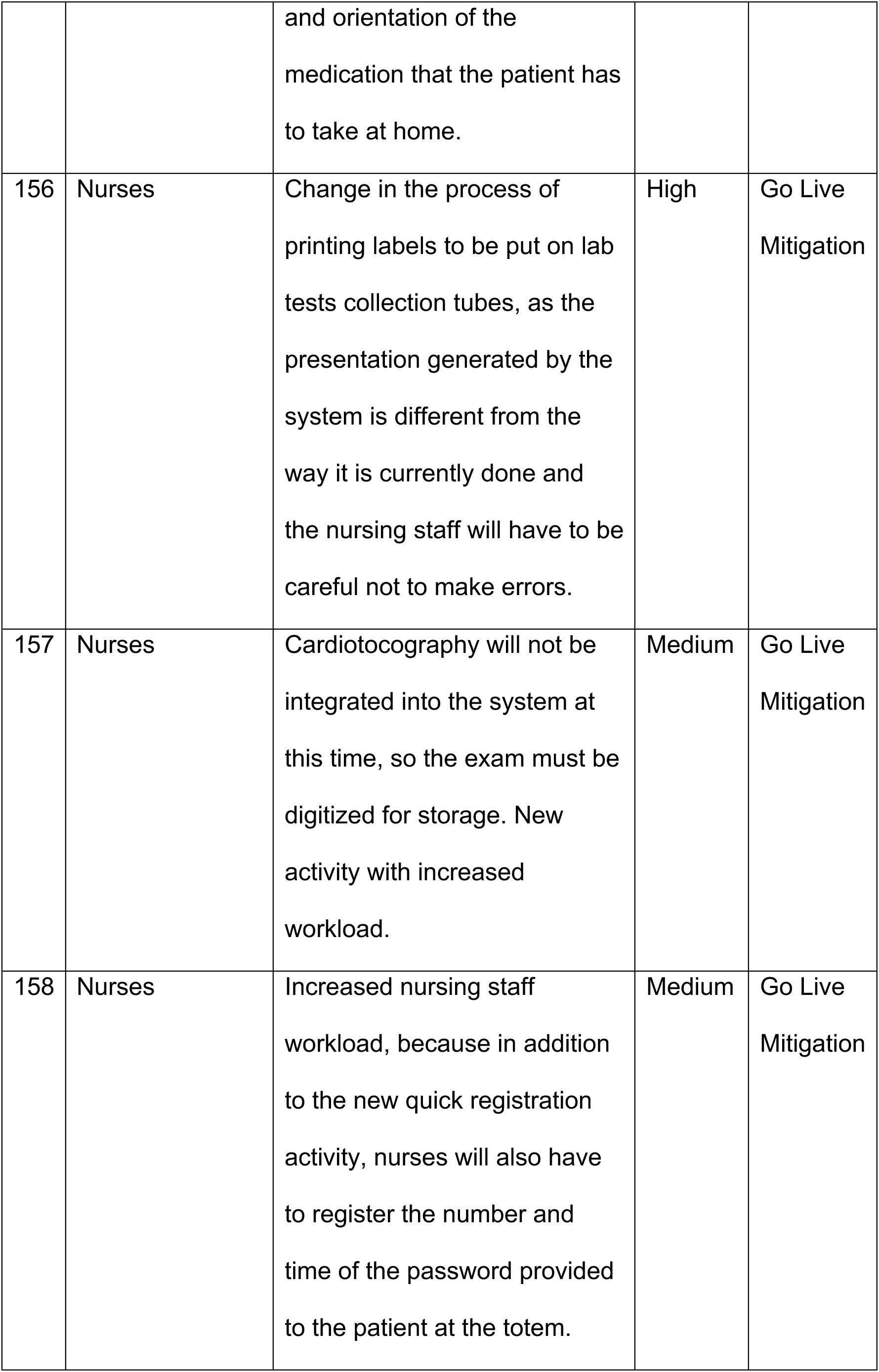
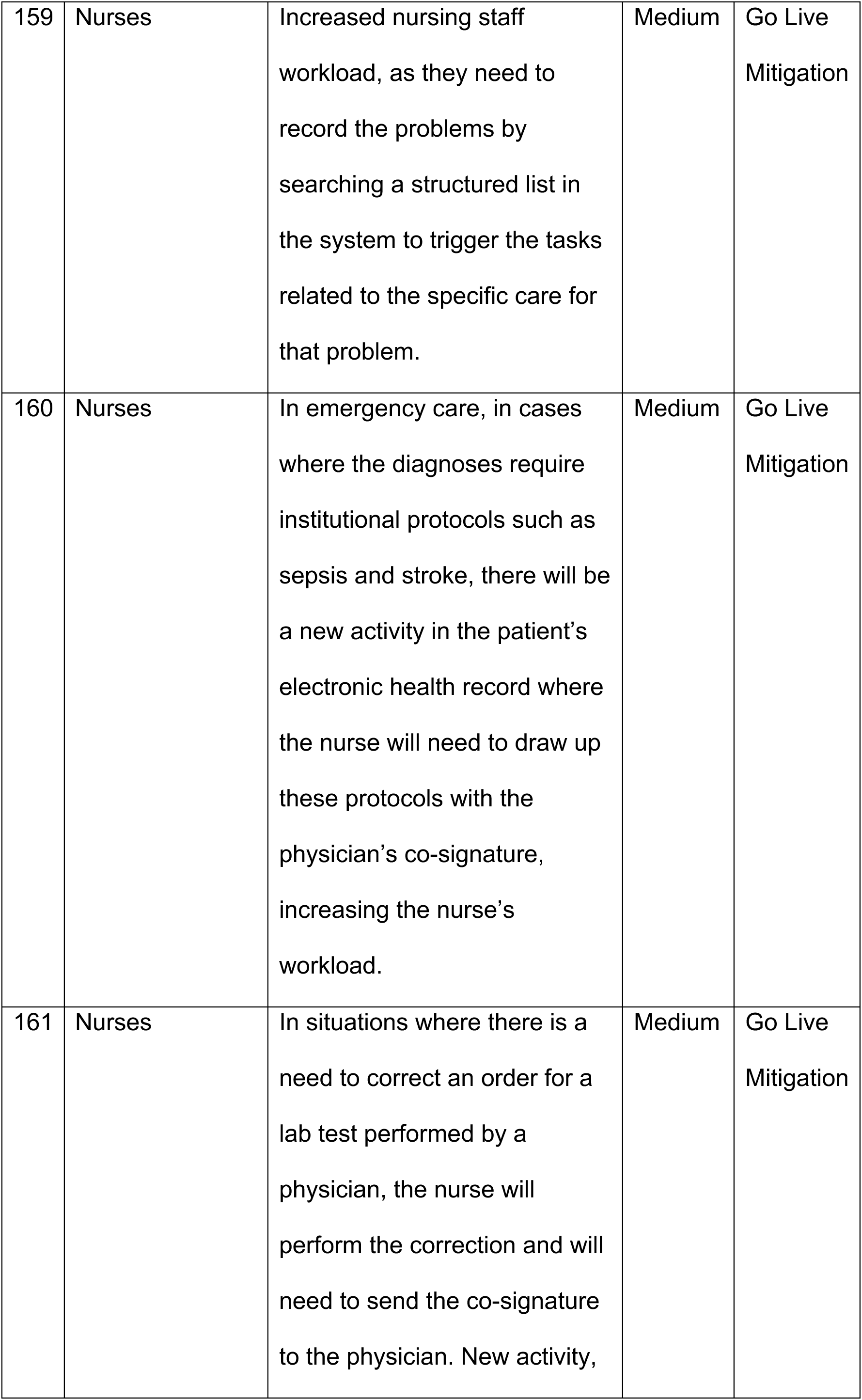
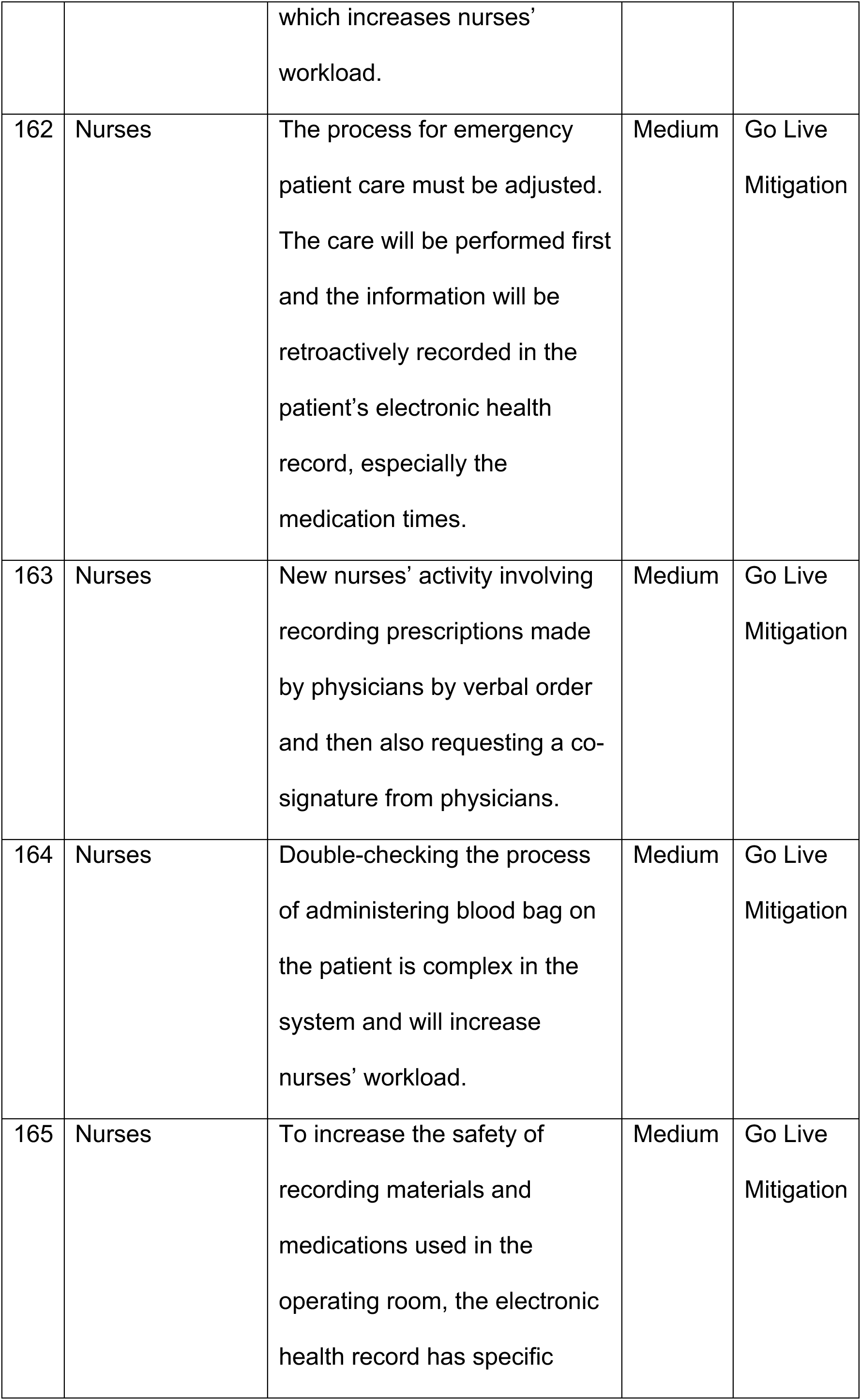
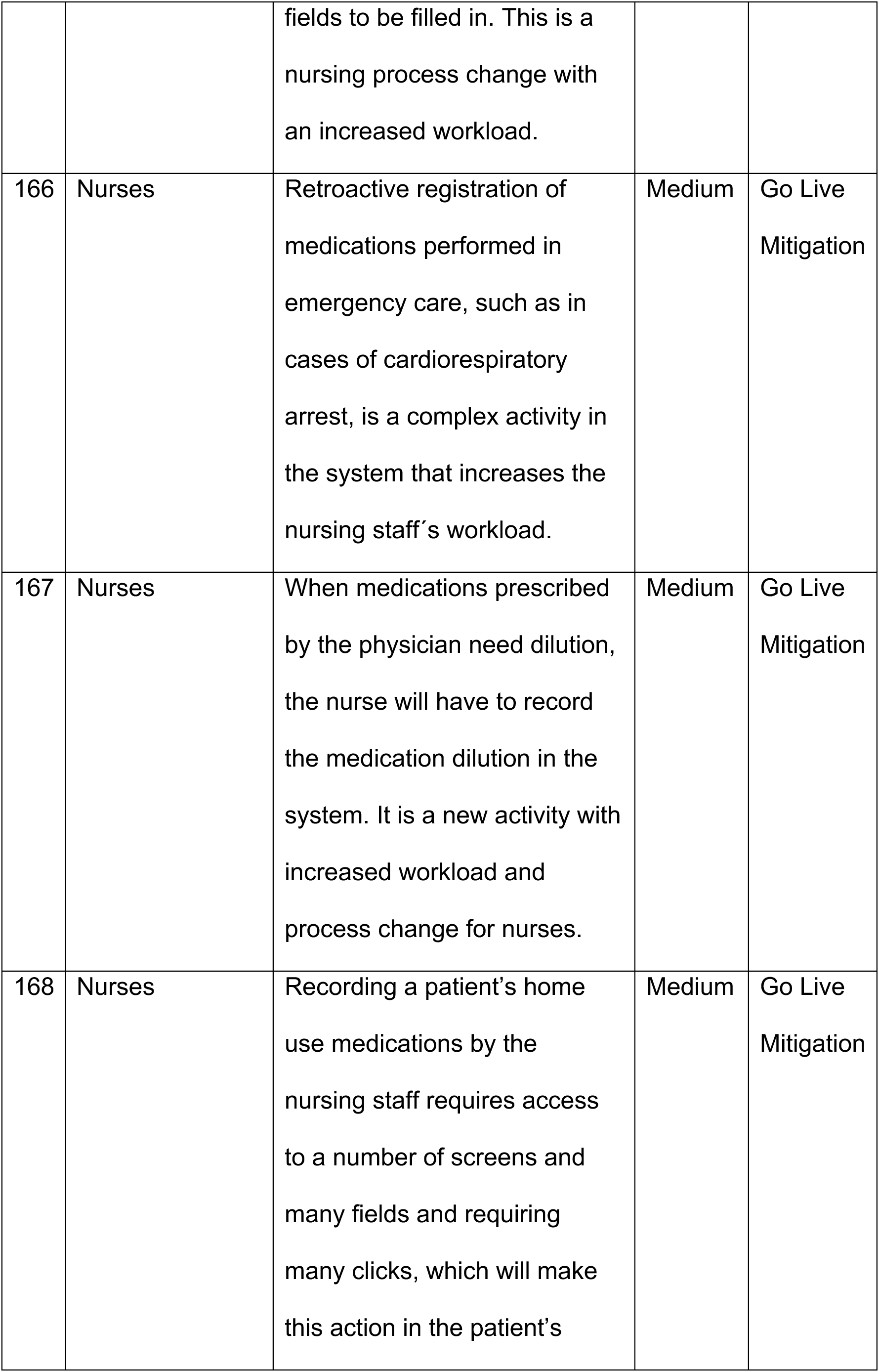
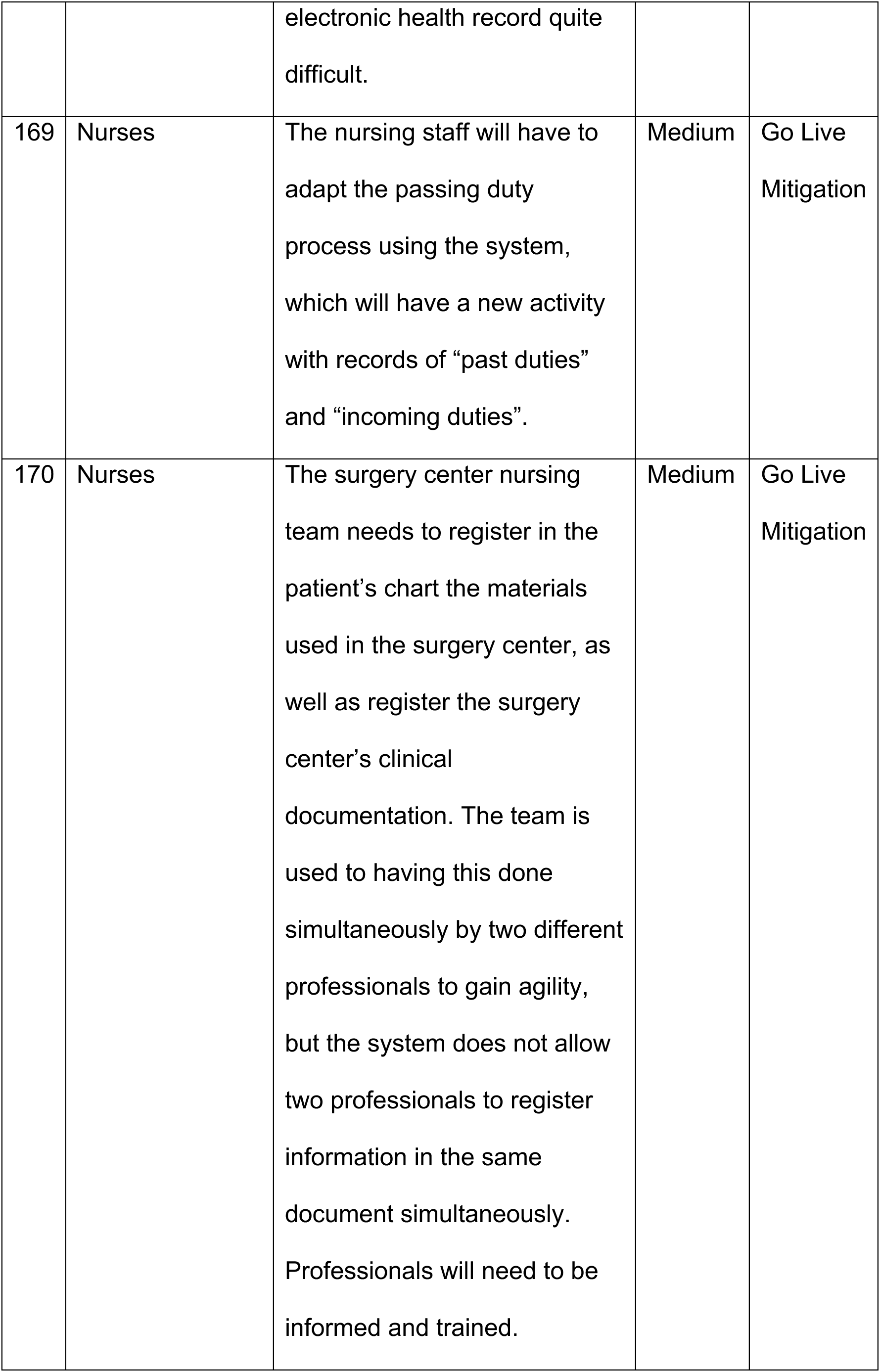
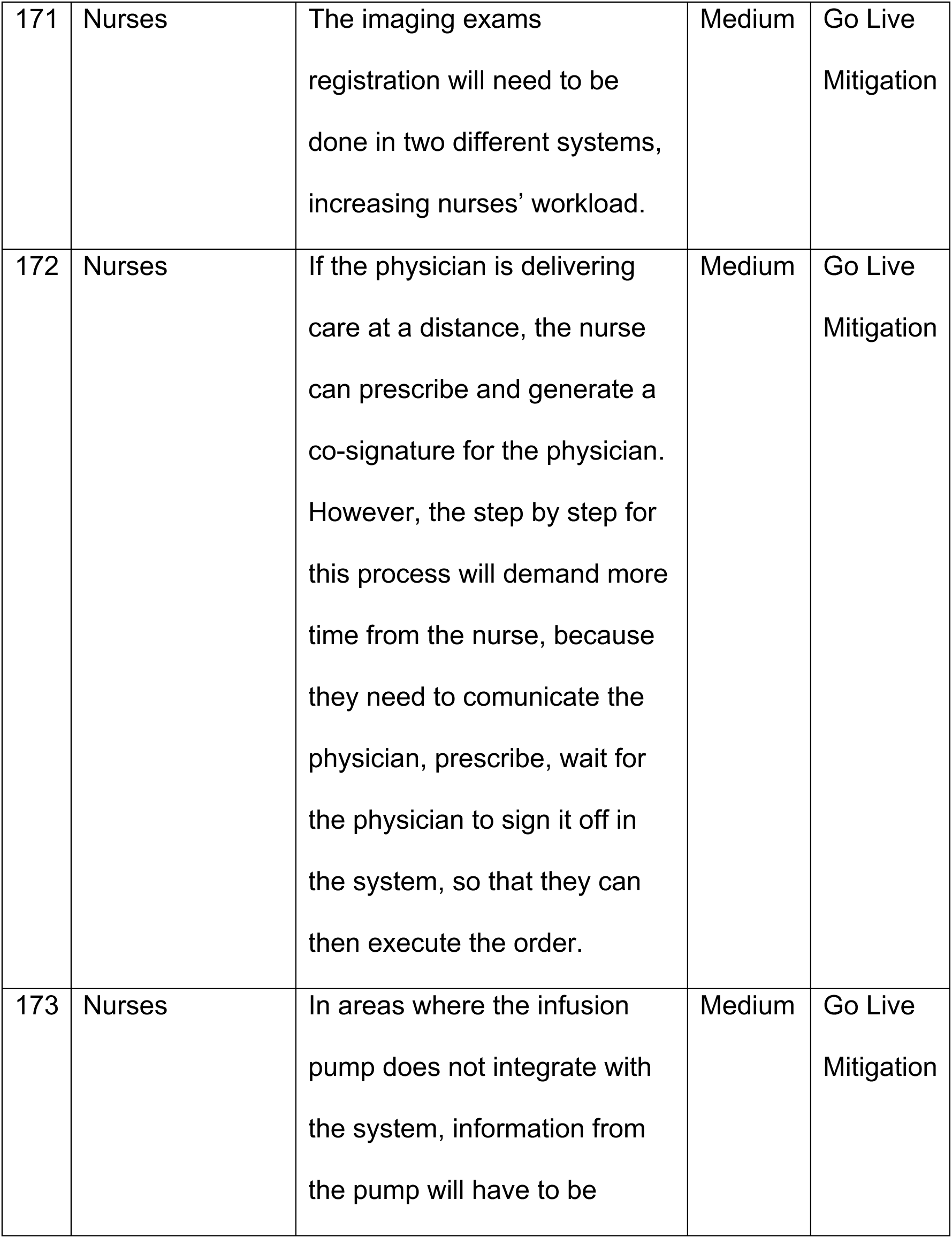
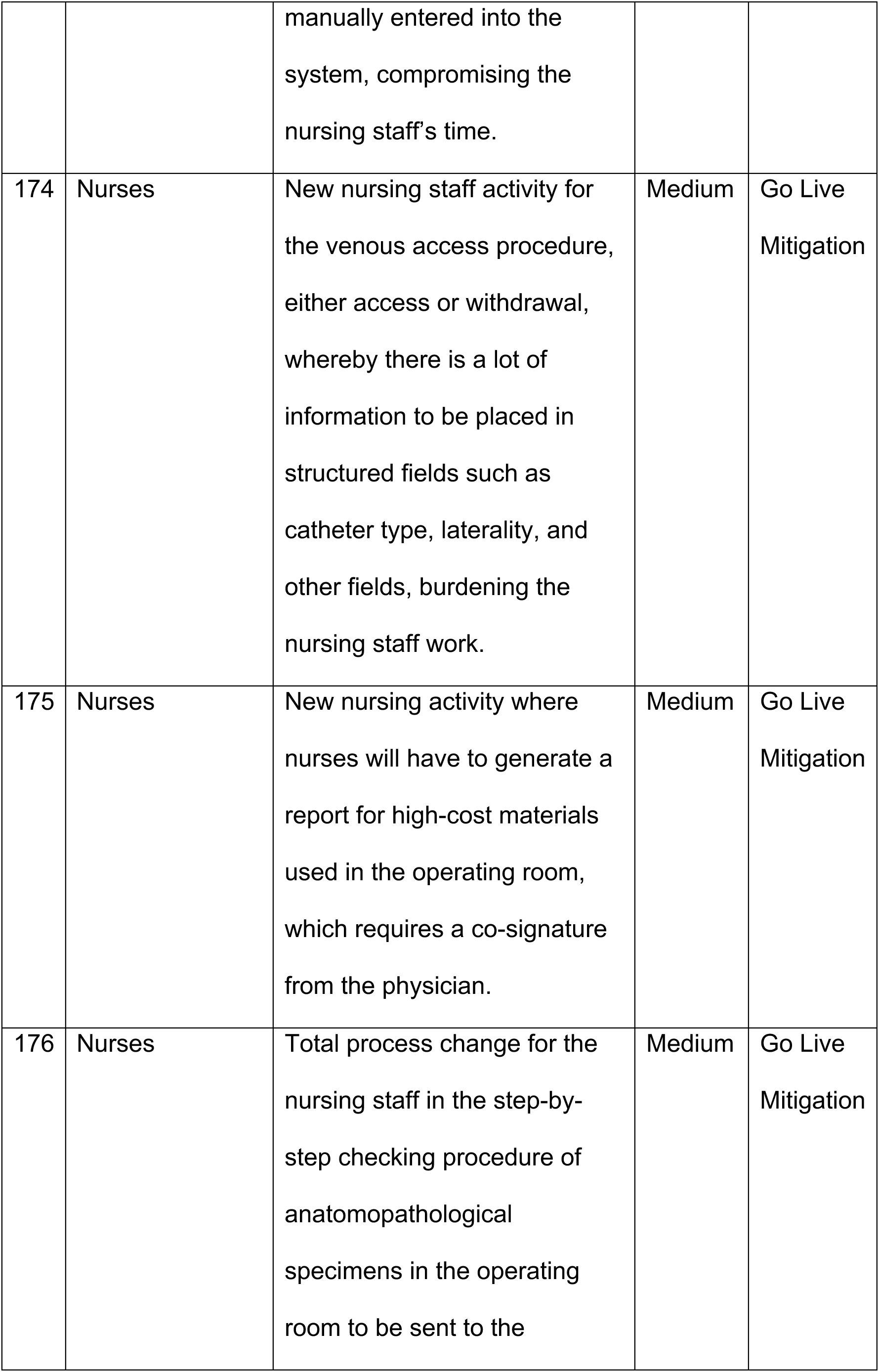
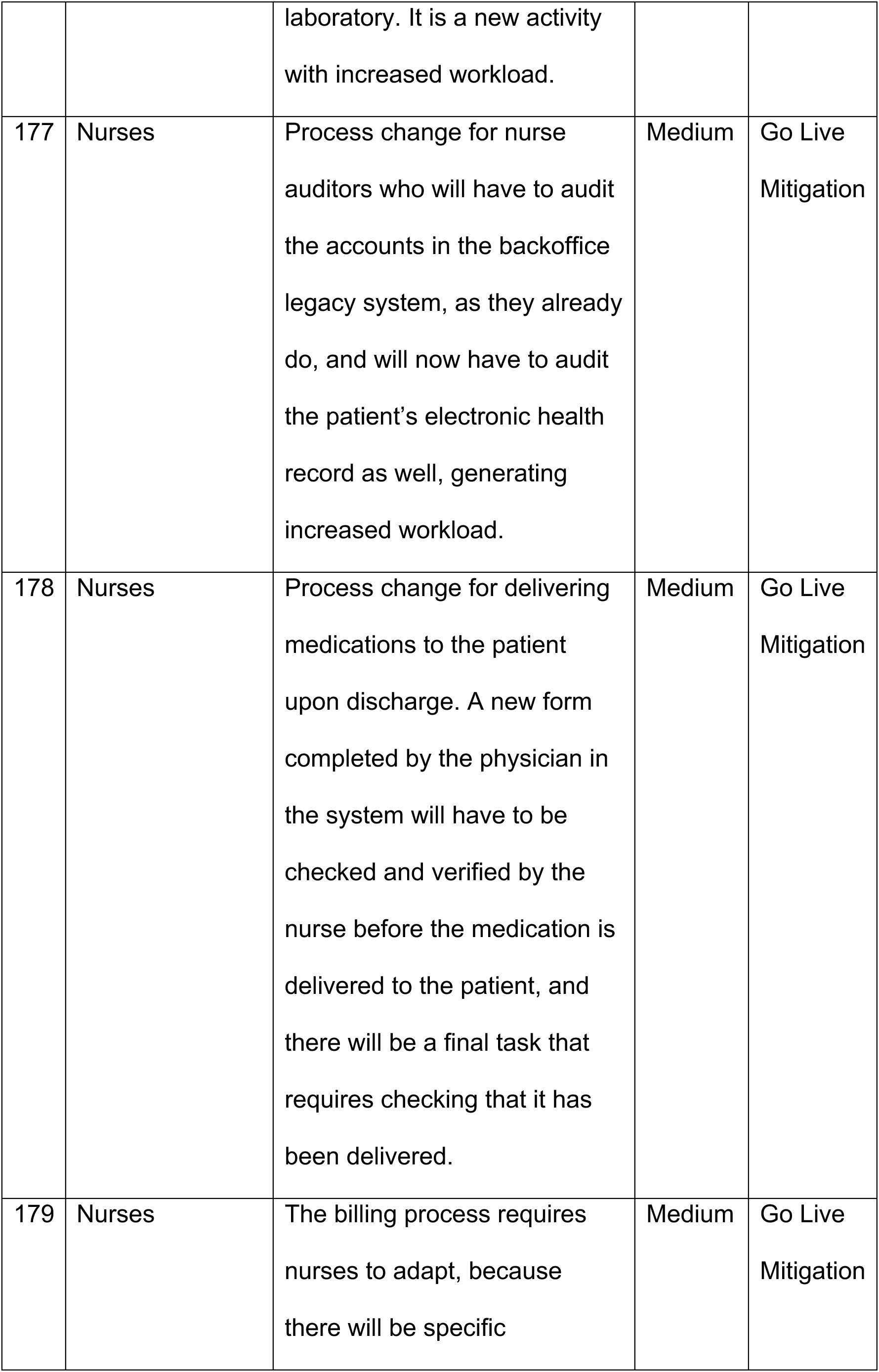
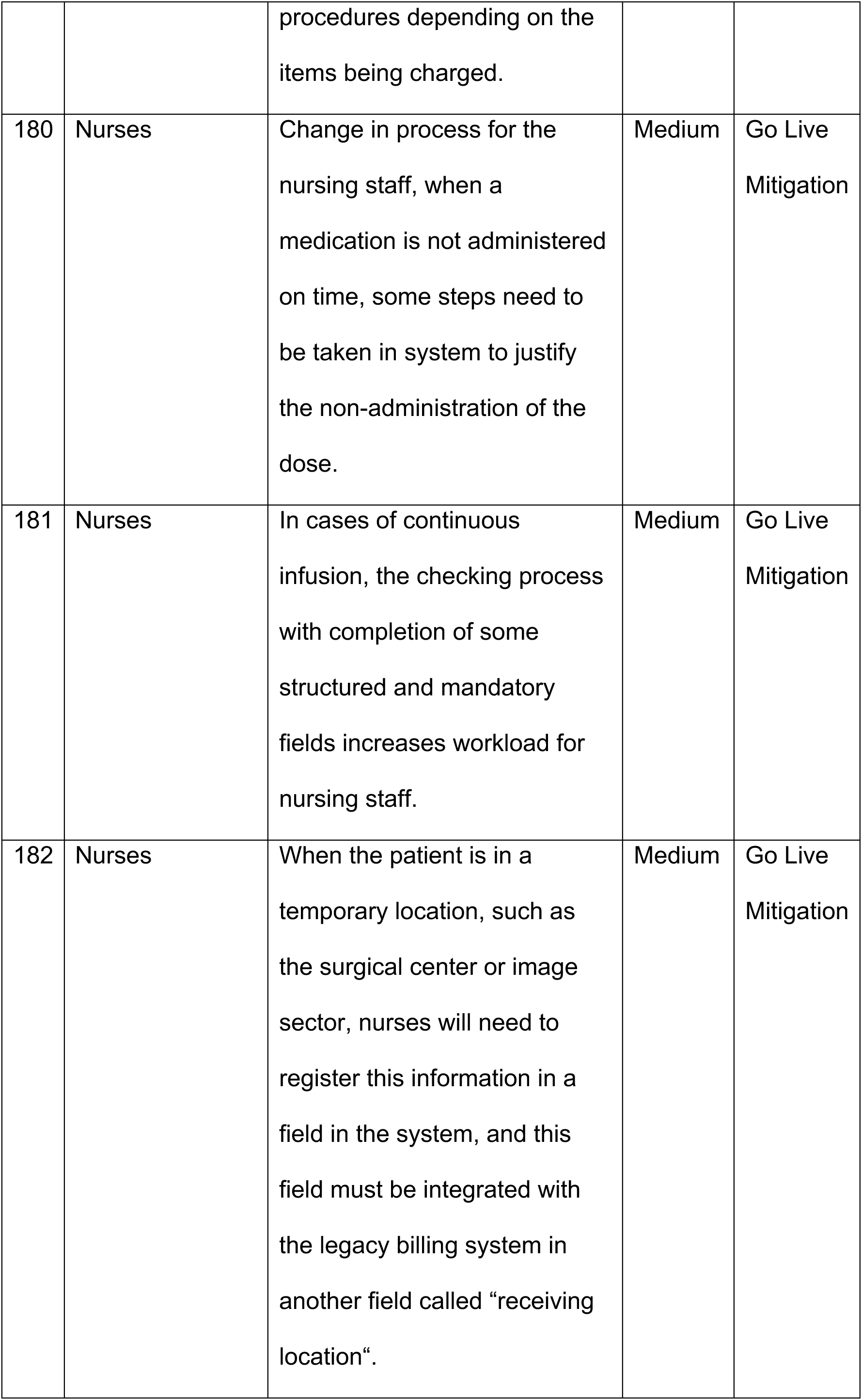
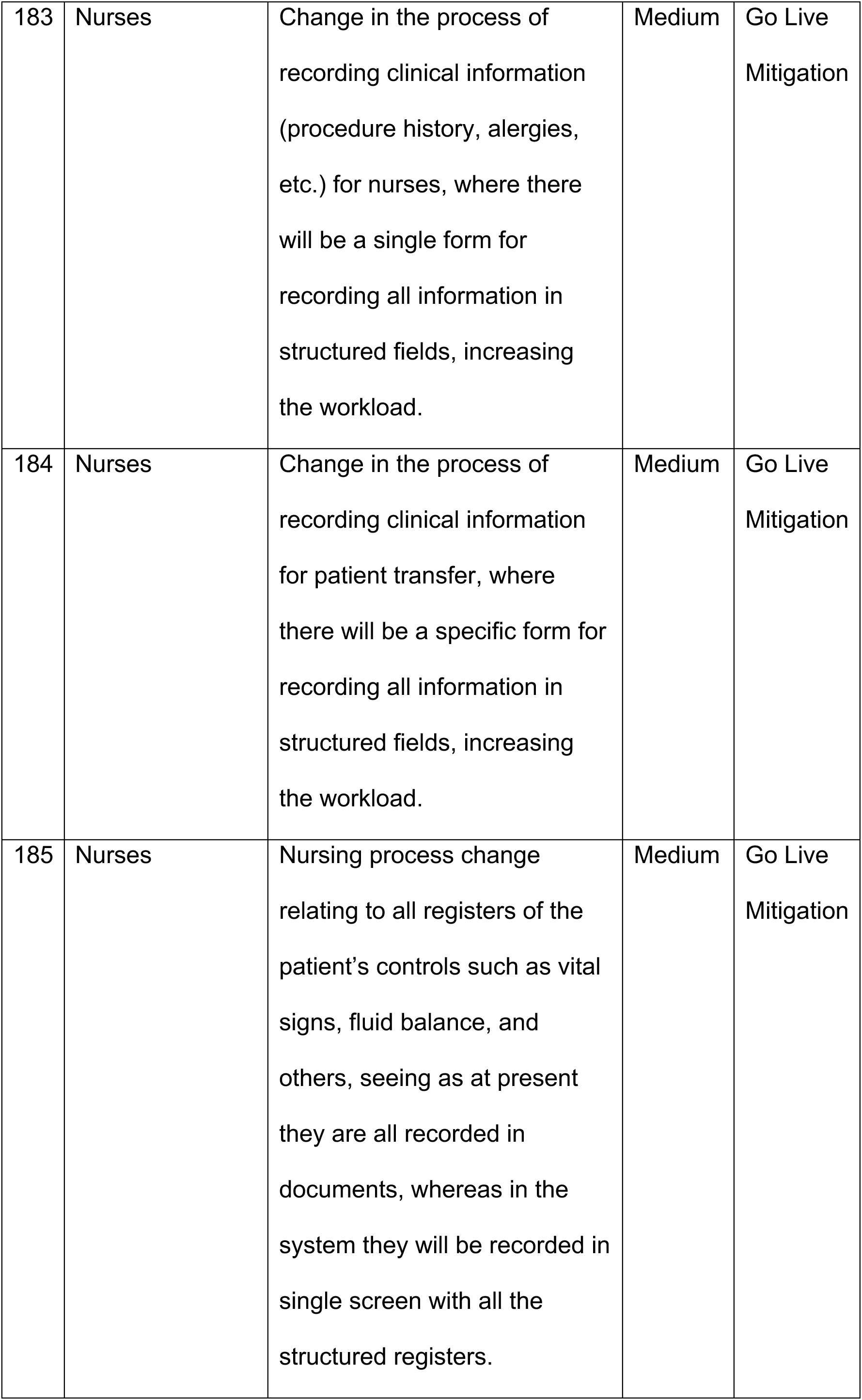
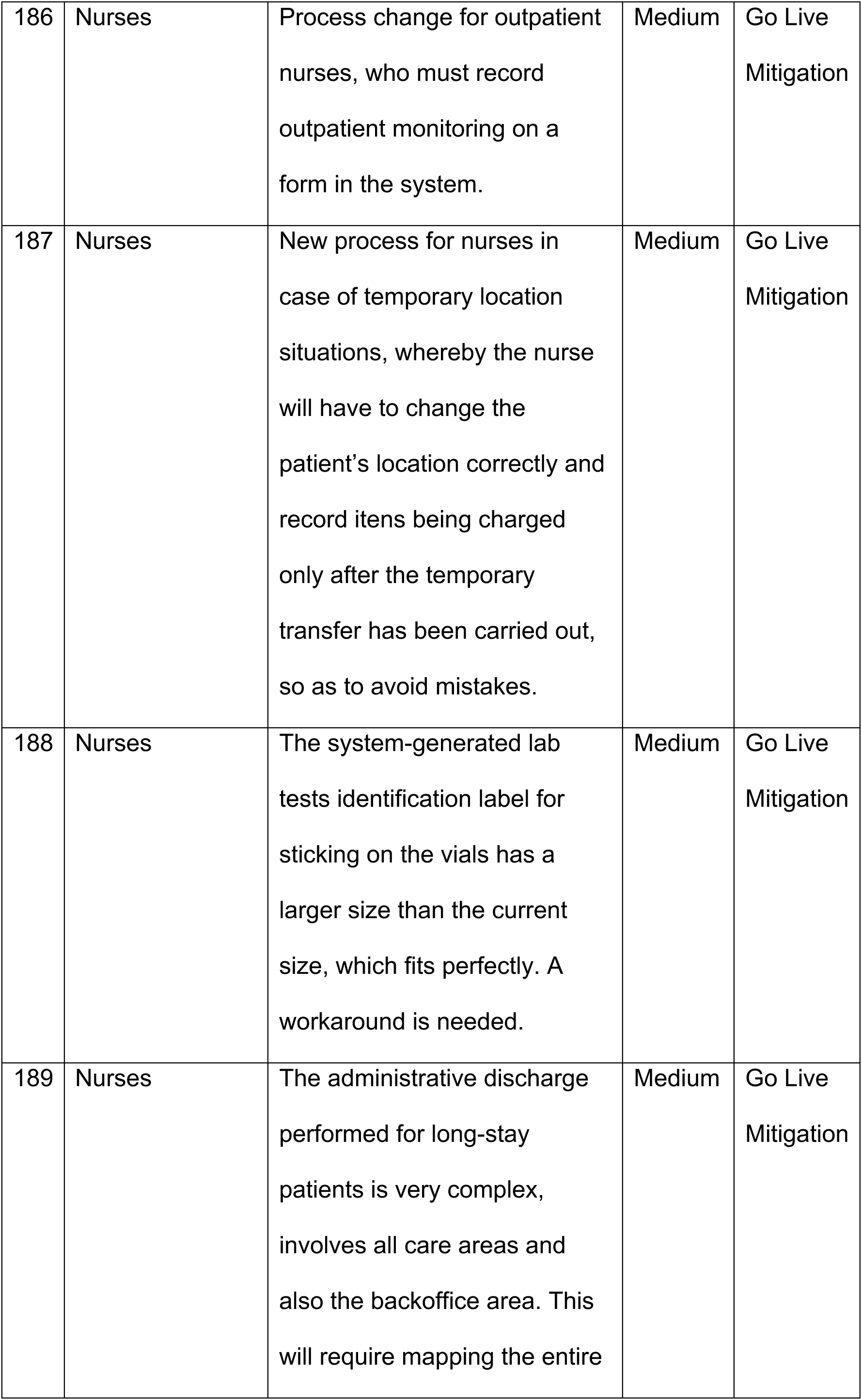
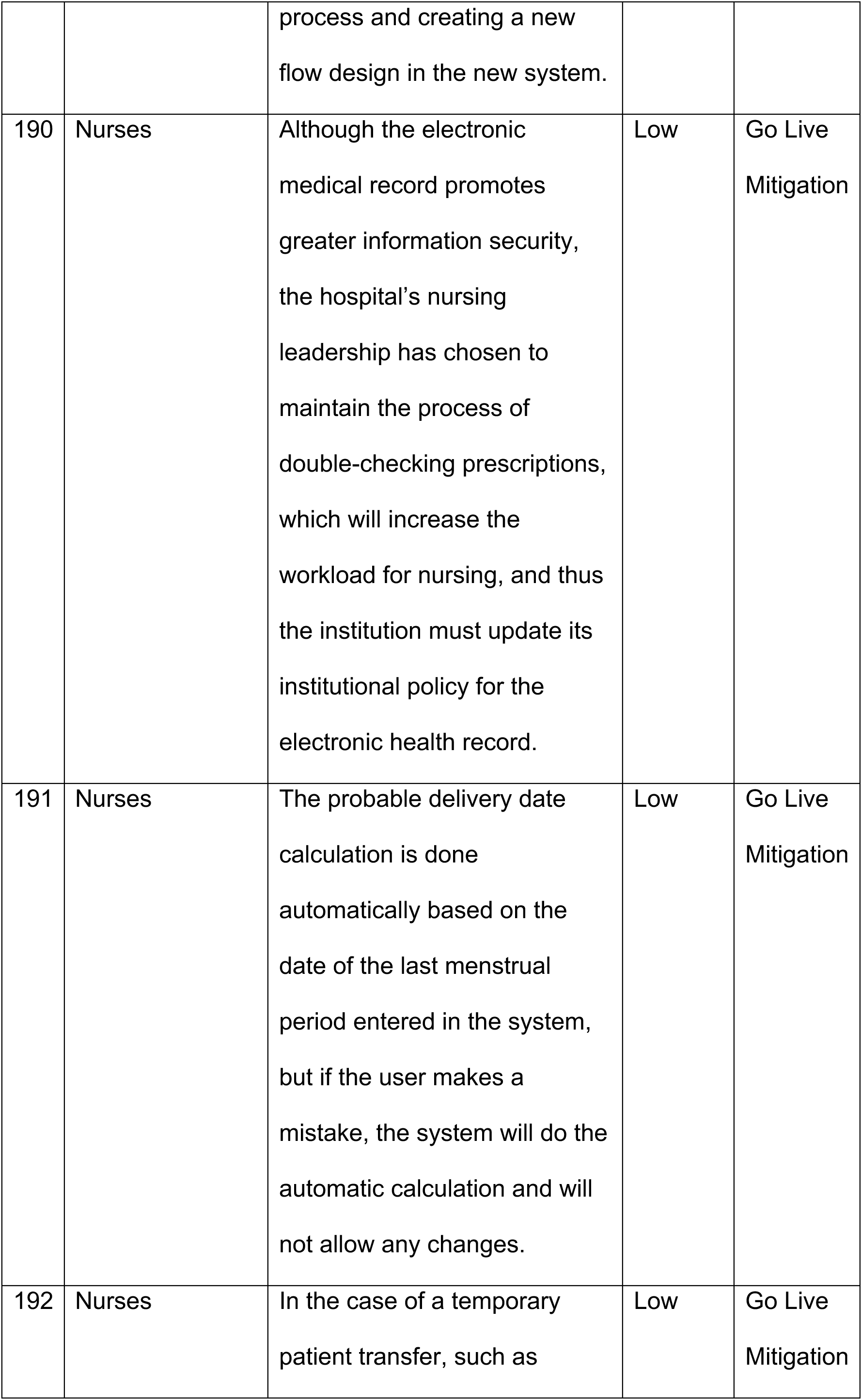
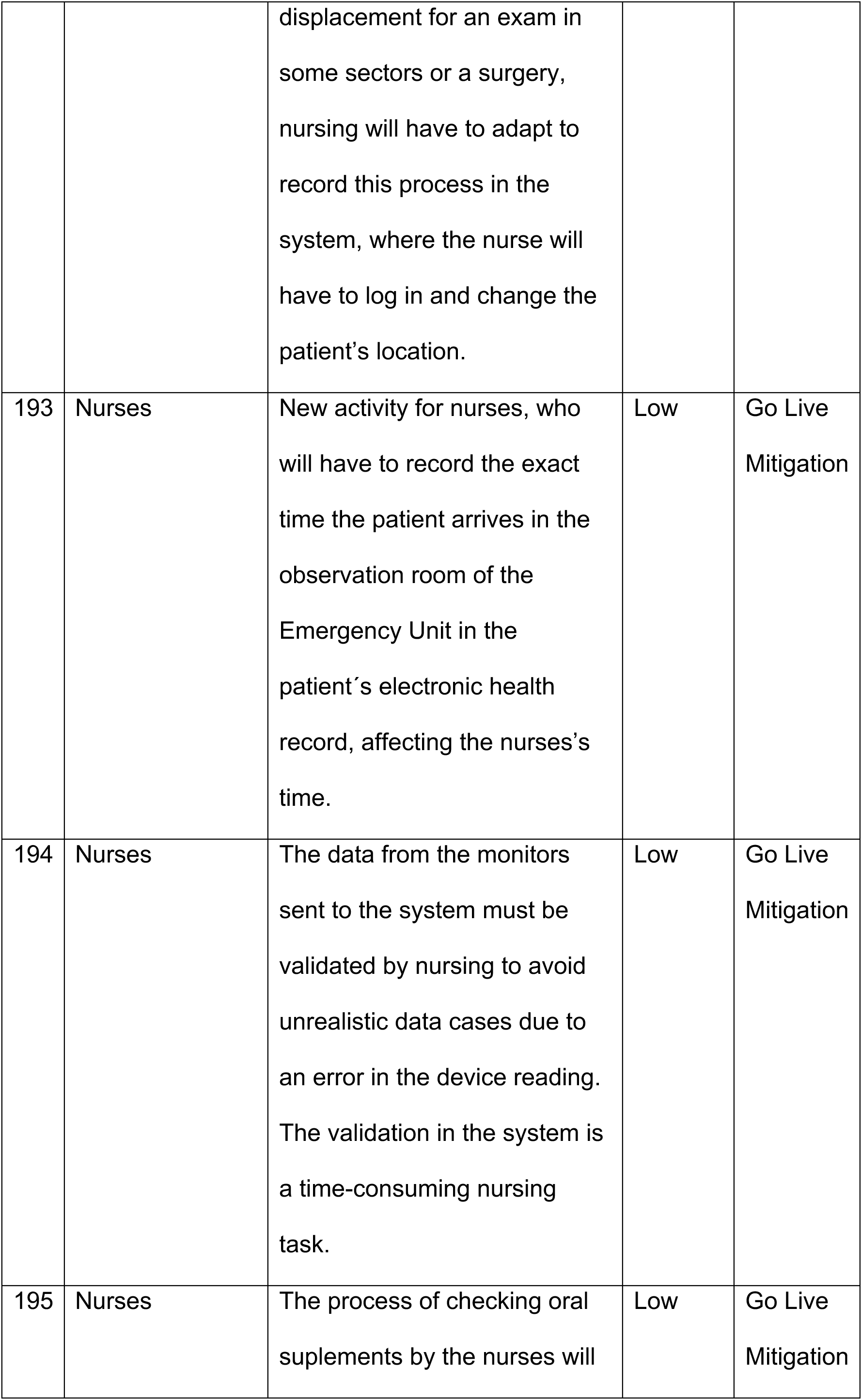
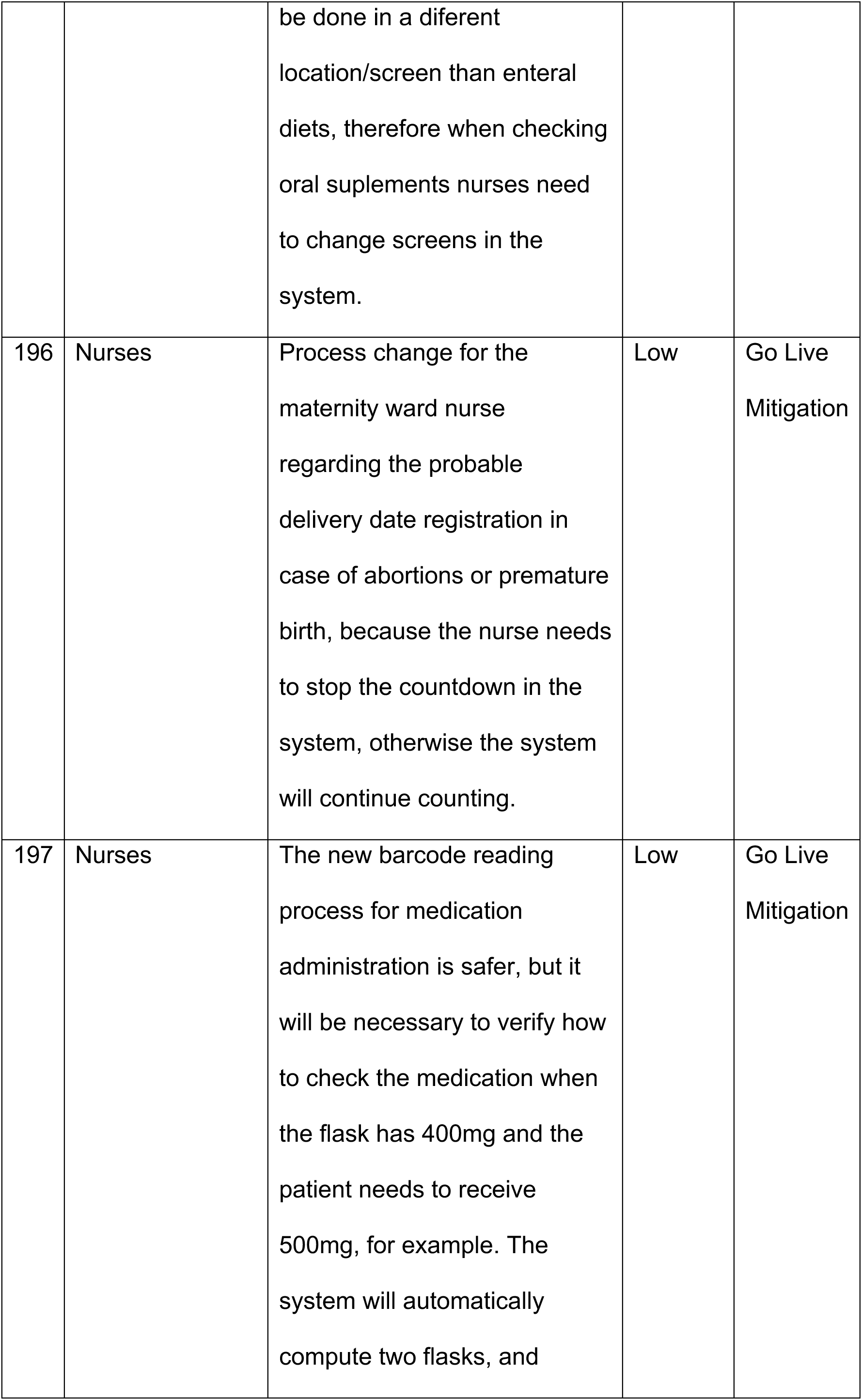
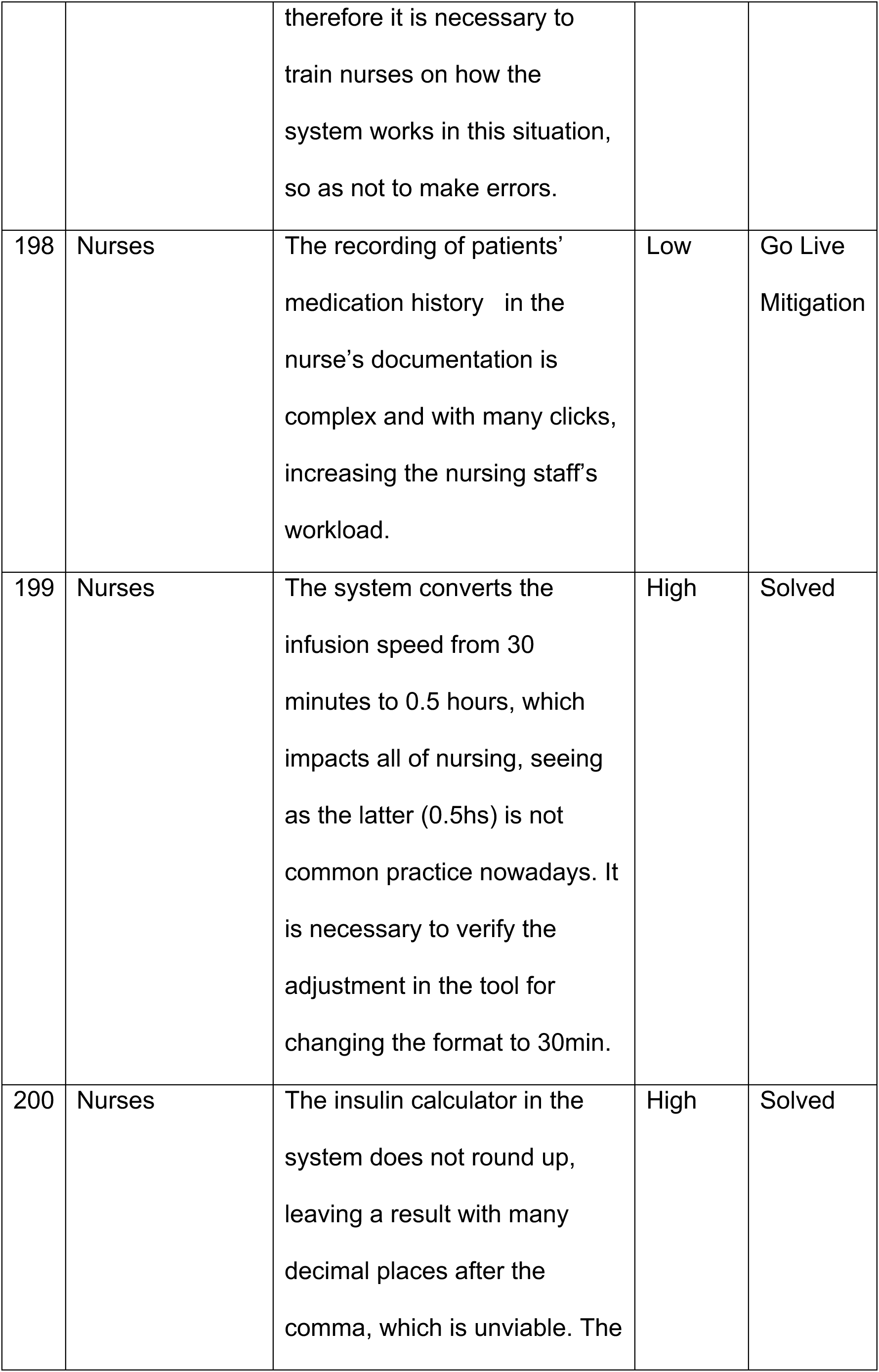
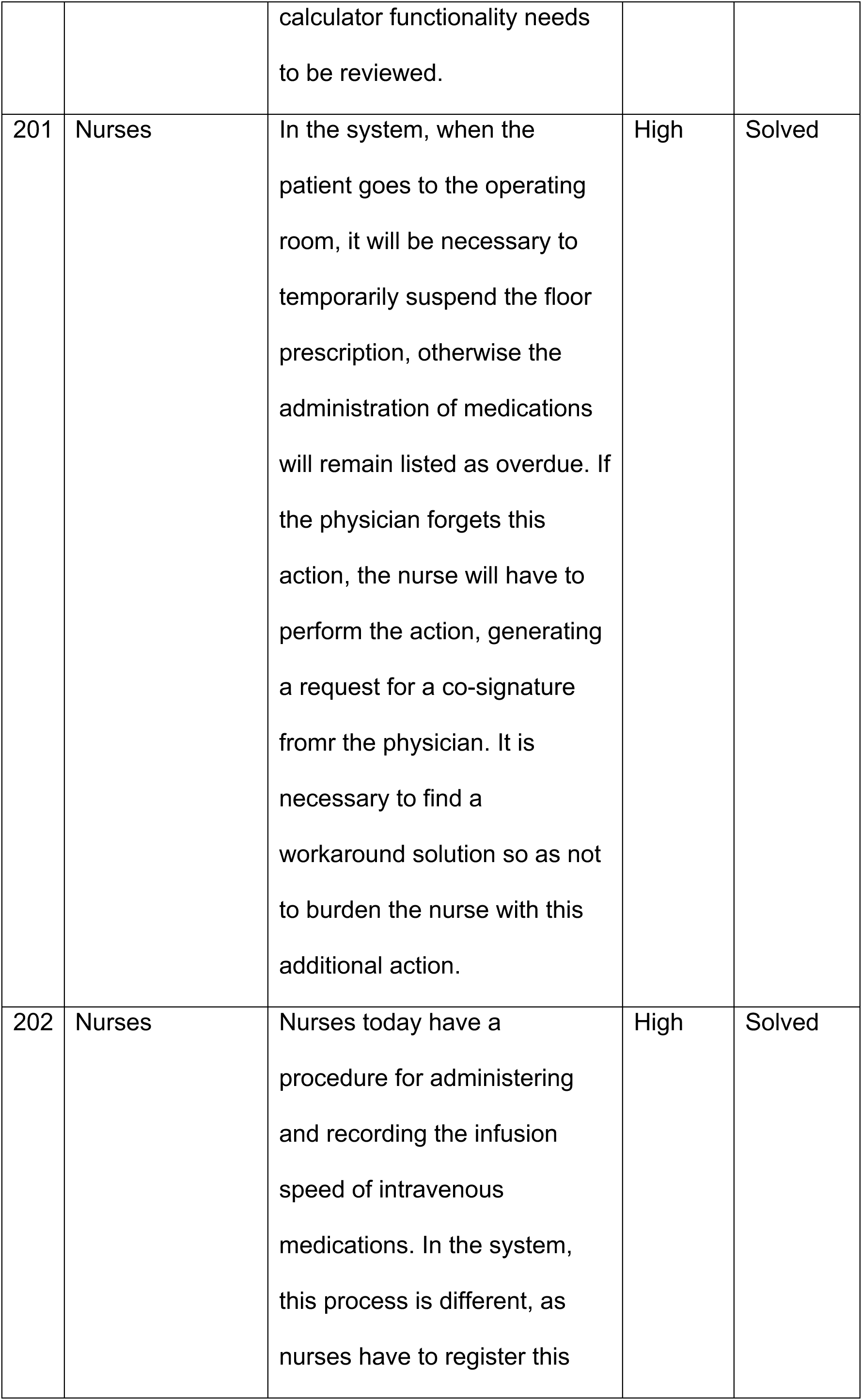
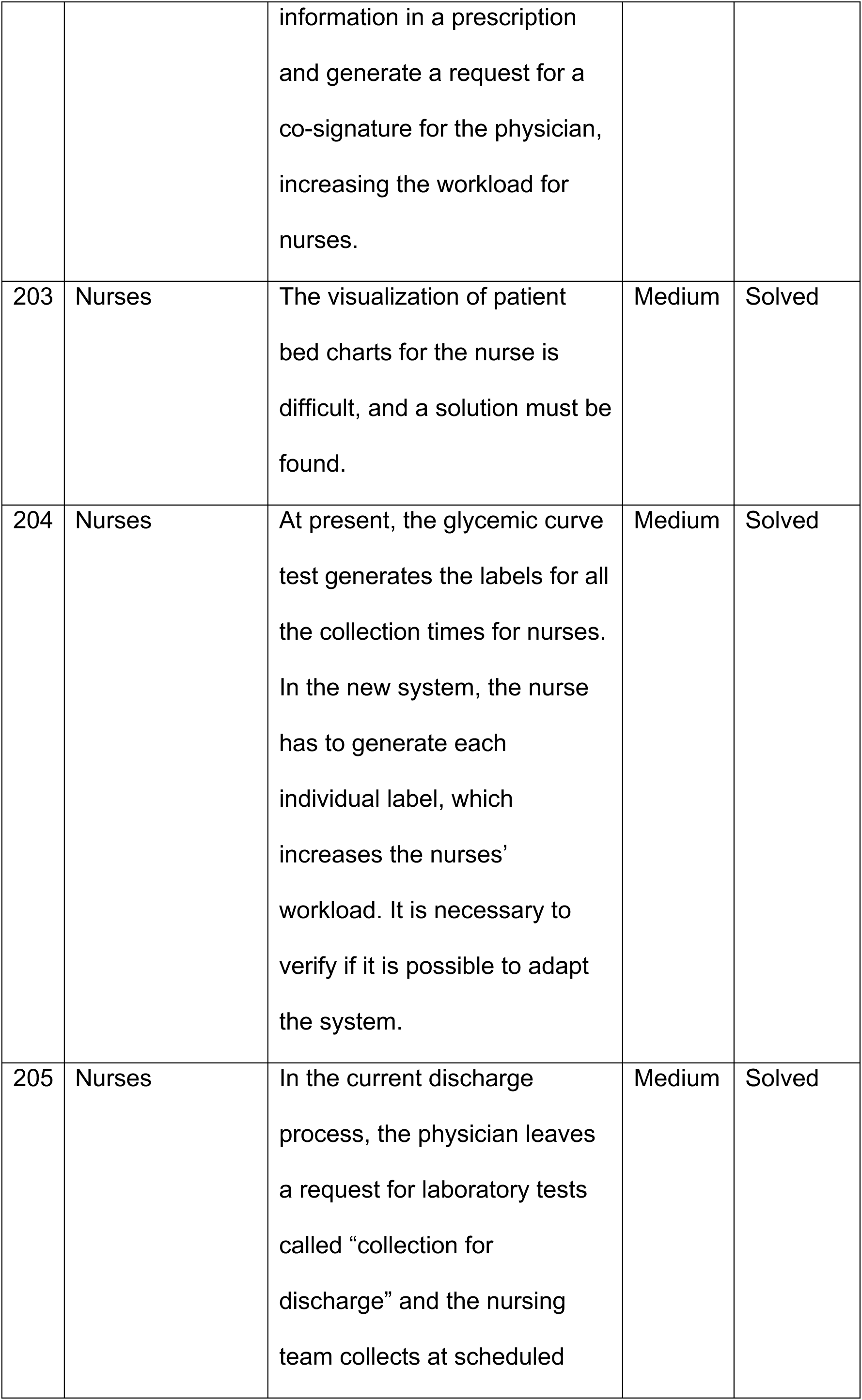
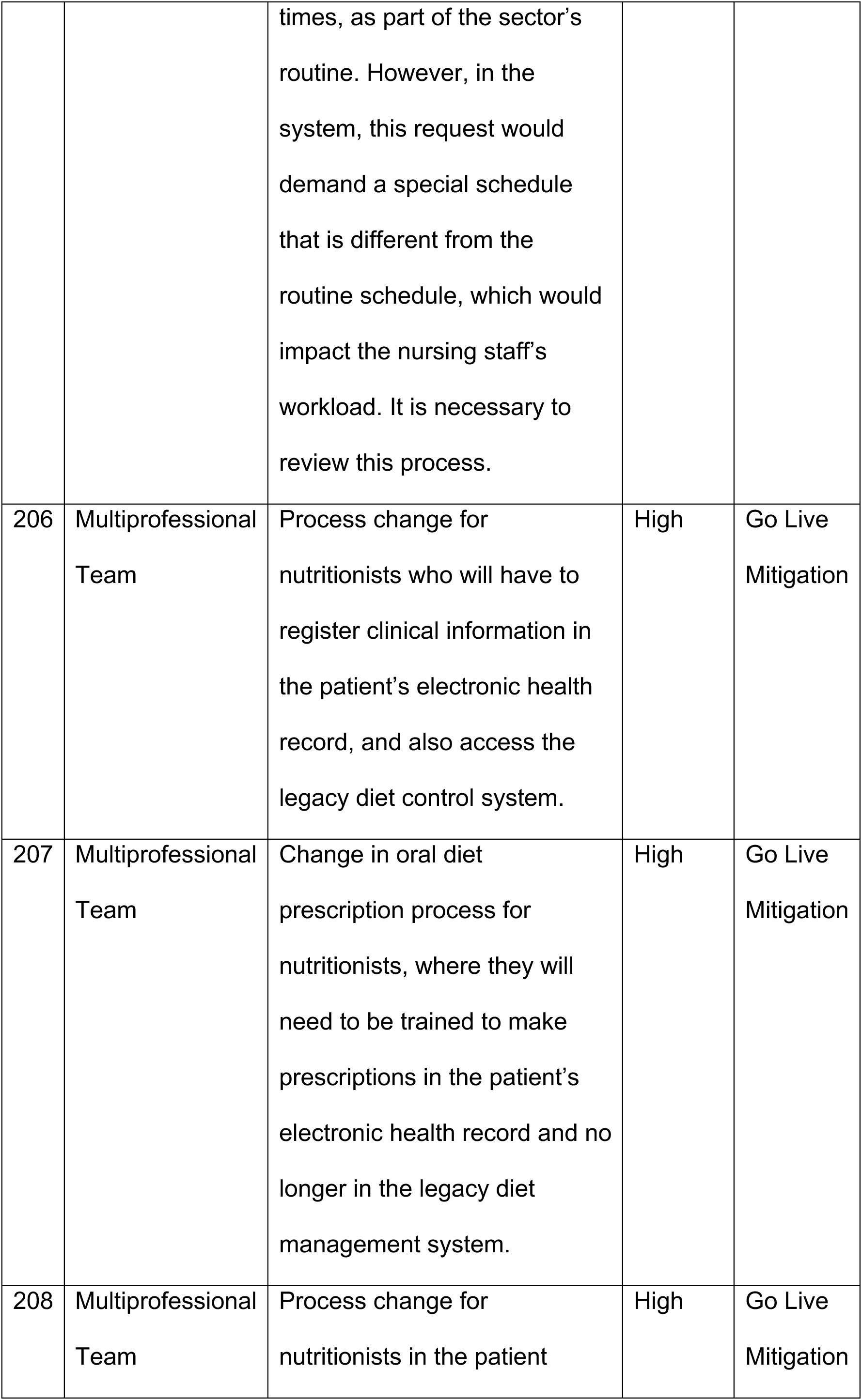
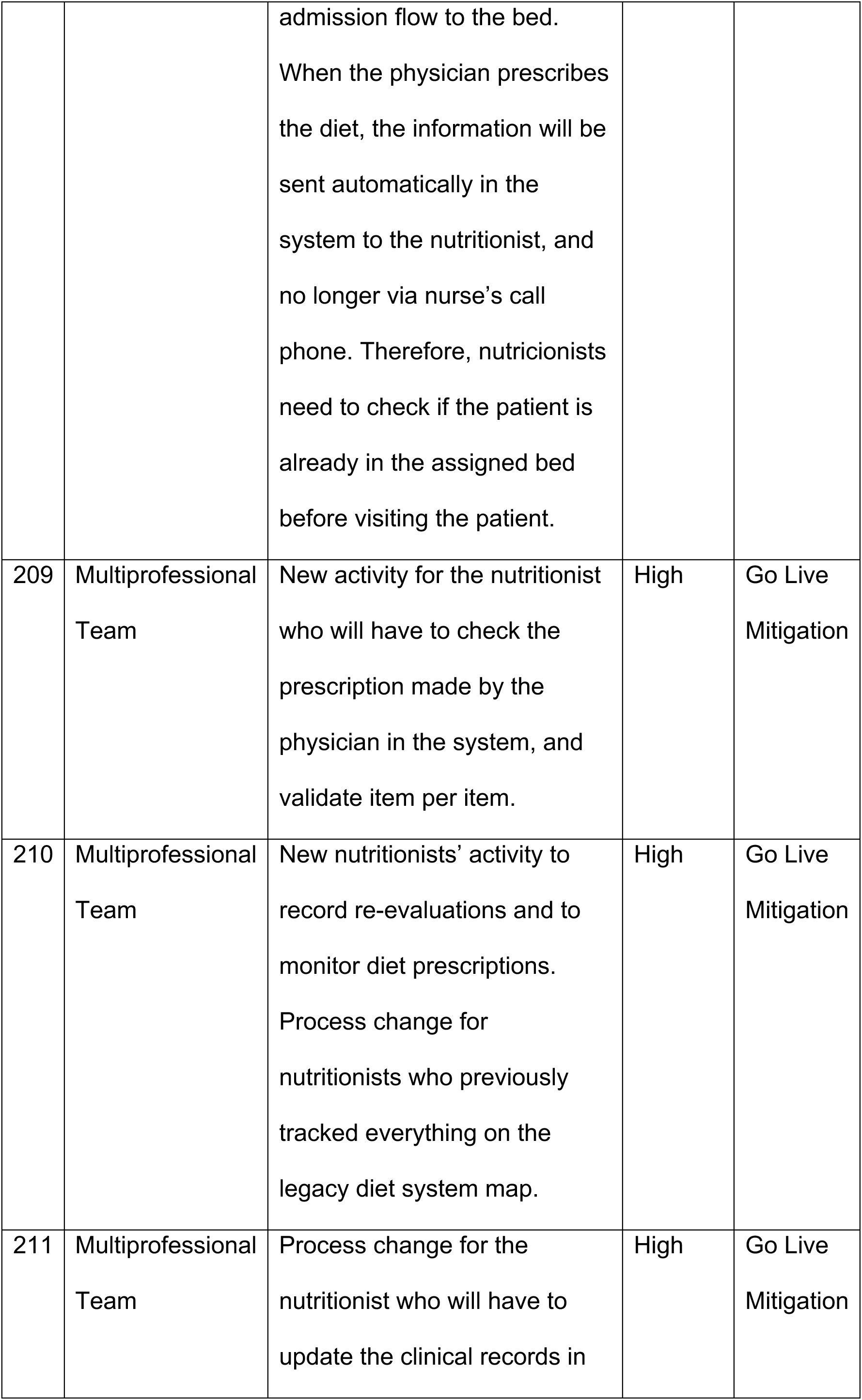
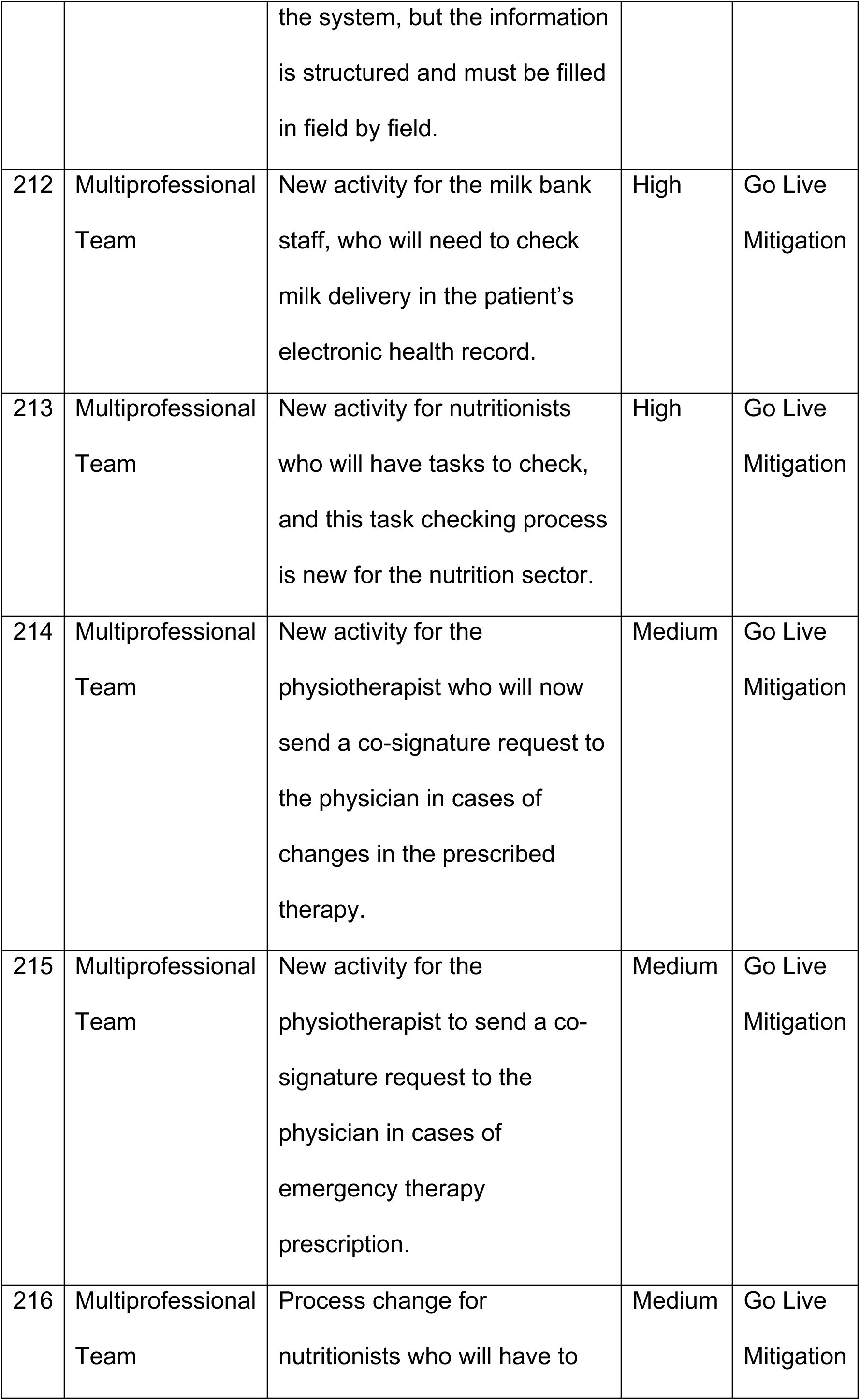
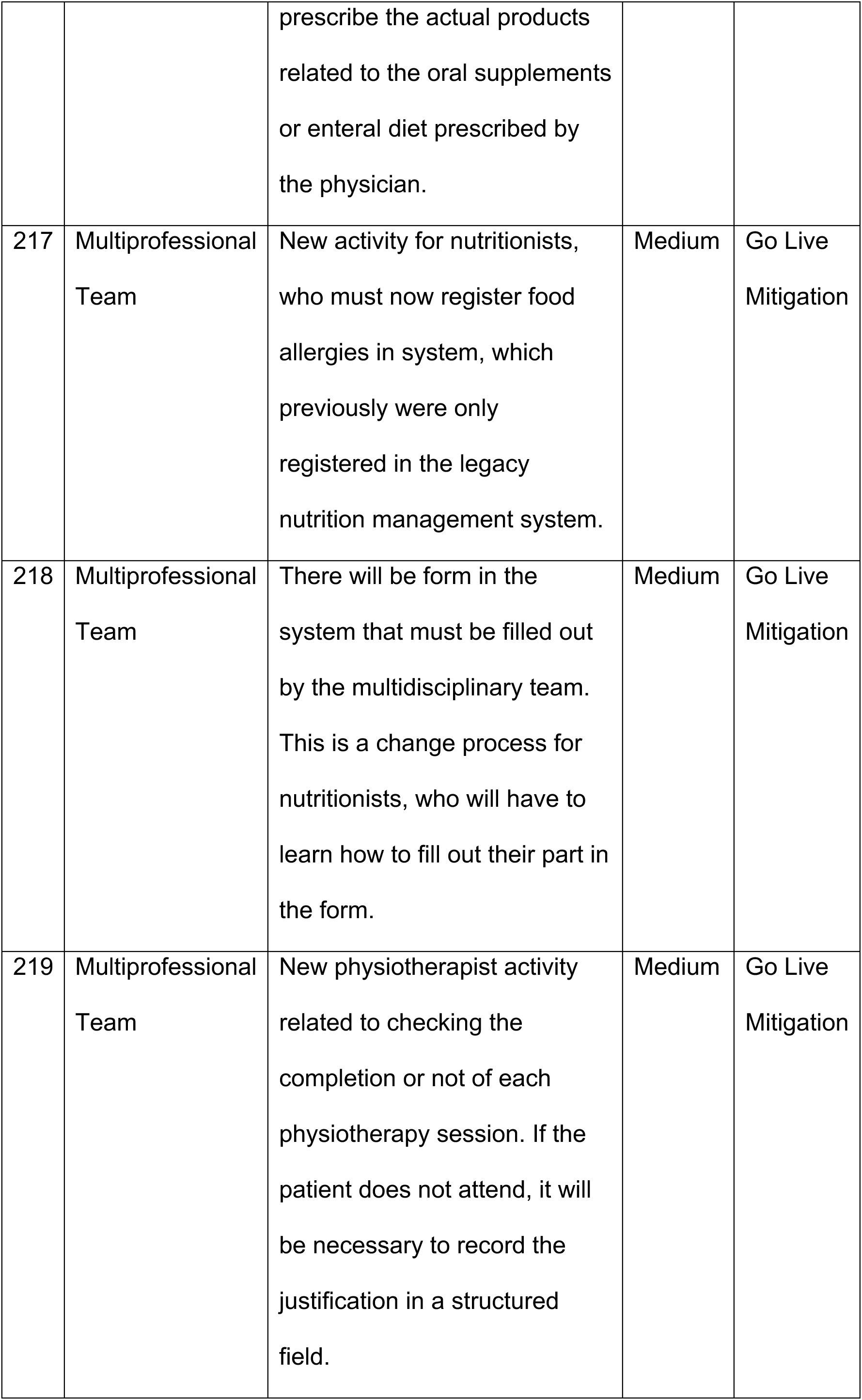
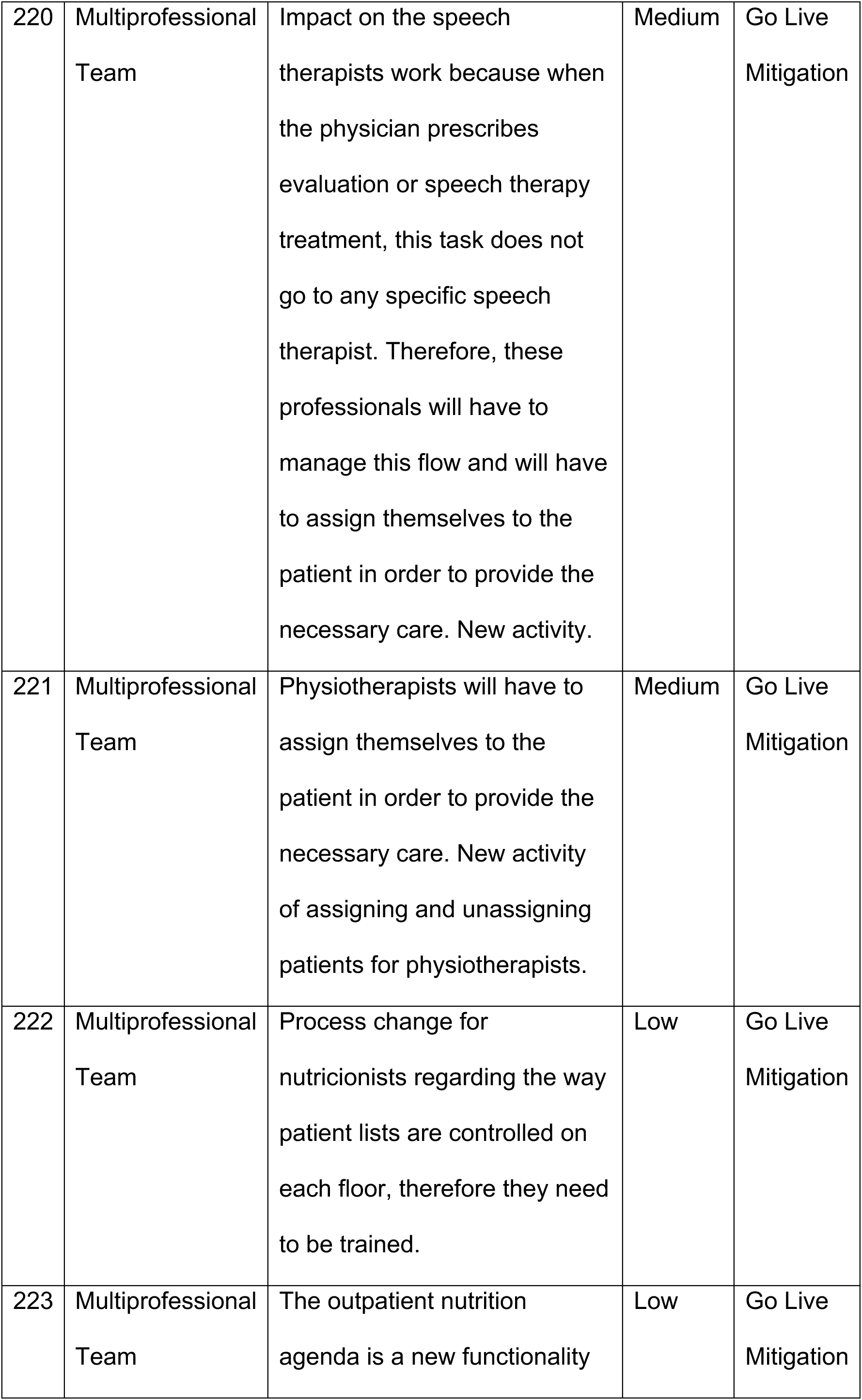
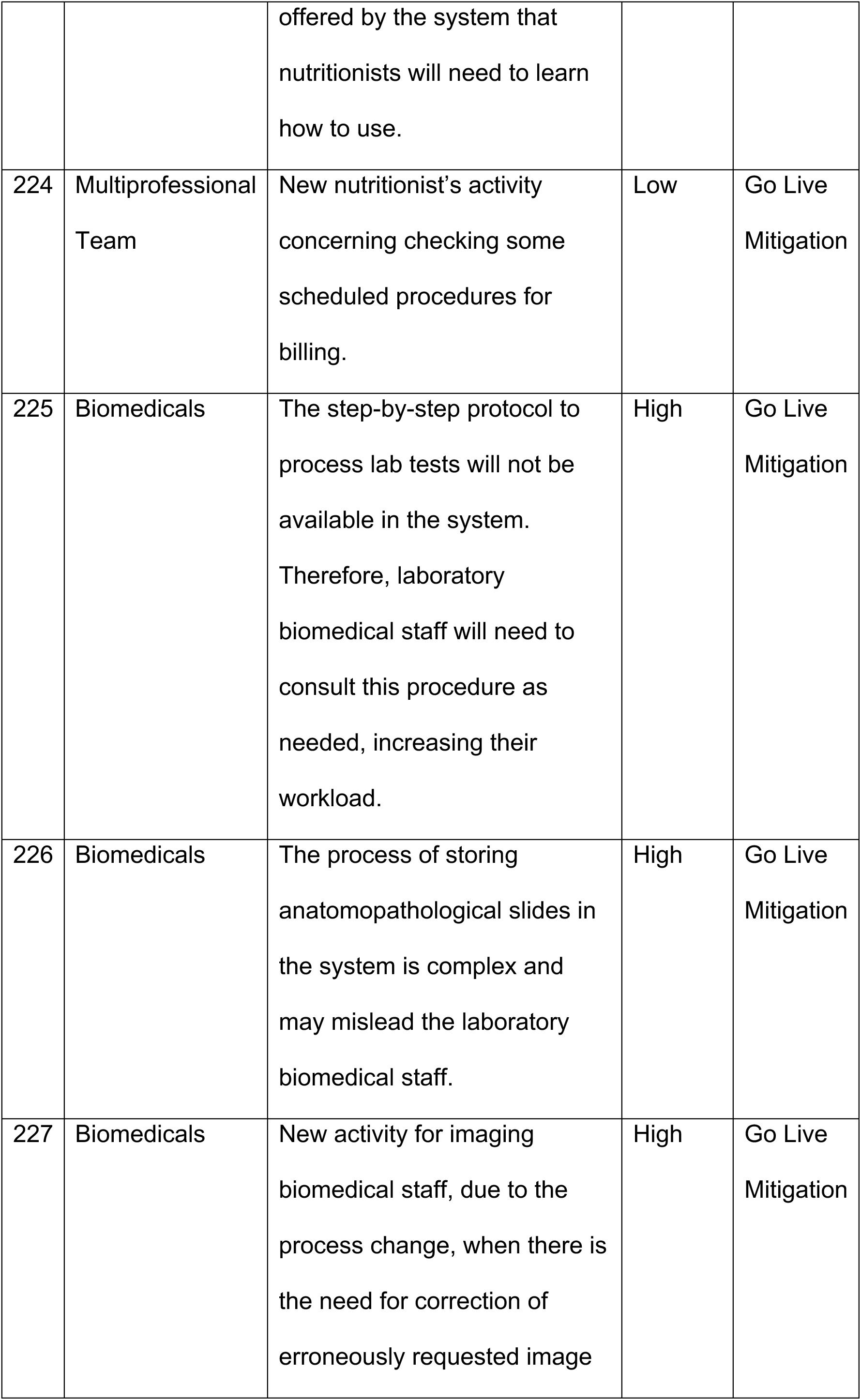
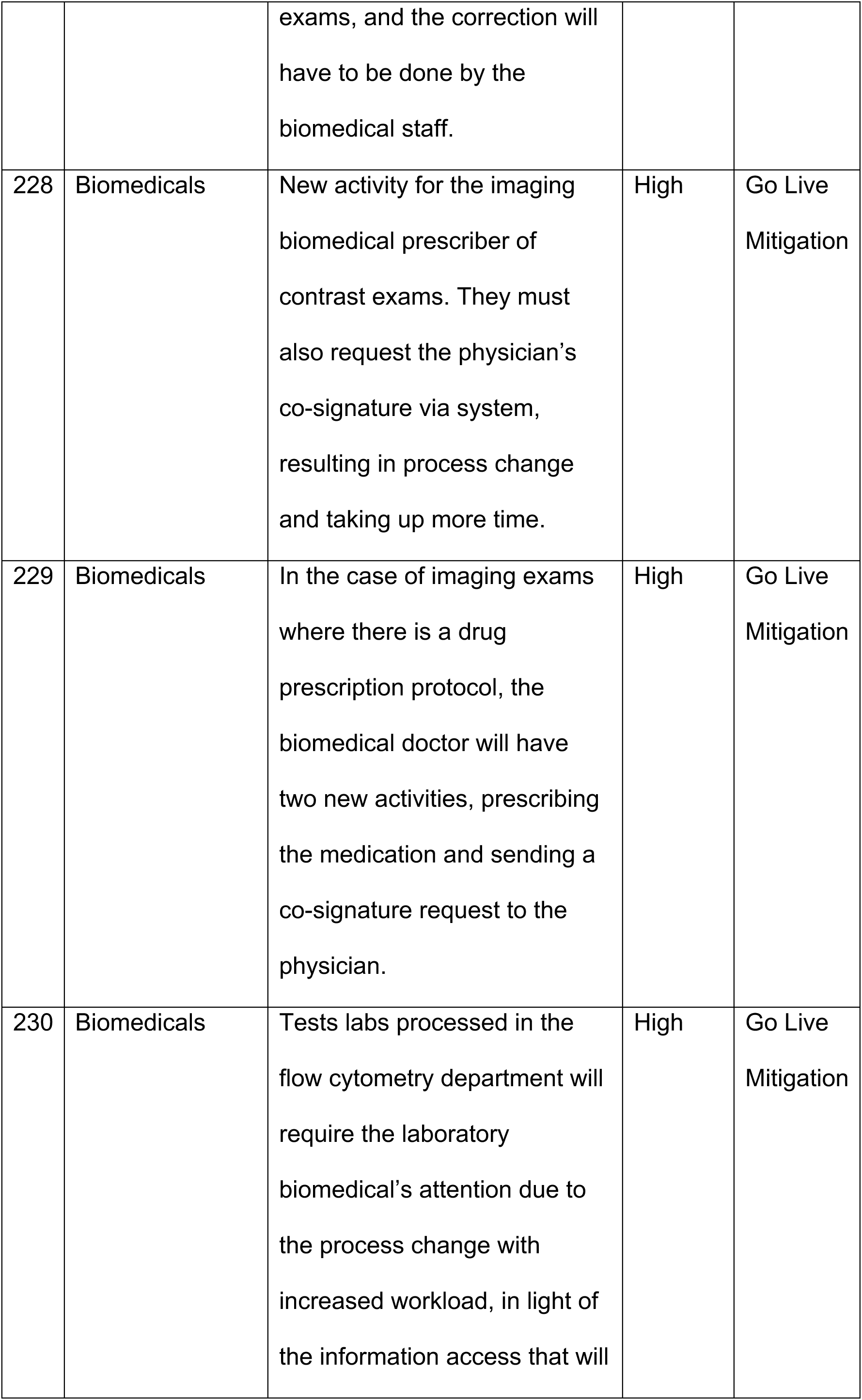
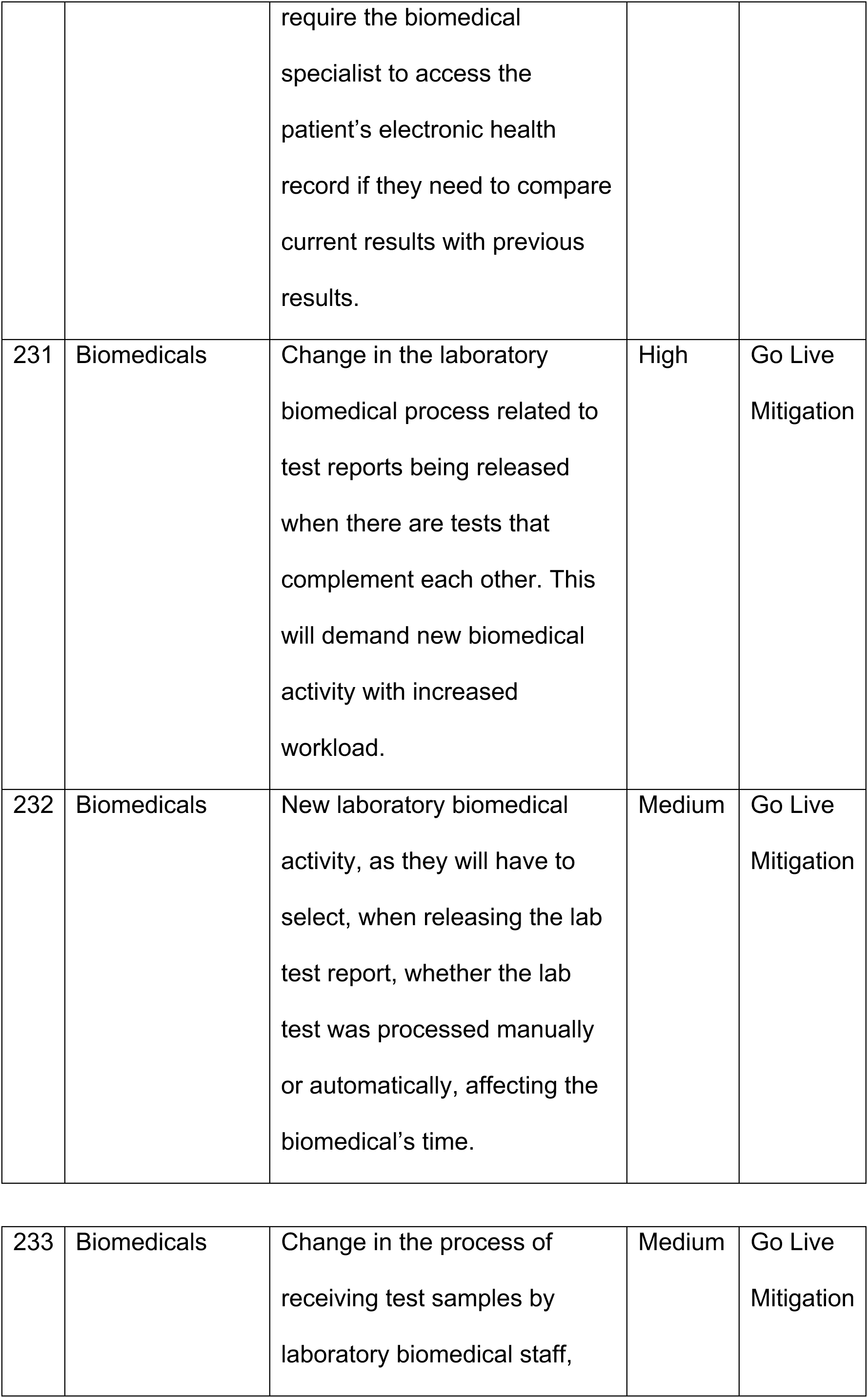
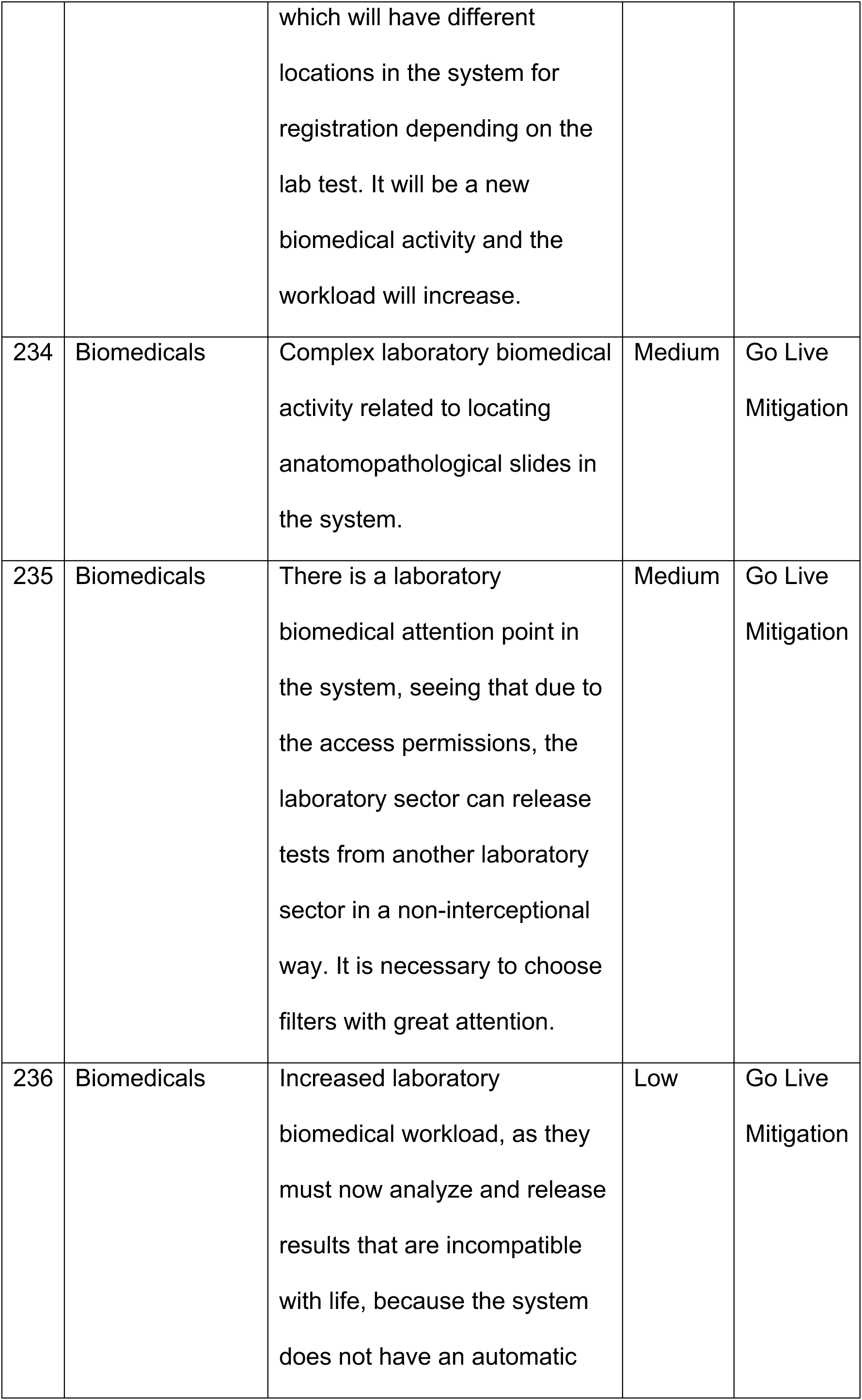
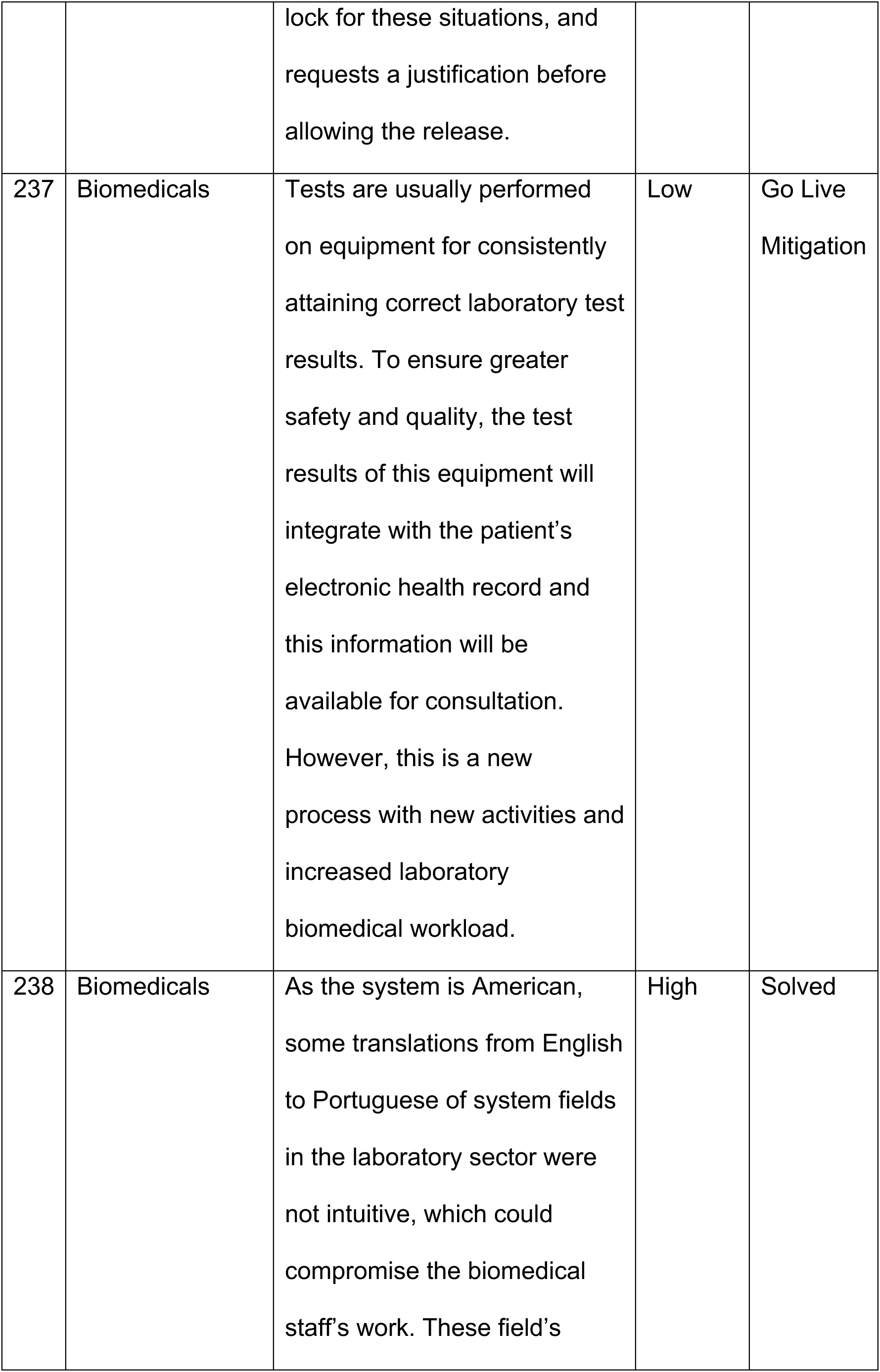
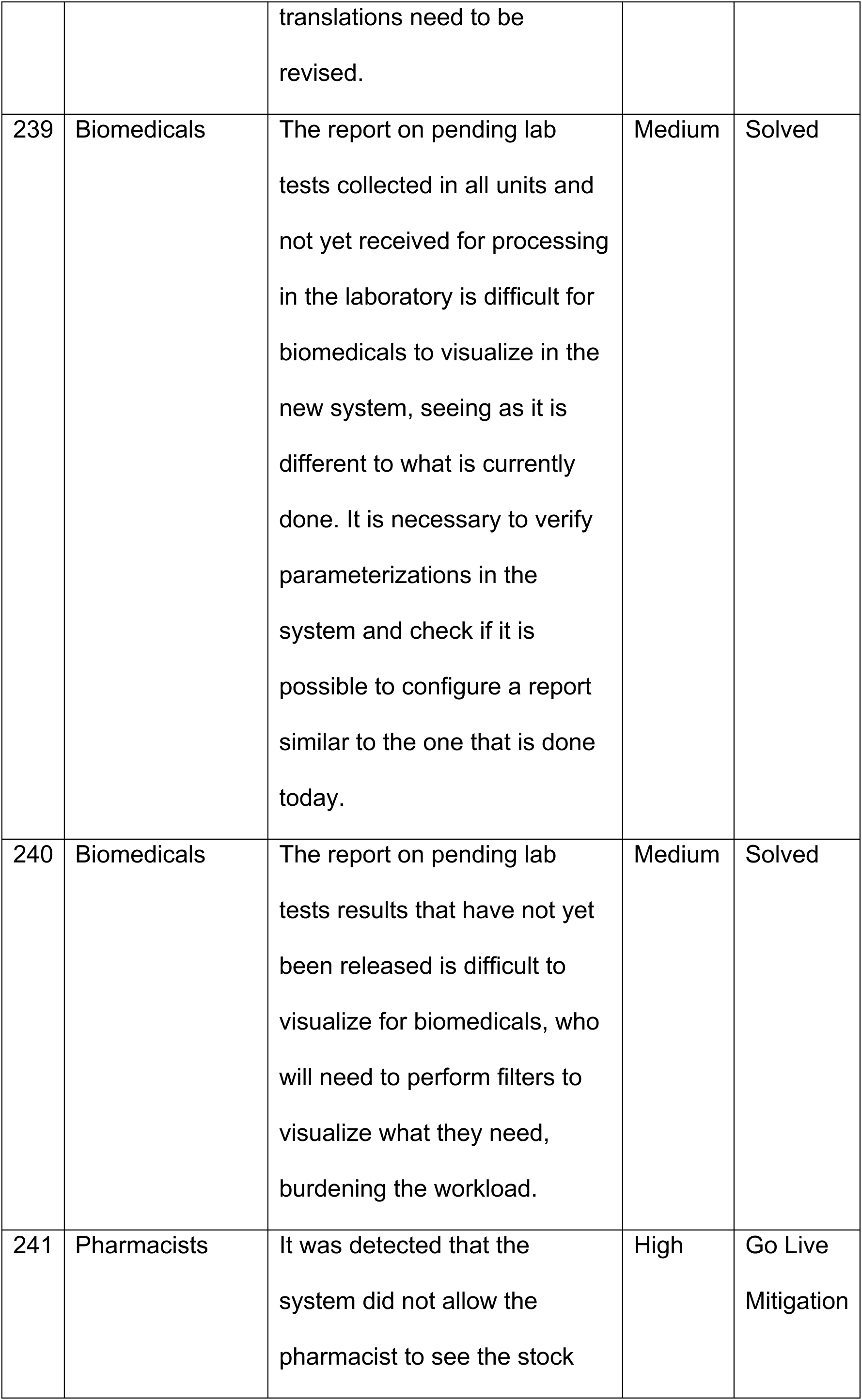
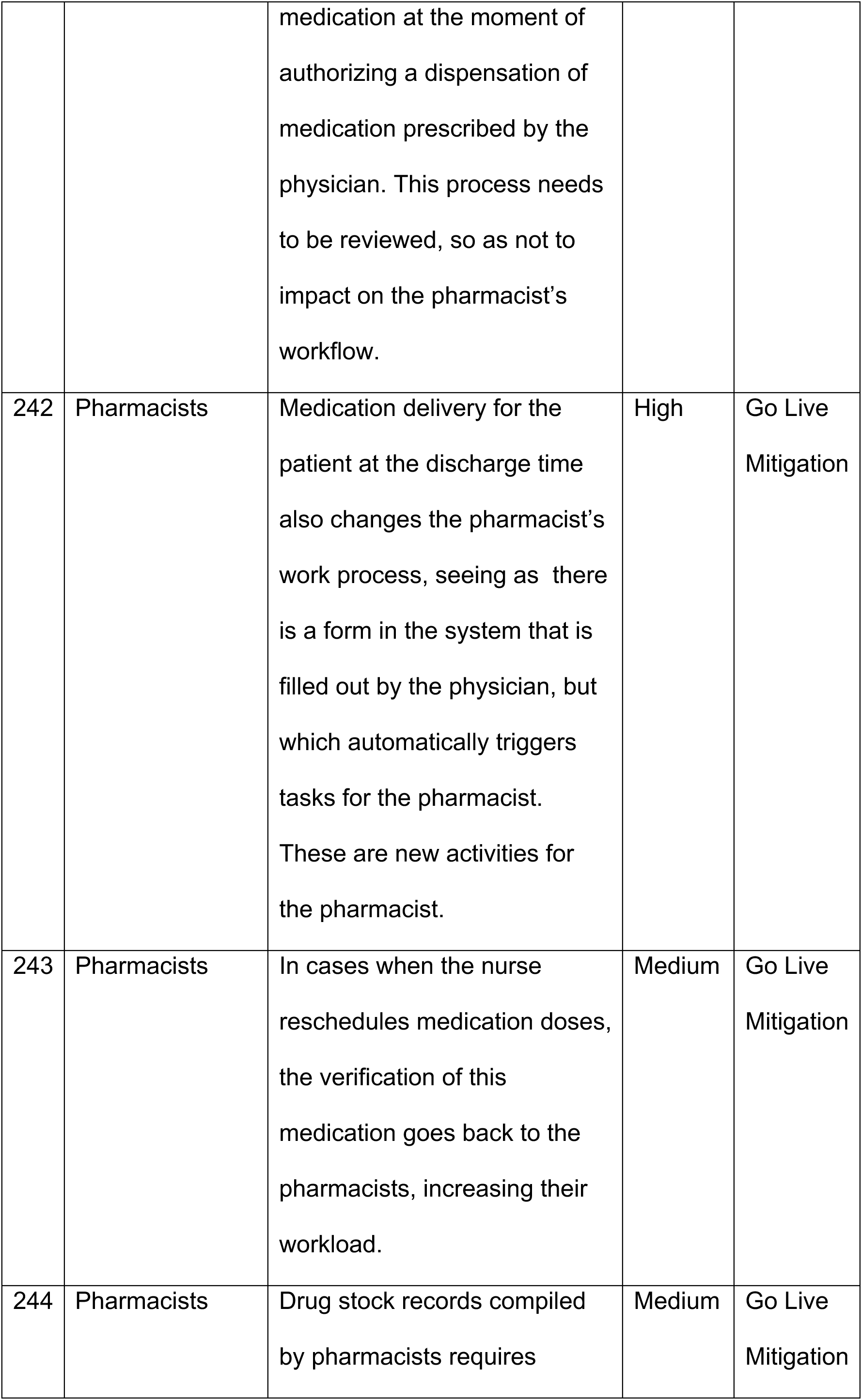
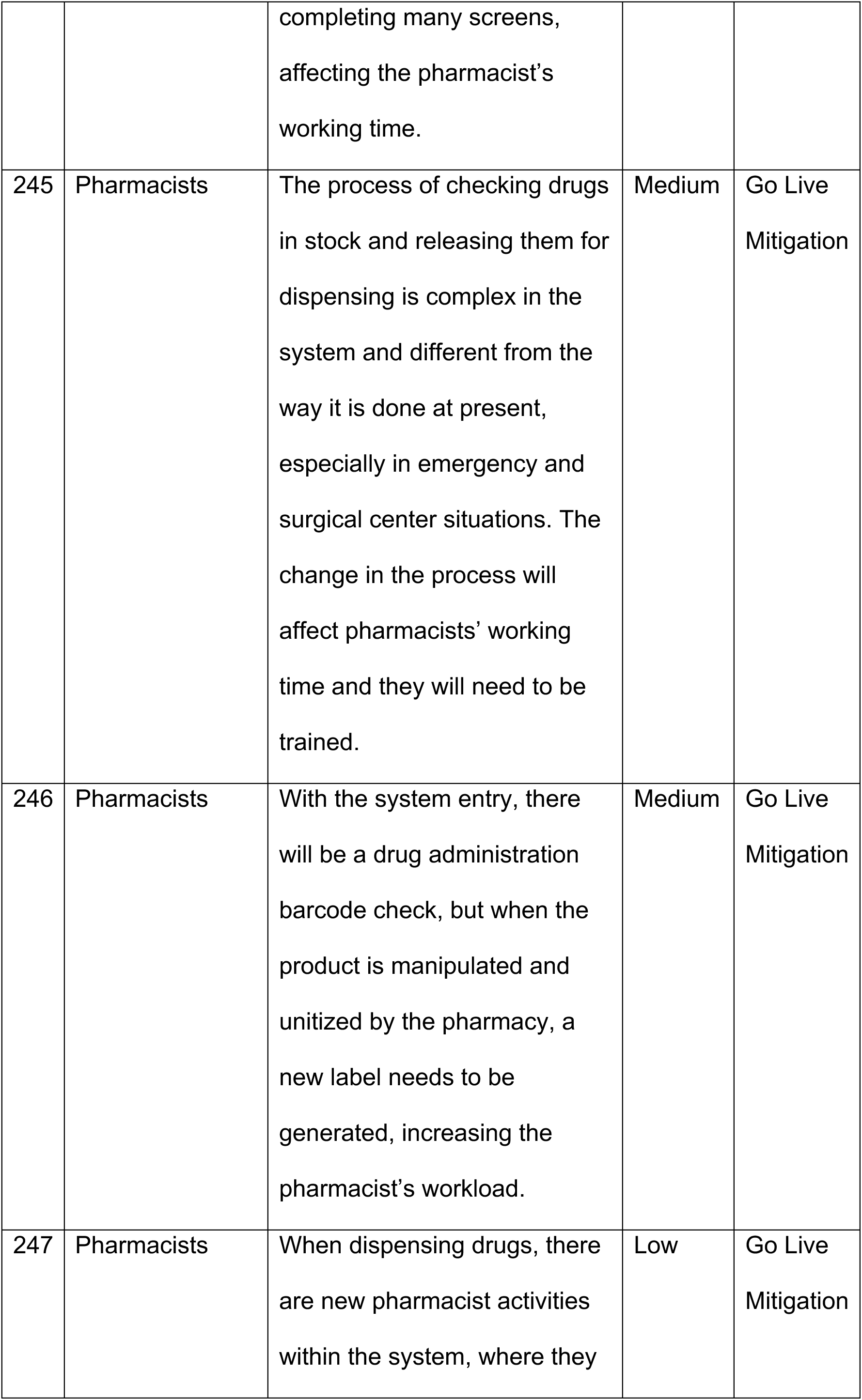
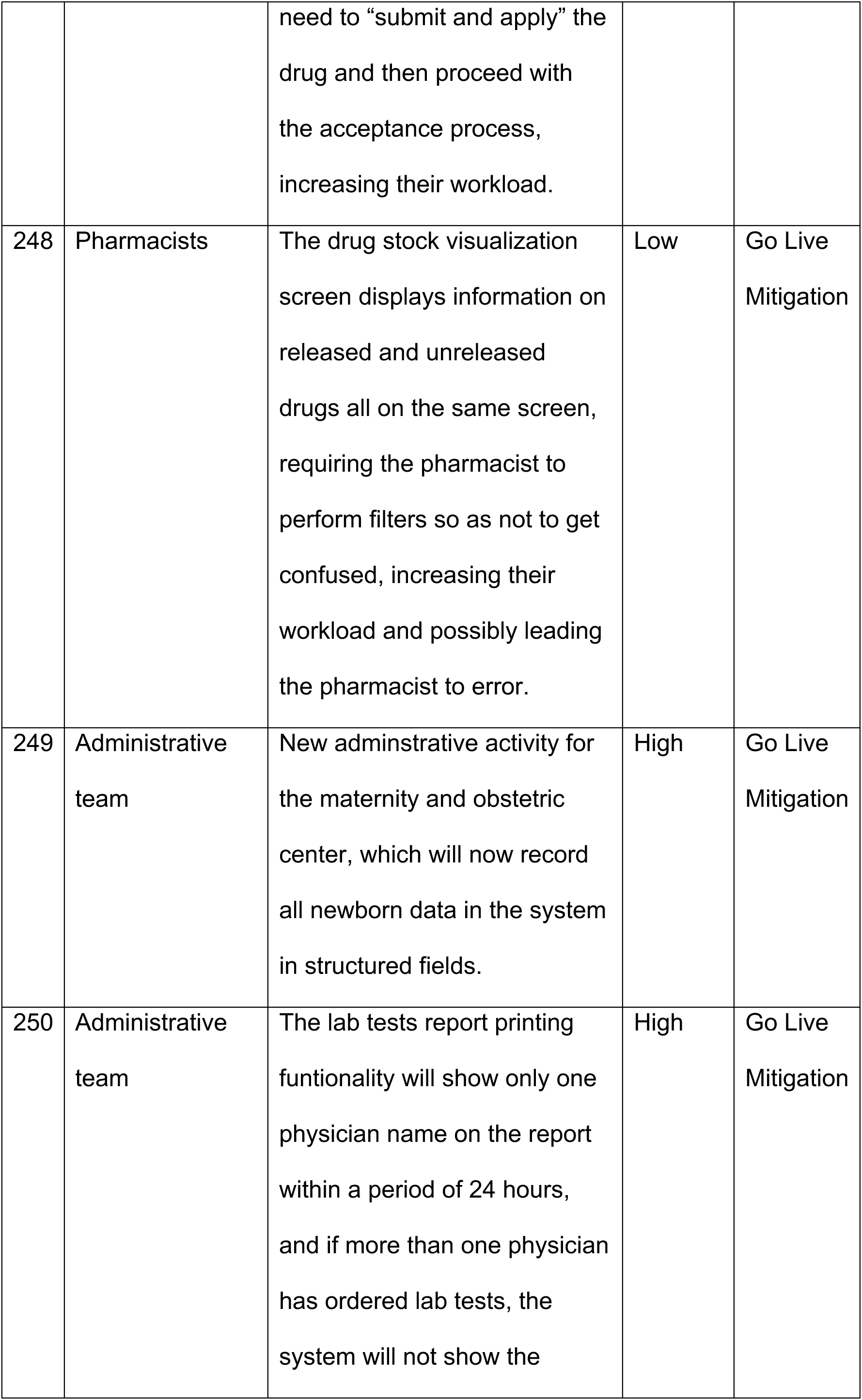
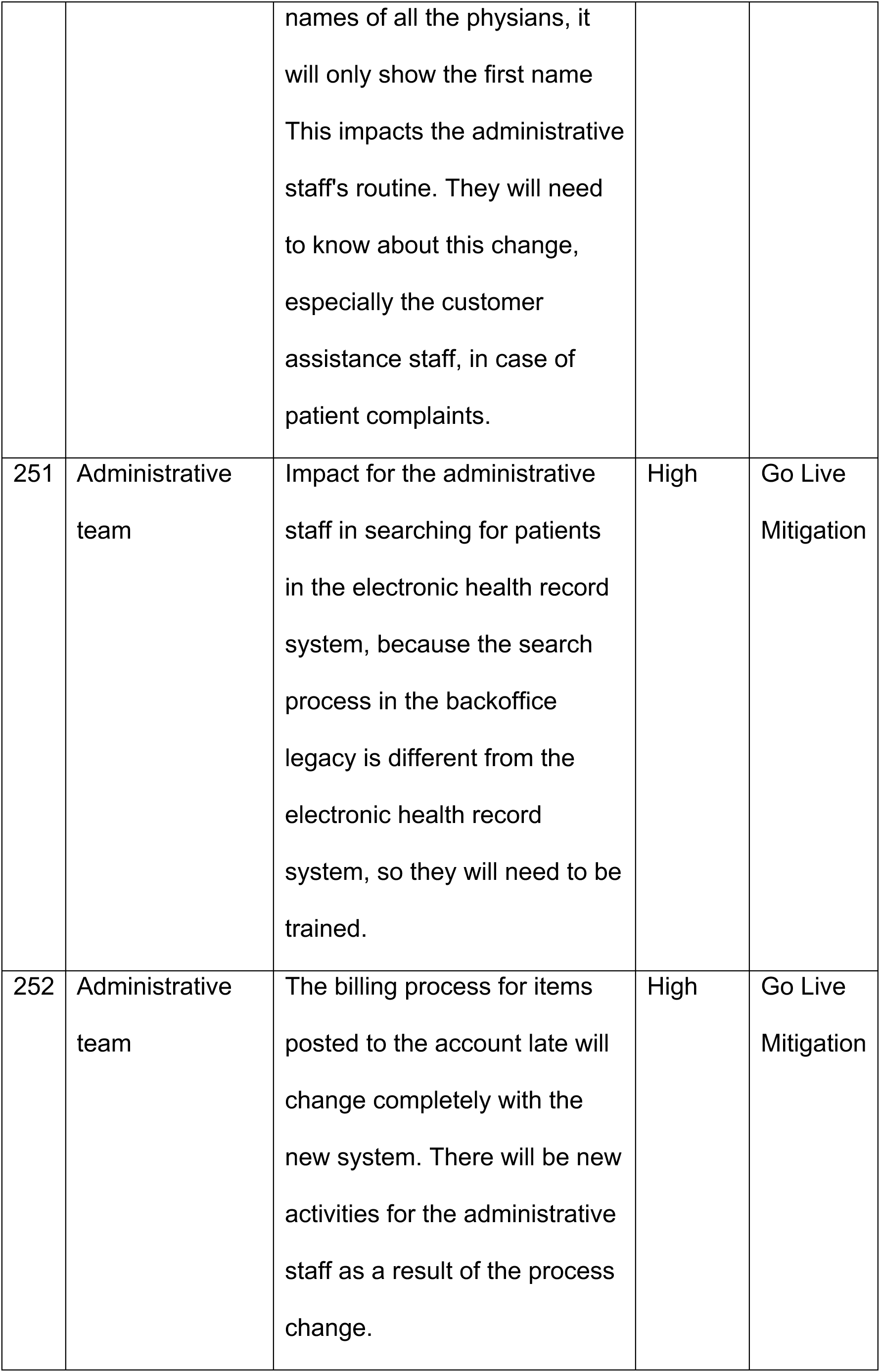
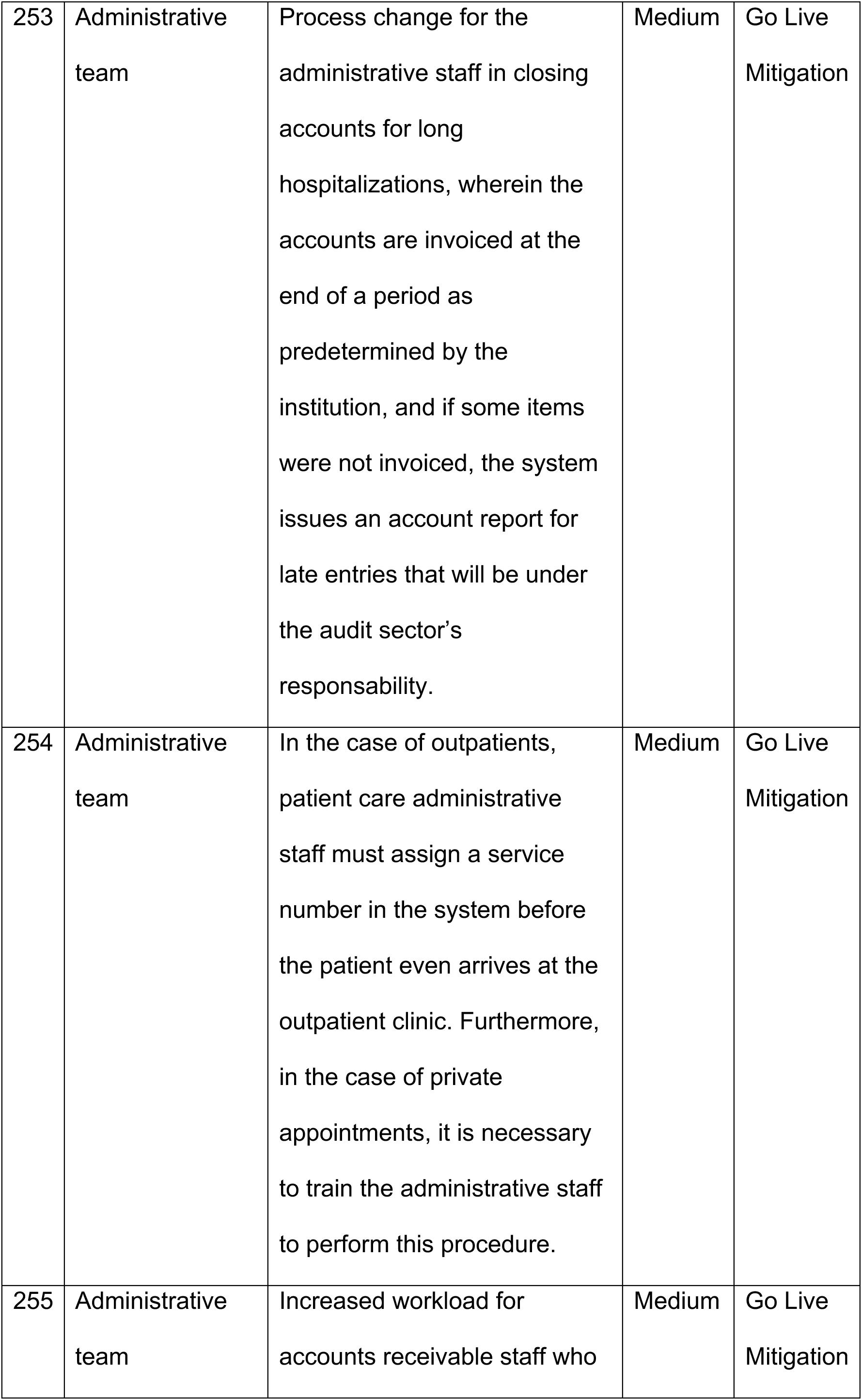
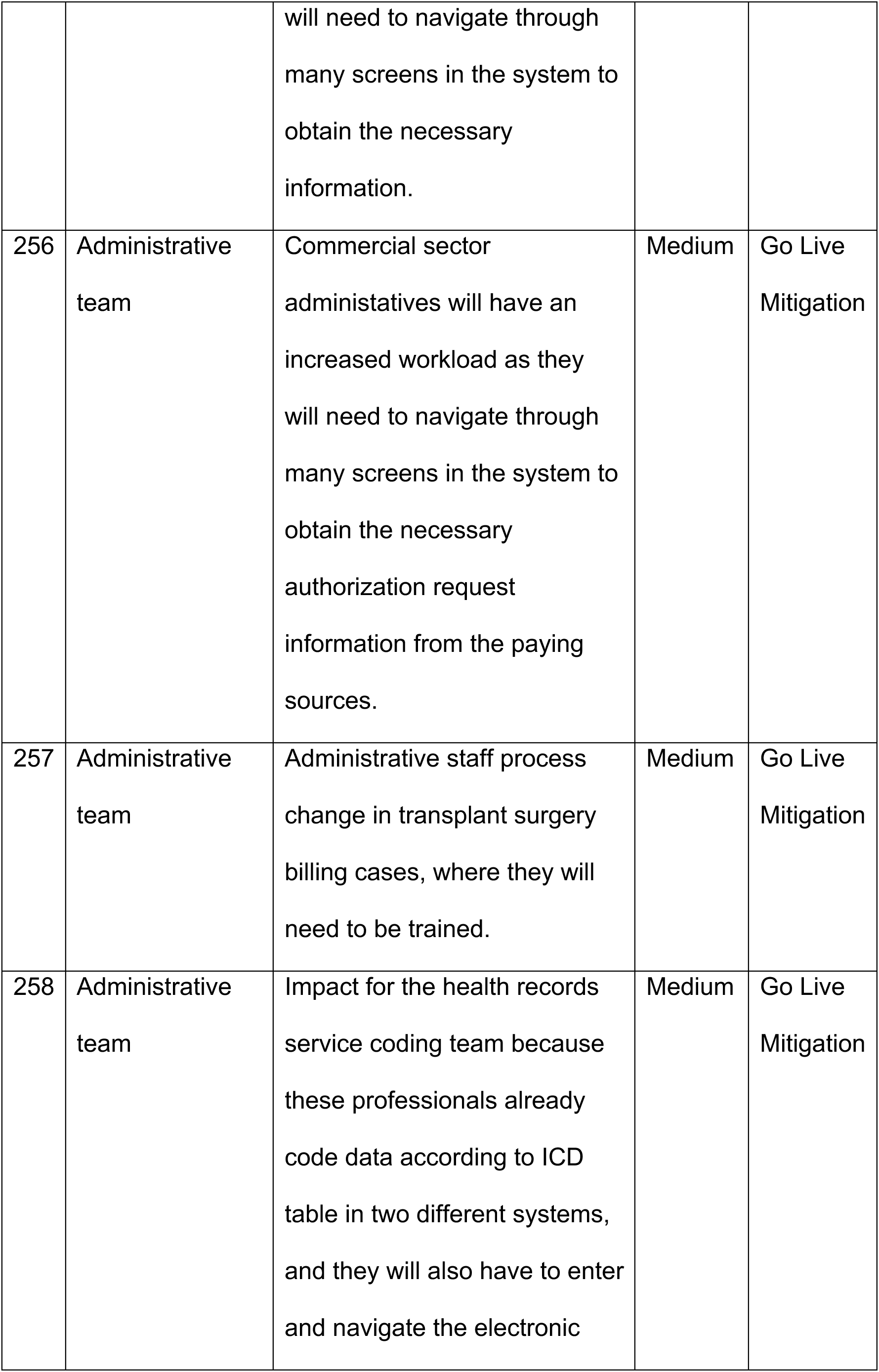
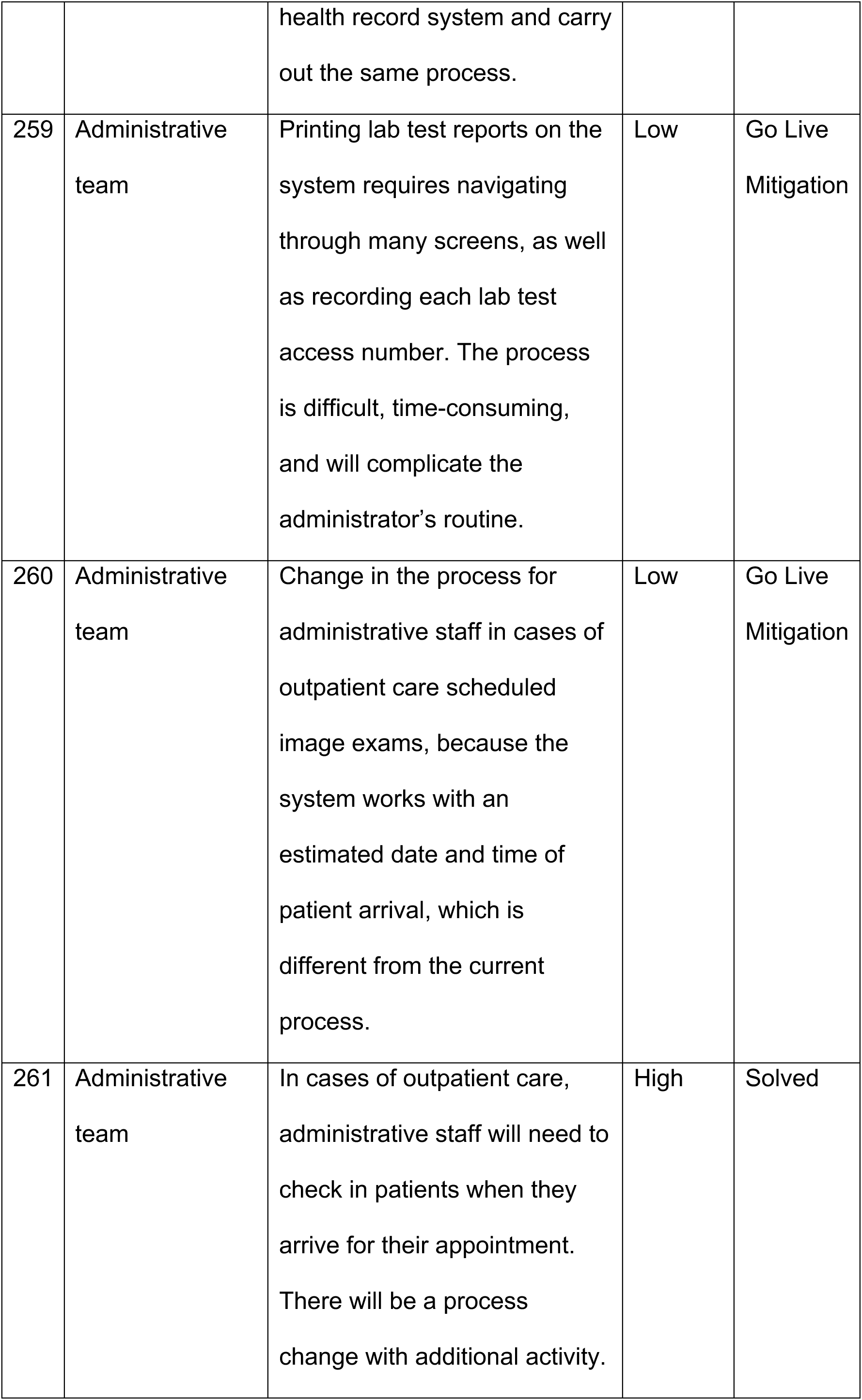
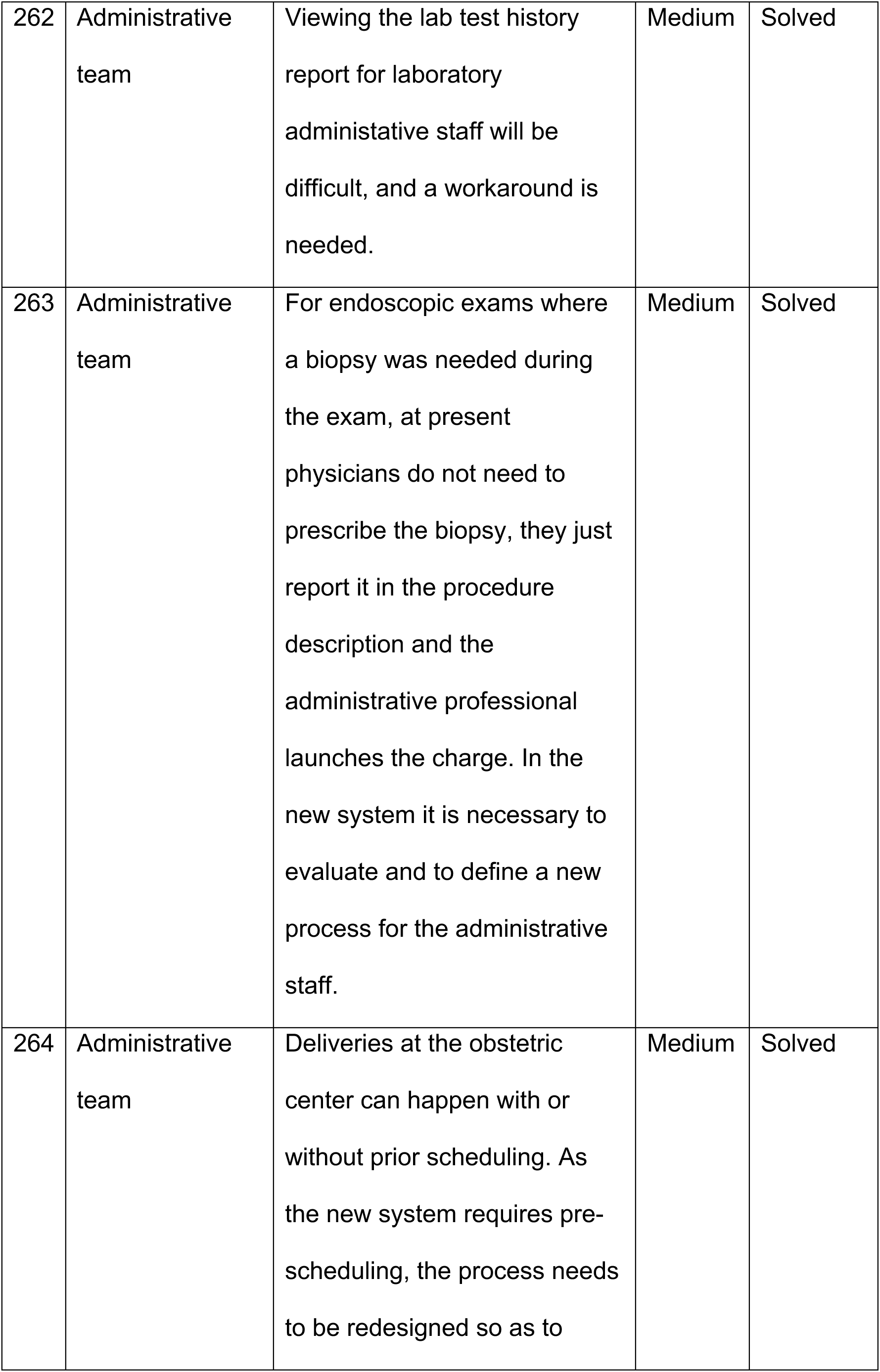
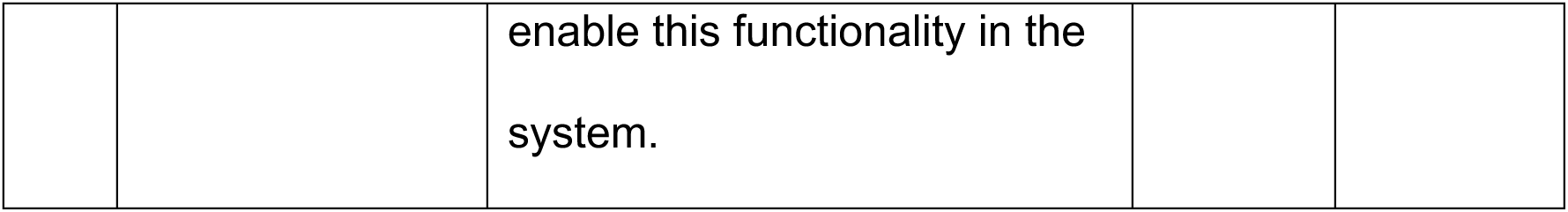
Description of impact according to professional group, criticality level and status.

